# The reciprocal relationship between short- and long-term motor learning and neurometabolites

**DOI:** 10.1101/2024.11.26.24317888

**Authors:** Melina Hehl, Shanti Van Malderen, Svitlana Blashchuk, Stefan Sunaert, Richard A. E. Edden, Stephan P. Swinnen, Koen Cuypers

## Abstract

**Introduction:** Skill acquisition requires practice to stimulate neuroplasticity. Changes in inhibitory and excitatory neurotransmitters, such as respectively gamma-aminobutyric acid (GABA) and glutamate, are believed to play a crucial role in promoting neuroplasticity.

**Methods:** Magnetic resonance spectroscopy (MRS) at 3T, using the MEGA-PRESS sequence, and behavioral data were collected from 62 volunteers. Participants completed a four-week protocol, practicing either complex (n = 32) or simple (n = 30) bimanual tracking tasks (BTT). Neurotransmitter levels and skill levels at baseline, after two, and four weeks of motor training were compared for left and right primary sensorimotor cortex (SM1) and left dorsal premotor cortex (PMd). Furthermore, task-related modulations of neurotransmitter levels in left PMd were assessed.

**Results:** Baseline neurotransmitter levels in motor-related brain regions predicted training success. While lower GABA+ (p=0.0347) and higher Glx (glutamate+glutamine compound) levels (p=0.0234) in left PMd correlated with better long-term learning of simple and complex tasks, respectively, higher GABA+ in right SM1 correlated with complex task learning (p=0.0064). Resting neurometabolite levels changed during the intervention: left SM1 Glx decreased with complex training towards week 4 (p=0.0135), while right SM1 Glx was increased at week 2 (p=0.0043), regardless of training type. Group-level analysis showed no task-related neurometabolite modulation in the left PMd. However, individual baseline GABA+ and Glx modulation influenced short-term motor learning (interaction: p=0.0213).

**Conclusion:** These findings underscore the importance of an interplay between inhibitory and excitatory neurotransmitters during motor learning and suggest potential for future personalized approaches to optimize motor learning.

**Key points:**

- ***Neurotransmitter dynamics predict training success:*** Baseline levels of GABA+ and Glx in motor-related brain regions (left PMd and right SM1) were found to predict long-term learning success, with specific patterns correlating with better performance in both simple and complex motor tasks.
- ***Training-induced changes in neurometabolites:*** Resting levels of Glx in SM1 changed significantly during the motor training, with a decrease in left SM1 Glx after four weeks of complex training, and an increase in right SM1 Glx after two weeks for both the simple and complex training group, indicating dynamic adaptations in response to motor training.
- ***Personalized motor learning potential:*** The interaction between baseline levels and task-related modulation of GABA+ and Glx influenced short-term motor learning outcomes.

## 1 Introduction

Motor learning, the process of acquiring and refining motor skills through practice and experience, is vital to efficiently engage with the environment, serving as an indispensable ability in both health and disease. Especially bimanual movements play an important role in everyday life, since most activities needed for functional independence, such as getting dressed, eating with knife and fork, or driving a car require the ability to coordinate movements of both hands/arms. However, complex bimanual skills are not merely the summation of independently moving both limbs, but require practice to overcome basic inherent movement constraints (such as mirroring movements) to progress towards a more dexterous interaction of both hands (Swinnen, 2002). This complex process of motor learning is often divided into two distinct phases, commonly classified as (a) early or fast learning characterized by rapid performance gains typically observed within the first practice session, and (b) late or slow learning, marked by gradual skill development and consolidation of performance occurring after several hours or even weeks and months of practice. Late learning often necessitates extended practice sessions and consolidation periods (Dayan & Cohen, 2011; Karni et al., 1998). Despite the central role of motor learning in both health and disease, the neurochemical mechanisms are yet to be explored, potentially informing future rehabilitation strategies for movement disorders.

Although the exact mechanisms underlying practice are debated, it is widely accepted that motor learning is driven by processes of synaptic plasticity. These processes are called long-term potentiation (LTP) or long-term depression (LTD)-like plasticity, i.e., the strengthening or weakening of a connection between two neurons due to higher or lower frequencies of stimulation, respectively (Castillo et al., 2011; Rosenkranz et al., 2007; Sanes & Donoghue, 2000; Ziemann et al., 2004). Ultimately, these lead to long-term changes in neural circuits, including structural remodeling and synaptic reorganization, which contribute to the consolidation and retention of motor skills over extended periods (Kleim et al., 2004). The neurotransmitters gamma-aminobutyric acid (GABA) and glutamate (Glu) play key roles in modulating synaptic transmission and plasticity within the motor cortex. GABA and Glu are, respectively, the most abundant inhibitory (Watanabe et al., 2002) and excitatory (Zhou & Danbolt, 2014) neurotransmitter in the mammalian central nervous system. Together, GABA and Glu regulate the excitability of neuronal circuits by maintaining an excitatory/inhibitory balance (E-I balance), are involved in learning processes and synaptic plasticity, and are consequently crucial for motor control and learning (Johnstone et al., 2021; Jongkees et al., 2017; McDonnell et al., 2007). Alterations in GABAergic and glutamatergic neurotransmission are also linked to motor deficits and rehabilitation of neurological disorders such as stroke (Blicher et al., 2015; Chen et al., 2020; Grigoras & Stagg, 2021), highlighting their significance in shaping motor performance and adaptation.

Magnetic resonance spectroscopy (MRS) offers a non-invasive approach to quantify neurometabolites in vivo in a pre-defined volume of interest (VOI) in the brain (Buonocore & Maddock, 2015), providing valuable insights into the neurochemical correlates of motor learning. For the quantification of GABA, a relatively low-concentration metabolite that overlaps with other high-concentration metabolites such as creatine, specialized scanning sequences such as the Mescher-Garwood Point Resolved Spectroscopy (MEGA-PRESS) use spectral editing to aid the quantification of GABA at 3T (Mescher et al., 1998; Mescher et al., 1996; Mullins et al., 2014). In the context of motor learning, resting-state MRS enables the measurement of baseline GABA and Glx (a compound measure of Glu and glutamine) levels, reflecting the steady-state neurochemical environment in the motor cortex or other regions. Task-related or functional MRS (fMRS), on the other hand, allows for the real-time assessment of neurotransmitter dynamics during task performance, capturing transient changes in neurotransmitter concentrations associated with motor task execution and learning processes (Pasanta et al., 2022). Hence, MRS can be used to quantify levels of neurometabolites in the context of motor learning to either observe changes within one session of motor learning (i.e., short-term effects in the context of fast learning), or across different days of motor learning (i.e., long-term effects that are associated with slow learning).

Several brain regions are crucial for bimanual motor control and learning, and particularly the primary sensorimotor cortex (SM1) and dorsal premotor cortex (PMd) are of main interest. SM1, comprising the primary motor cortex (M1) and primary somatosensory cortex (S1), is essential to motor learning (e.g., Huang et al., 2024; Ogawa et al., 2019; Sanes & Donoghue, 2000). In M1, voluntary movements are initiated and controlled (Pearson, 2000), while in S1 sensory input from the periphery is processed (Borich et al., 2015). PMd is densely connected to M1, the sensory cortex and associational areas (Genon et al., 2018), and plays a key role in integrating sensory information and modulating M1 during motor tasks (Chouinard & Paus, 2006; Dum & Strick, 1991), particularly during complex bimanual coordination (Beets et al., 2015; Debaere et al., 2004; Karabanov et al., 2023; Van Ruitenbeek et al., 2023; Verstraelen et al., 2021). The left PMd is especially significant in bimanual motor control, as it encodes movements of both upper limbs and is heavily involved in movement planning and skill acquisition (Debaere et al., 2004; Fujiyama et al., 2016; Hardwick et al., 2013; Merrick et al., 2022).

To date, studies investigating motor-related changes in neurotransmitter levels have mainly focused on SM1 but not PMd. A meta-analysis of functional MRS studies reported task execution to result in increased Glu/Glx levels across task domains and brain regions, and potential changes in GABA levels specific to the motor domain (Pasanta et al., 2022). For SM1, an increase in Glu/Glx during motor task execution such as finger tapping (Schaller et al., 2014), rhythmic hand clenching (Chen et al., 2017; Volovyk & Tal, 2020) and 30 minutes after reactivation of a learned finger tapping sequence (Eisenstein et al., 2023) have been reported. Furthermore, higher Glu levels at rest and at the start of executing a motor task have been associated with better learning of a serial reaction time task (SRTT) (Bell et al., 2023). In contrast, no change in Glu levels during motor learning (Bell et al., 2023; Floyer-Lea et al., 2006; Kolasinski et al., 2019; Maruyama et al., 2021), during the performance of a bimanual interference task (Rasooli et al., 2024) or after initial motor learning of an SRTT (Eisenstein et al., 2024) have been reported.

For GABA levels in SM1, a decrease in GABA levels (as measured during task performance) after a few minutes of continuous rhythmic hand clenching (Chen et al., 2017) and during motor learning of a force production motor sequence task (Floyer-Lea et al., 2006) or an SRTT (Kolasinski et al., 2019) have been reported. In constrast, some studies reported no change in GABA levels while learning an SRTT (Bell et al., 2023; Maruyama et al., 2021), while being exposed to a bimanual interference task (Rasooli et al., 2024), during finger tapping (Schaller et al., 2014) or after learning an SRTT (Eisenstein et al., 2023, 2024) or a bimanual coordination task (Chalavi et al., 2018). Moreover, baseline GABA levels in SM1 have also been linked to motor learning. One study reported that lower baseline GABA levels in SM1 predicted a better initial performance on the BTT, but that neither GABA nor Glx levels were linked to the learning progress over a three-day motor training (Chalavi et al., 2018). Similarly, lower baseline GABA levels in M1 were associated with better subsequent motor learning on a sequential reaction time task (Kolasinski et al., 2019). Another study investigated the association between baseline neurotransmitter levels in S1 and M1 and a five-day bimanual motor training using the BTT and found a positive association between baseline GABA levels and initial learning progress. More specifically, higher baseline levels in S1 or M1 showed an association with better learning when respectively augmented visual feedback was or was not provided (Li et al., 2024). However, later learning progress did not show an association with neurotransmitter levels at baseline (Li et al., 2024). Interestingly, one study in which anodal transcranial direct current stimulation (tDCS) was employed showed that the ability to modulate (i.e., decrease) GABA levels in M1 was associated with better motor learning on an SRTT (Stagg et al., 2011).

Although several studies have investigated the immediate effects of motor learning on Glu/Glx and GABA levels in SM1, there are no reports of neurotransmitter levels in PMd in contexts such as motor learning or working memory. Only one study has looked into long-term changes in neurotransmitter levels in SM1, revealing a decrease in GABA levels at rest after six weeks of low intensity (i.e., five days per week for 15 min), but not high intensity (30 min), juggling training (Sampaio-Baptista et al., 2015).

Since evidence suggests that LTP- and LTD-like neuroplasticity processes are mediated by changes in GABA (Kim et al., 2014) and/or Glu (Lüscher & Malenka, 2012) neurotransmitter levels, understanding the neurochemical changes within these cortical regions during bimanual motor learning provides valuable insights into the neural mechanisms underlying the acquisition and refinement of complex motor skills. Changes in the E-I balance might lead to an increased or decreased communication between neurons (Butefisch et al., 2000; Ziemann et al., 2001), which may ultimately facilitate synaptogenesis (Kleim et al., 2002; Kleim et al., 2004) and synaptic pruning (Wenger et al., 2017; Yang et al., 2009). This process of synaptic reorganization is vital for motor learning.

In this study, we aimed to explore the temporal dynamics of GABA and Glx changes within the SM1 and PMd during bimanual motor learning using a longitudinal MRS approach. By investigating both resting (bilateral SM1, left PMd) and functional (left PMd) neurotransmitter profiles, we unraveled the excitatory and inhibitory neurotransmitter dynamics underlying skill acquisition and consolidation. We hypothesized that the resting levels of neurotransmitters would change in the context of a four-week motor learning intervention and that the task-related modulation of excitatory and inhibitory neurotransmitters in left PMd would be more pronounced in the initial as compared to the final stage of the motor training, reflecting the need for more modulation of the motor output via the PMd when task proficiency is still low. Finally, we expected these neurotransmitter correlates to be linked to the short- and long-term performance gains.

## 2 Materials & Methods

### 2.1 Participants

Sixty-two volunteers (aged 25.94 ± 4.02 [mean ± SD] years, range 19–34; n = 37 [59.7%] female) participated in this MRS experiment: initial inclusion: n = 68; exclusion due to brain anomalies on initial structural magnetic resonance imaging [MRI] scan: n = 3; exclusion due to non-adherence to training protocol: n = 2; no MRS data due to technical difficulties: n = 1. Participants were right-handed according to the Edinburgh Handedness Inventory laterality quotient (EHI LQ; 86.61 ± 13.45%, range 50–100) (Oldfield, 1971) and showed no indication of cognitive impairments as assessed with the Montreal Cognitive Assessment (MoCA; 28.39 ± 1.50 points, range 25–30) (Rossetti et al., 2011). The study protocol was approved by the local ethics committee (Ethics Committee Research UZ/KU Leuven; reference S65077) and participants gave full written informed consent prior to study initiation, according to the latest amendment of the Declaration of Helsinki (World Medical Association, 2013).

### 2.2 Overview of the experiment

A four-week training on the BTT was conducted with three comprehensive measurement sessions at baseline (PRE), after two weeks (MID), and after four weeks of motor training (POST). During a screening session, participant’s safety and eligibility to participate in this research was checked. Further, participants were presented with a brief familiarization on the bimanual tracking task (BTT) (15 trials of the simplest 1:1 frequency ratio task, ∼3 min), while lying supine in a mock MRI scanner. Then a pre-test of the BTT was performed, and participants were pseudo-randomly (stratified by sex) assigned to either a complex or a simple training group of the BTT (task and training details, see below). During the PRE and POST measurement, a SynVesT1 positron emission tomography (PET) scan to assess synaptic density (subsample of n = 22; scan of 30 min), an MRI protocol (∼2 h), and a dual-site transcranial magnetic stimulation (TMS) protocol to assess the intra- and interhemispheric PMd–M1 connectivity (∼4 h) were conducted. At the MID measurement, only the MRI protocol was assessed. A single MRI session (conducted at PRE, MID and POST session) consisted of the following scans (in order): resting-state fMRI (∼15 min), high-resolution T1- and T2-weighted anatomical images (∼10 min), resting MRS of right and left SM1 (∼22 min), break outside of scanner (5 min), low-resolution T1-weighted anatomical scan (∼2 min), resting and task-related MRS of left PMd (∼45 min), diffusion-weighted imaging (∼10 min). All MRI sessions followed the exact same protocol and order. Importantly, none of the used measurement methods is of an interventional nature, meaning that the PET, MRI or TMS measurements themselves have no influence on the participant’s skill acquisition or brain characteristics (as opposed to, e.g., certain repetitive TMS protocols). In the present research, we will only describe the MRS data (and the T1-weighted anatomical data in function of it) of this research (for an overview of the experimental design regarding the MRS acquisition, see **Figure 1**).

**Figure 1.**
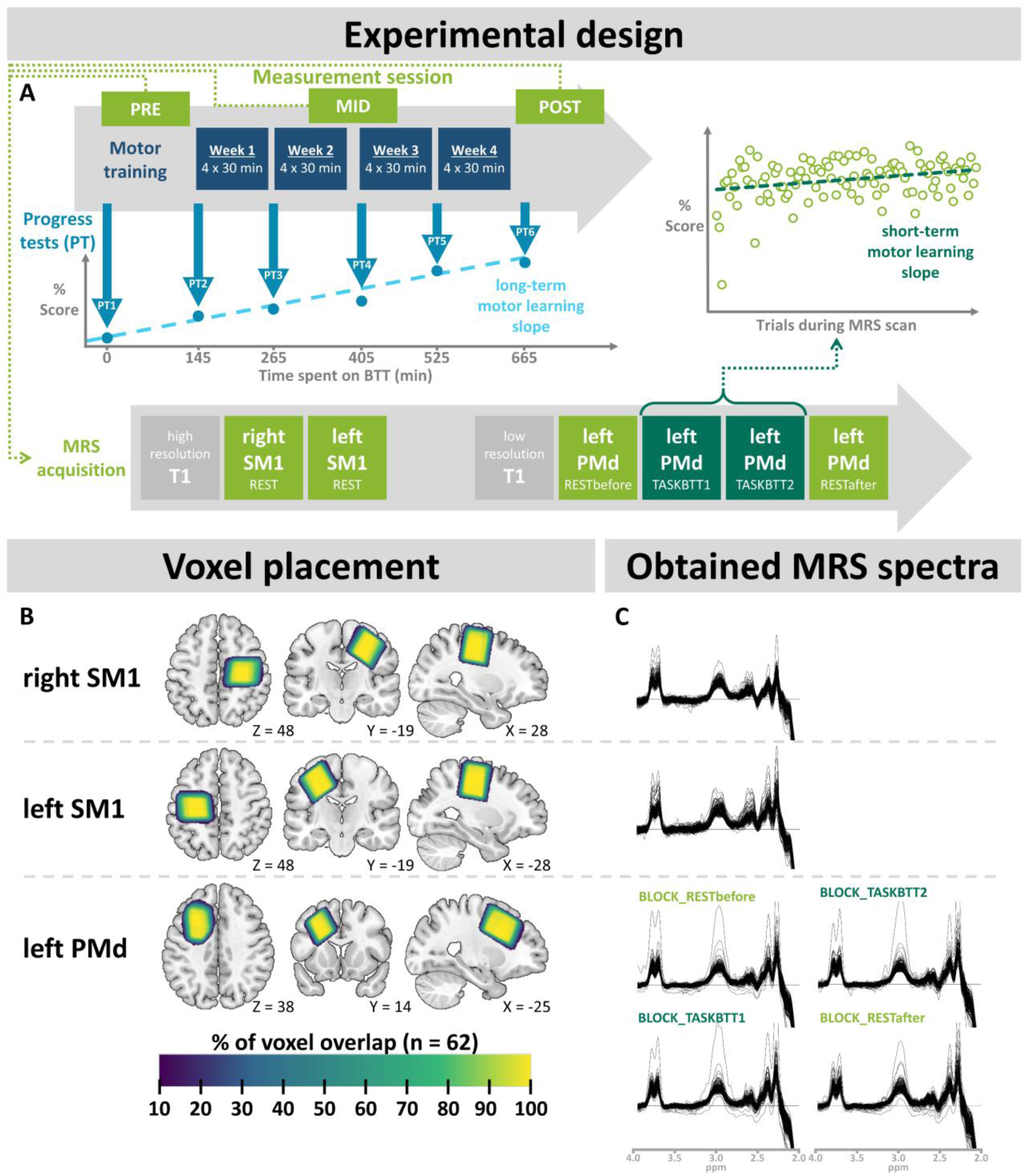
Overview experimental design. **[A]** Timing of the three measurements at baseline (PRE session), after two weeks (MID session), and after four weeks of motor training (POST session) on a bimanual tracking task (BTT). During each magnetic resonance (MR) measurement session, neurometabolite levels in the right and left primary sensorimotor cortex (SM1) were acquired at rest (each ∼8 min acquisition time), and for the left dorsal premotor cortex (PMd) both resting and task-related MRS were collected in four blocks (∼11 min each): one at rest (BLOCK_RESTbefore), two during the BTT (BLOCK_TASKBTT1, BTT2), and one at rest after the task (BLOCK_RESTafter). Short-term motor learning progress was defined by the linear slope of BTT scores within a session (right plot), while long-term progress was defined by the linear slope of the standardized progress test (PT) scores assessed at six time points over the four-week training period (left plot). **[B]** Voxel placement (warped to MNI space) during PRE session, serving as the reference for placement during the MID and POST session, and percentage overlap between participants. **[C]** Obtained MR spectroscopy (MRS) spectra from all three measurement sessions per region and task-block.

### 2.3 Bimanual motor training and behavioral measures

#### Motor training intervention

All participants followed a motor learning paradigm that lasted for four weeks, consisting of four 30-min (6 × 5 min, with short self-paced breaks after each 5-min block) training sessions per week. To induce training-related plasticity, the BTT (Fujiyama et al., 2016; Sisti et al., 2011; Zivari Adab et al., 2020) was selected. This is a novel task that does not belong to the daily motor repertoire but involves similar neural mechanisms like those required for everyday bimanual movements. Besides, the task allows an easy adaptation of the difficulty level based on the individual performance level to assure optimal challenge and motivation during motor learning.

During the BTT training, participants were seated in an upright position facing a 15-inch laptop and the task setup positioned in front of them (see **Figure 2A**). Participants were instructed to accurately track a visual target (white dot) presented on the screen. Two seconds after the beginning of each trial, the target started to move at a constant speed over the target line (blue line). Participants were asked to track the target as accurately as possible by rotating two dials with the left and right index finger to control respectively the vertical and horizontal movements of a cursor, resulting in a red path (reflecting the actual trajectory covered) (see **Figure 2B**) (Fujiyama et al., 2016; Sisti et al., 2011; Zivari Adab et al., 2020). After each trial, participants received brief on-screen visual feedback about their performance, expressed as a percentage ranging from 0–100%. The score reflected the overall accuracy considering speed, movement direction, and distance from the target (see **Appendix 1** for all BTT details and the score calculation).

**Figure 2.**
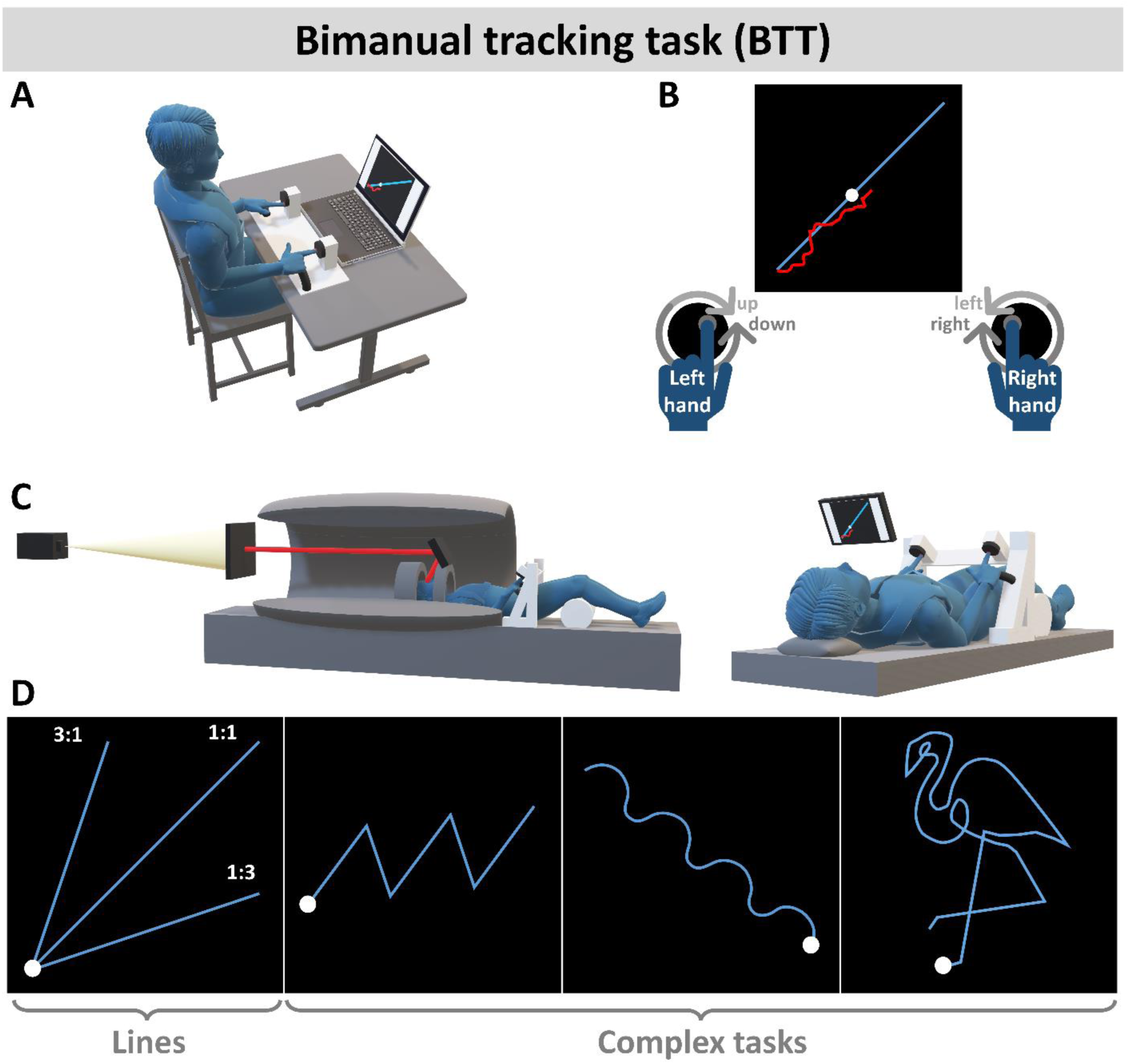
Overview of the bimanual tracking task (BTT). **[A]** BTT training setup for home training and progress tests. Participants were comfortably seated, the task setup and a laptop placed in front of them, with their hands resting on the handles of the task setup and index fingers in the grooves of the BTT dials. [**B**] The main goal of the BTT was to trace a white dot, moving at a constant speed along a blue target line, as accurately as possible. To do so, participants were instructed to rotate the left and right dial with their index fingers in order to respectively manipulate the vertical and horizontal movements of their own trajectory, indicated by a red line. **[C]** Magnetic resonance (MR) compatible BTT setup for task execution during MR spectroscopy. Participants were in supine position, elbows well-supported, hands resting on the BTT handles and index fingers in the grooves of the BTT dials. Visual task input was provided via a projector and a screen that participants could view via a mirror placed in front of their eyes (see mirror path displayed in red). **[D]** Standardized progress tests to evaluate each individual’s skill level throughout task practice. The set of easier straight-line tasks (frequency ratios of the left:right hand are indicated purely for illustrative purposes) and complex tasks (zig-zag, waves, flamingo) were assessed six times in total: at baseline (PRE), at the beginning of each of the four training weeks, and after finalizing the training paradigm (POST). The lines were presented four times per frequency ratio (once per quadrant of movement direction, to cover each movement speed and direction for each hand), the complex tasks were presented twice (original and vertically mirrored).

After successful screening, participants were pseudo-randomly assigned to a complex or simple training group, stratified by sex. In both training groups, participants performed the BTT four times per week for 30 minutes, with short self-paced breaks every 5 minutes. In the complex training group, maximal motor learning was pursued by continuously adjusting the task difficulty based on the individual performance level, using an adapting staircase-like level paradigm. More specifically, this training consisted of 280 different trials, combined and arranged to 5-minute levels starting with easier levels (i.e., consisting of straight-line trials) and progressing to more difficult levels (i.e., consisting of curves, zig-zags, waves and complex figures and various combinations including variations in speed). When performing well on a level (i.e., reaching on average > 65%), then participants would progress to the next level, whereas medium performance (40¬–65%) or bad performance (< 40%) would lead to stagnation or regression in levels (see **Appendix 1** for details). In contrast, the simple training group spent the same amount of time on the same task setup but only trained the simplest variant (1:1 frequency ratio line requiring both hands have to move equally fast, see **Figure 2D**, first panel), limiting the complexity level and thus the amount of motor learning as compared to the complex group. To compare motor learning independently of the training condition (complex vs. simple) and individual progress on the level scheme, participants of both training groups were presented with a progress test (PT). This test was administered six times (at baseline, at the beginning of each training week, and after finalizing the training paradigm) and consisted of a standardized set of tasks (see **Figure 2D**, and mirrored counterparts, total of 18 tasks, duration: ∼5 min). Furthermore, the performance (Score in %) on the progress tests was used to quantify the learning progress in both groups. Specifically, the linear slope of the progress test scores over time was estimated by a linear regression using the average scores of each of the progress tests as dependent variable and the time spent on the BTT (BTT_time_; summed over all training and measurement sessions, expressed in minutes) as the independent variable (see **Figure 1A**). To obtain a detailed picture of the long-term learning progress over the four-week training period, linear slopes were calculated for the average scores on the BTT progress tests (PT_BTT) of: (1) all straight line trials, i.e., 1:1, 1:3 and 3:1 (left:right hand) frequency ratios (PT_BTTslope_Lines_ derived from PT_BTTscore_Lines_), and (2) the three complex task variants, i.e., zig-zag, waves, and flamingo trials (PT_BTTslope_Complex_ derived from PT_BTTscore_Complex_) (see **Figure 2D**). These two groups of subtasks were analyzed separately since they reflect the skill level and motor learning progress on simpler and more complex tasks, respectively.

#### BTT during MRS scans

During the task-related MRS acquisition of the left PMd, participants performed the BTT inside of the MR scanner (see section “Magnetic resonance (MR) acquisition” below; duration: 20 min, about 100 trials in total). To do so, an adapted non-ferromagnetic version of the BTT setup was used, which could be placed over the participants’ hips in a bridge-wise manner and could effortlessly be operated in supine position (see **Figure 2C**). The visual input of the BTT was presented to the participant via a projection at the cranial end of the MR scanner and a mirror (∼14 × 9 cm) placed in front of (∼13 cm distance) the participant’s eyes (LCD projector: NEC NP-PA500U, 1920 × 1200 pixels; see red line in **Figure 2C**, representing the path of the visual input). During the MRS sessions, participants performed only straight-line tasks with the frequency ratios 1:1, 1:3 and 3:1 (left:right hand movements), with the target moving in the four possible directions (towards upper right/ upper left/ lower right/ lower left corner of the screen). Linear slopes for the progress made during the training session within the MR scanner were calculated with trial number as independent variable and score as dependent variable. Since only straight-line trials were performed in the scanner (i.e., 1:1, 1:3 and 3:1 frequency ratios), slopes were calculated for all tasks (MR_BTTslope_allLines_) to quantify short-term learning progress during the scanning session, representing performance progression as a result of learning (see **Figure 1A**, right panel). For a more detailed behavioral analysis, the motor learning progress slopes for 1:1 frequency ratio trials (MR_BTTslope_11_) and 1:3/3:1 frequency ratio trials together (MR_BTTslope_13,31_) were calculated as well.

### 2.4 Magnetic resonance (MR) acquisition

Magnetic resonance imaging and spectroscopy data were acquired using a 3 Tesla Philips Achieva dstream scanner (University Hospital Leuven, Gasthuisberg) with a 32-channel receiver head coil (Philips, Best, The Netherlands). A T1-weighted high-resolution anatomical image was collected using a three-dimensional turbo field echo (3DTFE) sequence (echo time [TE] = 4.6 ms, repetition time [TR] = 9.7 ms, flip angle = 8°, field of view = 256 × 242 × 182 mm, 182 sagittal slices, voxel size = 1 × 1 × 1 mm^3^; acquisition time ∼6 min), based on which the MRS VOIs for the right and left SM1 were placed. Since there was a brief break in the middle of the MRI session including participant removal from and replacement in the scanner bore (see section “Overview of the experiment”), an additional lower-resolution T1-weighted 3DTFE (TE = 4.6 ms, TR = 9.6 ms, flip angle = 8°, field of view = 256 × 244 × 182 mm, 182 sagittal slices, voxel size = 1.5 × 1.5 × 1.5 mm^3^; acquisition time ∼2 min) was acquired before MRS acquisition in the left PMd to inform VOI placement. During processing of the MRS data, high-resolution T1-weighted images were used for all voxels (see section “MRS data processing” below).

MRS spectra were acquired using a MEGA-PRESS sequence (Edden & Barker, 2007; Mescher et al., 1998) (TE = 68 ms, TR = 2000 ms, samples = 1024, spectral bandwidth = 2kHz), with ON and OFF spectra being collected in an interleaved fashion and editing pulses (basing pulse duration = 14 ms) being applied at 1.9 or 7.46 ppm, respectively. An automatic first-order pencil-beam (PB) shimming procedure was performed, and Multiply Optimized Insensitive Suppression Train (MOIST; bandwidth 140 Hz) water suppression was used. For each MRS acquisition, an additional 16 unsuppressed water averages were acquired and used for online interleaved water referencing. The edited signal detected at 3 ppm contains co-edited contributions from both macromolecules (MM) and homocarnosine, and hence it will be referred to as GABA+ rather than GABA.

During the first session, MRS VOIs were placed based on anatomical landmarks, individually defined on the T1-weighted MRI (see **Figure 1B** for VOI visualization and **C** for obtained spectra). For follow-up scans, views of each VOI in all three planes and superimposed on the T1-weighted image acquired at the first session were used to guide positioning of each voxel during the second and third session. MRS VOIs were placed in the right and left SM1 [voxel dimensions = 30 × 30 × 30 mm^3^; 112 edit-ON/112 edit-OFF spectra acquired; ∼8 min acquisition time (Mikkelsen et al., 2018)], with the center of the VOI placed over the respective hand knob (Yousry et al., 1997) in the axial view and alignment of the VOI with the cortical surface in the coronal and sagittal view. A third MRS VOI was placed in the left PMd (voxel dimensions = 40 × 25 × 25 mm^3^; 160 ON/160 OFF spectra acquired; ∼11 min acquisition time per acquisition block). The VOI was placed in the axial view just anterior to the left hand knob, with the posterior surface of the VOI aligned with the precentral sulcus, the medial surface parallel to the longitudinal fissure (Greenhouse et al., 2016; Maes et al., 2021), and the longitudinal center of the VOI over the superior frontal sulcus (Maes et al., 2020). The VOI was then aligned to the cortical surface in the sagittal and coronal view. For the left PMd, four directly adjacent acquisitions were conducted (total of ∼45 min) with the participant at rest during the first acquisition block (BLOCK_RESTbefore), performing the BTT inside of the MR scanner during the second and third acquisition block (BLOCK_BTT1 and BLOCK_BTT2, respectively), and resting again during the fourth block (BLOCK_RESTafter) (see **Figure 1A**).

### 2.5 MRS data processing

MRS data were analyzed using the MATLAB-based (vR2022a, MathWorks, Natick, MA) software toolkit ‘Gannet’ (version 3.3.1) (Edden et al., 2014), specifically designed to analyze edited single-voxel MRS data. GABA+ and Glx quantification were conducted according to the following steps: (1) frequency and phase alignment of free induction decays with spectral registration in the time domain (Near et al., 2015); (2) subtraction of aligned and averaged edit-ON from edit-OFF spectra to obtain GABA+-edited (3.0 ppm) and Glx (Glu + glutamine; 3.75 ppm) co-edited difference spectra; (3) fitting a three-Gaussian function using nonlinear least-squares fitting to quantify the GABA+ and Glx peak areas between 4.1 and 2.79 ppm, and a Gaussian-Lorentzian model to fit the water signal between 5.6 and 3.8 ppm which was used as reference (Mikkelsen et al., 2019); (4) water-scaling using the unsuppressed water reference signal and tissue-correction of GABA+ and Glx levels based on gray matter (GM), white matter (WM) and cerebrospinal fluid (CSF) fractions within each VOI. To obtain these, the three-dimensional T1-weighted high-resolution images were segmented using SPM 12 (Statistical Parametric Mapping v7771, Wellcome Trust Centre for Human Neuroimaging, University College, London, UK), and the individual MRS VOIs were coregistered to the structural image in Gannet (Edden et al., 2014). Since PMd VOIs were placed on a lower-resolution T1-weighed image, a co-registration of the high-resolution to the lower-resolution anatomical image was performed, and further analysis of the PMd MRS data was done based on the segmentation of that co-registered high-resolution T1-weighed image. Tissue correction was performed and included correction for CSF (i.e., assumption of negligible GABA in CSF) and α-correction (i.e., assumption of twice as much GABA in GM as in WM) (see (Harris et al., 2015), Equation 5).

Data quality of individual MRS spectra was assessed by visual inspection of the GannetLoad and GannetFit output (screening for lipid contamination, unsuccessful water suppression, overall spectral and fit quality) and quantitatively excluding data with high fit errors [cutoff fit error > 12% (Puts et al., 2018)] or low signal-to-noise ratio (SNR) of the GABA+ or Glx signal [cutoff SNR_GABA+_ < mean – 3SD (Maes et al., 2021)].

### 2.6 Statistical analysis

Behavioral and MRS data were analyzed using R Studio (R version 4.2.2, RStudio 2022.12.0 Build 353; α set to 0.05 unless specified otherwise). If necessary, to comply with model assumptions, a transformation of the dependent variable was performed. Models were manually simplified according to the stepwise backwards procedure, where non-significant effects (based on type III analysis of variance [ANOVA] fixed effect test, p > 0.05) were removed from the model one-by-one, keeping main effects that were still included in an interaction in the model. This procedure is described in detail in the appendix for significant results. Where applicable, post hoc t-tests with Bonferroni correction were applied.

#### Behavioral data

To investigate progress in motor skill over the four-week training period, linear mixed models (LMM) were used with BTT scores of the progress tests as dependent variable (one LMM for each, simple and complex subtasks respectively: PT_BTTscore_Lines_ and PT_BTTscore_Complex_), SUBJECT as random effect, and GROUP (simple vs. complex training group), and BTT_time_ (in min) as independent variables. Furthermore, the interaction GROUP×BTT_time_ was tested: PT_BTTscore ∼ GROUP + BTT_time_ + GROUP×BTT_time_ + (1|SubjectID), with two levels for GROUP (simple vs. complex training group) and BTT_time_ being a continuous variable. Since BTT_time_ was a continuous variable, classic post-hoc testing was not possible. Nevertheless, the exact timing of training progress and differences between groups can be informative to understand changes in neurometabolites. Hence, t-tests/Wilcoxon rank sum tests with Bonferroni correction were used to compare scores on consecutive progress tests and, where applicable, the effect of the training group on the performance (PT_BTTscore_Lines_ and PT_BTTscore_Complex_). Lastly, for the learning slopes as measures of learning progress (PT_BTTslope_Lines_ and PT_BTTslope_Complex_), a group comparison (simple vs. complex training group) was performed using independent Welch t-tests to assess differences in learning speed.

For in-scanner learning on the BTT, the learning slopes during the MRS scans (MR_BTTslope_allLines_, MR_BTTslope_11_, MR_BTTslope_13,31_) were compared per training group between measurements (PRE, MID, POST) using paired t-tests, and per measurement between training groups (complex vs. simple training) using independent Welch t-tests. Only the compound measure MR_BTTslope_allLines_ was used for analyzing associations between short-term learning and neurometabolite levels. Additionally, the 1:1 and 1:3/3:1 frequency ratio learning slopes were behaviorally analyzed in order to investigate whether there were training group effects for each of the subtask groups.

#### Baseline neurometabolite levels to predict motor learning progress

The predictive value of GABA+ and Glx levels at rest during the PRE measurement were assessed using six different multiple linear regressions. Each motor learning outcome (MR_BTTslope_allLines_ of the PRE measurement, PT_BTTslope_Lines_, and PT_BTTslope_Complex_ respectively for motor learning progress in the short-term [i.e., within the first MRS session], in the long-term [i.e., over the four-week motor learning intervention] on simple subtasks, and in the long-term on complex subtasks) was regressed by the neurometabolite (NM) levels in the three VOIs (left SM1, right SM1, left PMd), resulting in two linear models per motor learning outcome: Short-term motor learning outcome ∼ NM_L-SM1 + NM_R-SM1 + NM_L-PMd, with NM = GABA+ for linear model 1, and NM = Glx for linear model 2. For short-term motor learning (where only PRE measurement data was included), the GROUP assignment was not of interest for the model since a group specific intervention was only applied afterwards. For the long-term motor learning outcome measures concerning the whole training phase, GROUP was additionally added to the model, as well as the interaction effects between GROUP and the neurometabolite level in each VOI: Long-term motor learning outcome ∼ NM_L-SM1 + NM_R-SM1 + NM_L-PMd + GROUP + NM_L-SM1×GROUP + NM_R-SM1×GROUP + NM_L-PMd×GROUP, with two levels for GROUP (simple vs. complex training group) and the NM levels being continuous variables.

#### Changes in resting neurometabolite levels as a result of motor learning

To assess changes in resting GABA+ and Glx levels per VOI associated with motor learning, six LMMs (one per neurometabolite [GABA+, Glx] and per VOI [left SM1, right SM1, left PMd]) were constructed according to the formula: NM_VOI ∼ GROUP + SESSION + GROUP×SESSION + (1|SubjectID), with two levels for GROUP (simple vs. complex training group) and three levels for SESSION (PRE vs. MID vs. POST).

#### Modulation of left PMd neurometabolites during task execution and changes in modulation with motor learning

The modulation of left PMd GABA+ and Glx levels was investigated for the PRE measurement using an LMM with the neurometabolite levels in left PMd during the PRE measurement (GABA+ or Glx) as dependent variable, GROUP with two levels (simple vs. complex training group), TASK with four levels (11-minute blocks of MRS acquisition: BLOCK_RESTbefore, BLOCK_BTT1, BLOCK_BTT2, BLOCK_RESTafter), and their interaction as fixed effects of interest, the linewidth of the water signal as covariate of no interest [since changes in BOLD have previously been associated with changes in linewidth at 7T (Stanley & Raz, 2018), which despite lacking evidence might influence the GABA+ and Glx quantifications at 3T (Dwyer et al., 2021)], and SubjectID as random effect: NM_L-PMd ∼ GROUP + TASK + GROUP×TASK + Water_FWHM + (1|SubjectID). Changes in neurometabolite modulation associated with the four-week motor training intervention were assessed with LMMs as stated above but including data of all three measurements and SESSION respective interactions up to the second degree as fixed effects: NM_L-PMd ∼ GROUP + SESSION + TASK + GROUP×SESSION + SESSION×TASK + GROUP×TASK + Water_FWHM + (1|SubjectID).

#### Relationship between modulation of left PMd neurometabolites and motor learning

To investigate whether the modulation of neurometabolites was related to the short-term or long-term motor learning progress, an additional measure of the maximal modulation per subject and session was calculated. The absolute maximal modulation (|MODmax|) was calculated as |MODmax| = max(|NM[BLOCK_BTT] - NM[BLOCK_RESTbefore]|). In other words, the neurometabolite level at baseline (BLOCK_RESTbefore) was subtracted from the task-related block with the maximal change in neurometabolite (NM[BLOCK_BTT], corresponding to either NM[BLOCK_BTT1] or NM[BLOCK_BTT2]). In addition, a directional measure of |MODmax| was used (MODmax), where decreases in neurometabolite levels from baseline to task were indicated by a negative sign, and increases by a positive sign (i.e., MODmax = NM[BLOCK_BTT] - NM[BLOCK_RESTbefore]). Hence, to summarize, the absolute maximal modulation |MODmax| focuses on the magnitude of the modulation, ignoring the direction, whereas the directional maximal modulation MODmax additionally incorporated the direction of this change in neurometabolite levels.

To evaluate the influence of neurometabolite modulation during the first session on short- and long-term motor learning progress, multiple linear regressions were used to model each motor learning outcome (MR_BTTslope_allLines_ of the PRE measurement, PT_BTTslope_Lines_, and PT_BTTslope_Complex_ respectively for overall short-term, simple long-term and complex long-term motor learning progress) by the GABA+ and Glx modulation in left PMd during the PRE measurement: Motor learning outcome ∼ GABA_MODmax + Glx_MODmax + GABA_MODmax×Glx_MODmax. The same was done for |MODmax| instead of MODmax to investigate the influence of the strength of modulation on motor learning, resulting in a total of six multiple linear regressions (3 motor learning outcomes × 2 modulation measures).

## 3 Results

### 3.1 MRS data quality and VOI placement

#### MRS data quality

Of the 1140 acquired spectra, two (right and left SM1 of the same participant from PRE measurement) were excluded based on low SNR of the GABA+ signal (outlier: SNR < mean–3*SD). No further data points were excluded. An overview of the data quality can be found in **Table 1**, displaying the linewidth of GABA+, Glx, Water, and creatine in full-width half maximum (FWHM) as a measure of spectral resolution and the degree of signal overlap, the SNR for the metabolites of interest (GABA+, Glx), the FitError for the GABA+:Water and Glx:Water quantification as fitted by the Gannet toolbox, and the voxel fractions of GM, WM, and CSF in each VOI.

**Table 1.**
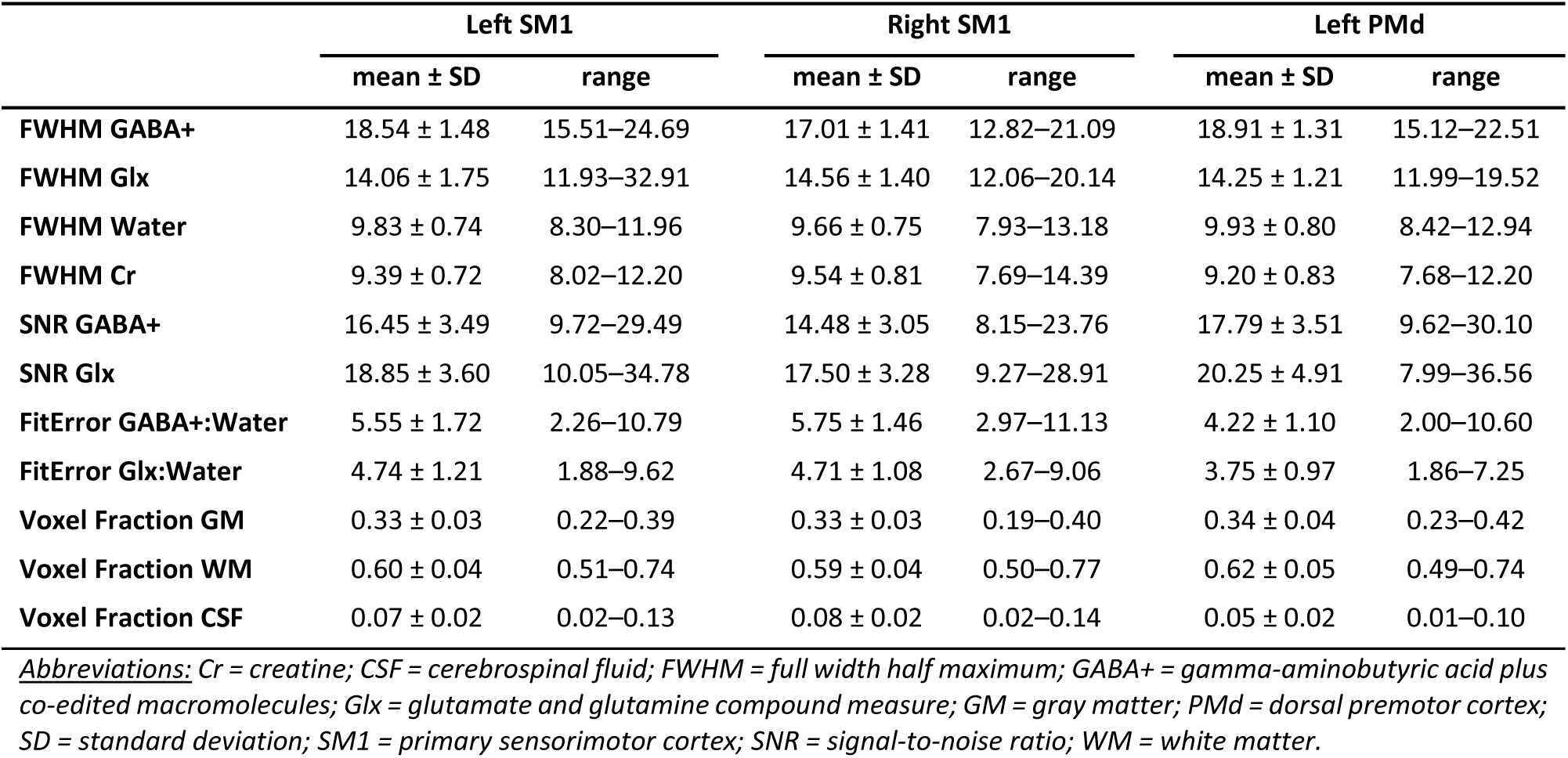
Magnetic resonance spectroscopy (MRS) data quality measures for left primary sensorimotor cortex (SM1), right SM1, and left dorsal premotor cortex (PMd). Data are pooled over the three measurement sessions (PRE, MID, and POST).

|  | Left SM1 |  | Right SM1 |  | Left PMd |  |
| --- | --- | --- | --- | --- | --- | --- |
| | mean ± SD | range | mean ± SD | range | mean ± SD | range |
| **FWHM GABA+** | 18.54 ± 1.48 | 15.51–24.69 | 17.01 ± 1.41 | 12.82–21.09 | 18.91 ± 1.31 | 15.12–22.51 |
| **FWHM Glx** | 14.06 ± 1.75 | 11.93–32.91 | 14.56 ± 1.40 | 12.06–20.14 | 14.25 ± 1.21 | 11.99–19.52 |
| **FWHM Water** | 9.83 ± 0.74 | 8.30–11.96 | 9.66 ± 0.75 | 7.93–13.18 | 9.93 ± 0.80 | 8.42–12.94 |
| **FWHM Cr** | 9.39 ± 0.72 | 8.02–12.20 | 9.54 ± 0.81 | 7.69–14.39 | 9.20 ± 0.83 | 7.68–12.20 |
| **SNR GABA+** | 16.45 ± 3.49 | 9.72–29.49 | 14.48 ± 3.05 | 8.15–23.76 | 17.79 ± 3.51 | 9.62–30.10 |
| **SNR Glx** | 18.85 ± 3.60 | 10.05–34.78 | 17.50 ± 3.28 | 9.27–28.91 | 20.25 ± 4.91 | 7.99–36.56 |
| **FitError GABA+:Water** | 5.55 ± 1.72 | 2.26–10.79 | 5.75 ± 1.46 | 2.97–11.13 | 4.22 ± 1.10 | 2.00–10.60 |
| **FitError Glx:Water** | 4.74 ± 1.21 | 1.88–9.62 | 4.71 ± 1.08 | 2.67–9.06 | 3.75 ± 0.97 | 1.86–7.25 |
| **Voxel Fraction GM** | 0.33 ± 0.03 | 0.22–0.39 | 0.33 ± 0.03 | 0.19–0.40 | 0.34 ± 0.04 | 0.23–0.42 |
| **Voxel Fraction WM** | 0.60 ± 0.04 | 0.51–0.74 | 0.59 ± 0.04 | 0.50–0.77 | 0.62 ± 0.05 | 0.49–0.74 |
| **Voxel Fraction CSF** | 0.07 ± 0.02 | 0.02–0.13 | 0.08 ± 0.02 | 0.02–0.14 | 0.05 ± 0.02 | 0.01–0.10 |

**Abbreviations:** Cr = creatine; CSF = cerebrospinal fluid; FWHM = full width half maximum; GABA+ = gamma-aminobutyric acid plus co-edited macromolecules; Glx = glutamate and glutamine compound measure; GM = gray matter; PMd = dorsal premotor cortex; SD = standard deviation; SM1 = primary sensorimotor cortex; SNR = signal-to-noise ratio; WM = white matter.

#### Within-participant VOI overlap between sessions

To evaluate the accuracy of VOI placement, the overlap between sessions was calculated. Within participants, VOI overlap between sessions was consistently high. Average VOI overlap was 91.99 ± 6.27% between PRE and MID measurement, and 92.69 ± 4.52% between PRE and POST measurement (left SM1: 92.21 ± 7.81% and 93.36 ± 3.98%; right SM1: 92.14 ± 6.50% and 93.04 ± 4.00%; left PMd: 91.63 ± 3.99% and 91.67 ± 5.34% for PRE vs. MID and PRE vs. POST, respectively). The part of the VOIs that overlapped between all three measurement sessions was 87.55 ± 7.17% (left SM1: 88.05 ± 8.28%, right SM1: 87.82 ± 7.49%, left PMd: 86.79 ± 5.51%).

#### VOI overlap between left SM1 and PMd

On average, the left PMd VOI overlapped with 6.15 ± 5.71% (i.e., 1.66 ± 1.54 ml) of the left SM1 VOI, indicating that overlap was minimal and that the two VOIs carry primarily different information despite their adjacent location.

### 3.2 Behavioral outcomes of motor learning paradigm

Training adherence was high, with an overall 99.75 ± 0.79% of the 5-min training blocks being completed. There was no significant difference between training groups (complex group: n = 32, 99.90 ± 0.42%; simple group: n = 30, 99.75 ± 0.79%; independent t-test: t(60) = 0.9493, p = 0.3463).

#### Performance improvements but no group difference for simple task variants with motor training

Participants’ performance on the progress tests improved significantly when more time was spent on the BTT for both, simpler and more complex task variants, with a difference between the performance and motor learning progress of the two training groups only being present for the complex task variants. Furthermore, for the simpler straight-line tasks of the progress tests (PT_BTTscore_Lines_), cubic transformation of the dependent variable was necessary to comply with the assumption of normally distributed residuals. In a stepwise backwards procedure, first the GROUP×BTT_time_ interaction (p = 0.6715) and then the main effect of GROUP (p = 0.0936) were removed from the model. This resulted in a final model with a significant effect of time spent on the BTT on the cubic transformed PT_BTTscore_Lines_ (β = 660.71, SE = 16.65, t(310.0) = 39.671, p < 0.0001) (see **Figure 3A**; for model details, see **Appendix 2A**). All subsequent progress test pairs showed a significant increase in PT_BTTscore_Lines_ (all, p < 0.0007 with Bonferroni correction α = 0.05/5 = 0.01; for tabular results, see **Appendix 2B**). In accordance with the non-significant effect of GROUP on PT_BTTscore_Lines_, comparing the motor learning slopes for the straight line tasks (PT_BTTslope_Lines_) yielded no significant difference between the groups (p = 0.9263, t(55.4) = -0.093, mean ± SD for simple / complex training group respectively: 0.076 ± 0.018 / 0.076 ± 0.025; see **Figure 3B**).

**Figure 3.**
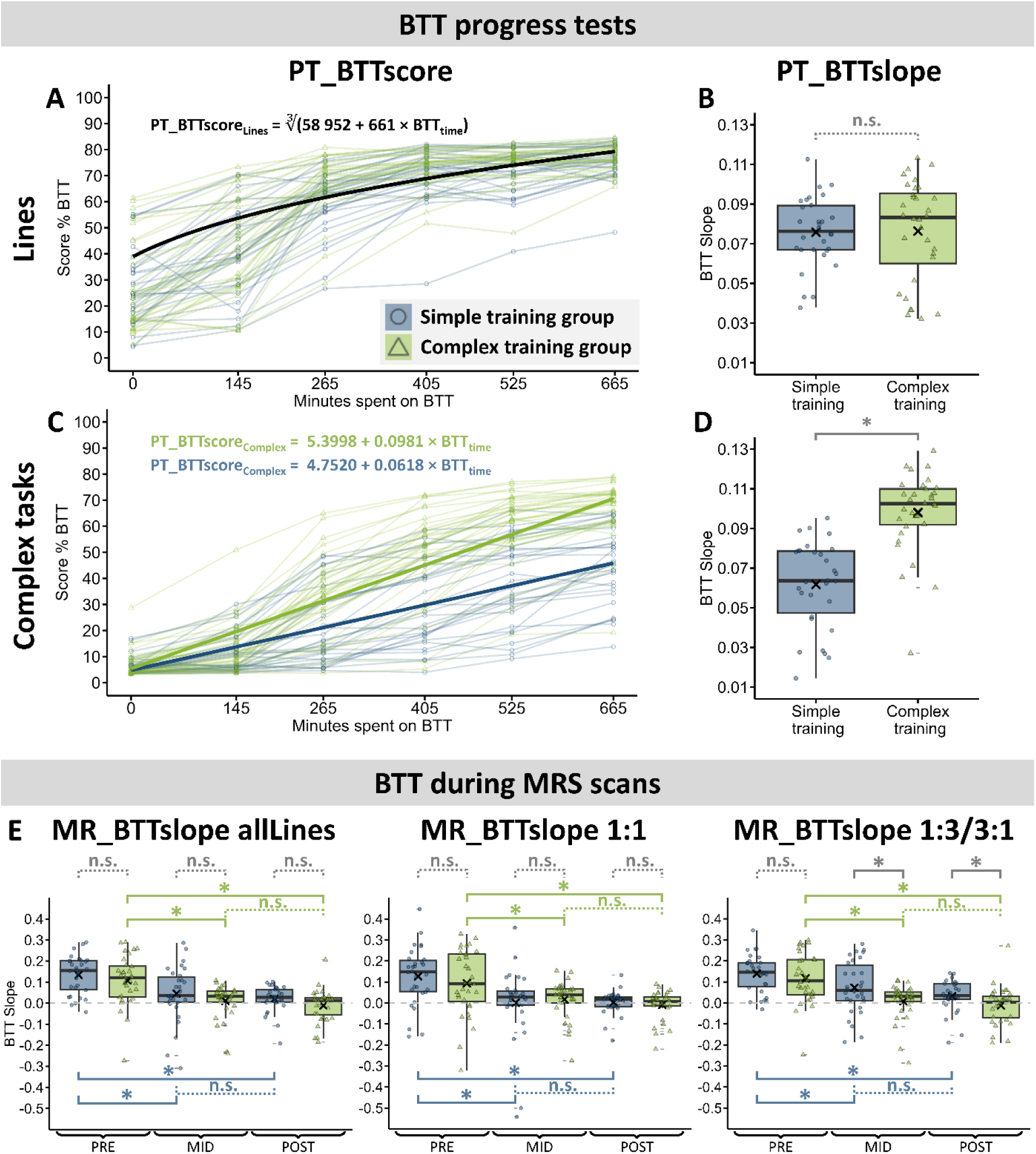
Results of the behavioral analysis of the bimanual tracking task (BTT). **[A]** and **[C]** Regression analysis of the BTT scores over the time spent on the BTT (in min) for the easier straight-line tasks and the complex tasks, respectively. **[B]** and **[D]** Long-term motor learning progress expressed as slopes of individual linear regressions over the progress test scores during the four-week motor learning paradigm, i.e., expressing the average improvement on the BTT Score (%) per minute training on the BTT. **[E]** Short-term motor learning progress expressed as slopes of individual linear regressions over the individual trials performed during the MRS acquisition at PRE, MID and POST measurement. Slopes indicate the average improvement on the BTT Score (%) per minute. Slopes close to 0 (gray dashed line) indicate no learning. **Boxplots:** The box and horizontal bar indicate the interquartile range (Q1–median–Q3), the whiskers span over the min and max values (up to 1.5 × IQR, outliers marked by a dash), and “×” marks the mean. Asterisks indicate a significant difference in BTT slope as compared to non-significant (n.s.) results.

#### Higher improvements on complex subtasks for the complex training as compared to the simple training group

In contrast, training group assignment did show an influence on performance and motor learning progress on the complex subtasks of the BTT progress test. For PT_BTTscore_Complex_, the time spent on the BTT (β = 0.0618, SE = 0.0027, t(310.00) = 22.967, p < 0.0001) and the interaction GROUP×BTT_time_ (GROUP×BTT_time_: F_1,310_ = 93.766, p < 0.0001; GROUP[complex]×BTT_time_: β = 0.0363, SE = 0.0037, t(310.00) = 9.683, p < 0.0001) had a significant effect on task performance (see **Figure 3C**; for model details, see **Appendix 2C**). When comparing the mean performances of the two groups for each progress test, group differences started to emerge from the third progress test onwards, but not the first two progress tests during which participants had not yet started the motor training program (PT1: p = 0.4733; PT2: p = 0.3462; PT3–6: all p < 0.004; with Bonferroni correction α = 0.05/6 = 0.0083; tabular results, see **Appendix 2D**; overview of measurements and progress tests, see **Figure 1A**). Moreover, all subsequent progress tests showed a significantly better performance for the later as compared to the earlier test moment in each of the groups separately (all, p < 0.0001, with Bonferroni correction α = 0.05/5 = 0.01 for each group; for tabular results, see **Appendix 2D**). The result of a significant GROUP×BTT_time_ effect on the progress test scores was supported by a significant GROUP difference in the learning slopes of the complex tasks (PT_BTTslope_Complex_; p < 0.0001, t(58.9) = -6.733, mean ± SD for simple / complex training group respectively: 0.062 ± 0.022 / 0.098 ± 0.020; see **Figure 3D**), indicating a steeper slope and consequently a faster learning progress for participants training on the complex training paradigm as compared to the simple one.

### 3.3 Behavioral outcomes during MRS acquisition

For the BTT learning progress during MRS scans, learning slopes (MR_BTTslope_allLines_, MR_BTTslope_11_, and MR_BTTslope_13,31_) for each group were steeper during the PRE measurement as compared to the MID and the POST measurement (all, p ≤ 0.0165, with Bonferroni correction α = 0.05/3 = 0.0167 for each group of three tests), but not different between the MID and POST measurement (all, p ≥ 0.0758). When comparing the two training groups at each of the three sessions across time, the learning slopes of the simple training group are steeper than the ones of the complex training group at MID and POST session for the MR_BTTslope_13,31_ (both p ≤ 0.0398, not surviving Bonferroni correction α = 0.05/3 = 0.0167), but not during the PRE session or for the MR_BTTslope_11_ or MR_BTTslope_allLines_ (see **Figure 3E**; tabular results, see **Appendix 2E**). The steeper learning slopes observed during the MRS acquisition in the MID and POST sessions for the simple training group, compared to the complex training group, are likely due to the lack of exposure to task variations during the simple training intervention. This resulted in a greater challenge for the simple training group participants and consequently led to more in-scanner motor learning progress.

### 3.4 Baseline neurometabolite levels to predict motor learning progress

#### No relationship between short-term motor learning and GABA+ or Glx levels

There was no relationship between short-term motor learning during the PRE measurement (i.e., in-scanner change in BTT performance, quantified as MR_BTTslope_allLines_) and the resting GABA+ or Glx levels in any of the VOIs during the PRE measurement (regression on GABA+ levels: stepwise removing GABA_R-SM1 [p = 0.9368], GABA_L-SM1 [p = 0.6082], GABA_L-PMd [p = 0.4271]; regression on Glx levels: stepwise removing Glx_R-SM1 [p = 0.9584], Glx_L-PMd [p = 0.3640], Glx_L-SM1 [p = 0.2405]).

#### Long-term motor learning on simple tasks is negatively associated with GABA+ levels in left PMd, but has no relationship with Glx levels

Long-term motor learning progress over the four-week training period on the simpler straight-line tasks (PT_BTTslope_Lines_) was negatively associated with GABA+ levels in left PMd (see **Figure 4A**). When regressed on GABA+ levels, the dependent variable first required a cubic transformation to ensure normality of the residuals and hence comply with model assumptions. In a stepwise procedure, the GABA_R-SM1×GROUP interaction (p = 0.3772), GABA_L-SM1×GROUP interaction (p = 0.2633), GABA_L-SM1 main effect (p = 0.3171), GABA_R-SM1 main effect (p = 0.3951), GABA_L-PMd×GROUP interaction (p = 0.1628) and GROUP main effect (p = 0.5022) were removed from the model. The final model (PT_BTTslope_Lines_)^3^ ∼ GABA_L-PMd (F_1,60_ = 4.667, p = 0.0348) showed a significant effect of GABA_L-PMd (β = -0.00027, SE = 0.00013, t(60) = -2.160, p = 0.0347), indicating that lower baseline GABA+ levels in left PMd are associated with better motor learning on the simpler task variants, independent of the training group (see **Appendix 3A** for model details). In comparison, there was no significant influence of Glx levels in either of the VOIs on PT_BTTslope_Lines_ (stepwise removing Glx_L-PMd×GROUP interaction [p = 0.9212], Glx_R-SM1×GROUP interaction [p = 0.5602], Glx_R-SM1 main effect [p = 0.8012], Glx_L-SM1×GROUP interaction [p = 0.3753], GROUP main effect [p = 0.4350], Glx_L-PMd main effect [p = 0. 5003], Glx_L-SM1 main effect [p = 0.2925]).

**Figure 4.**
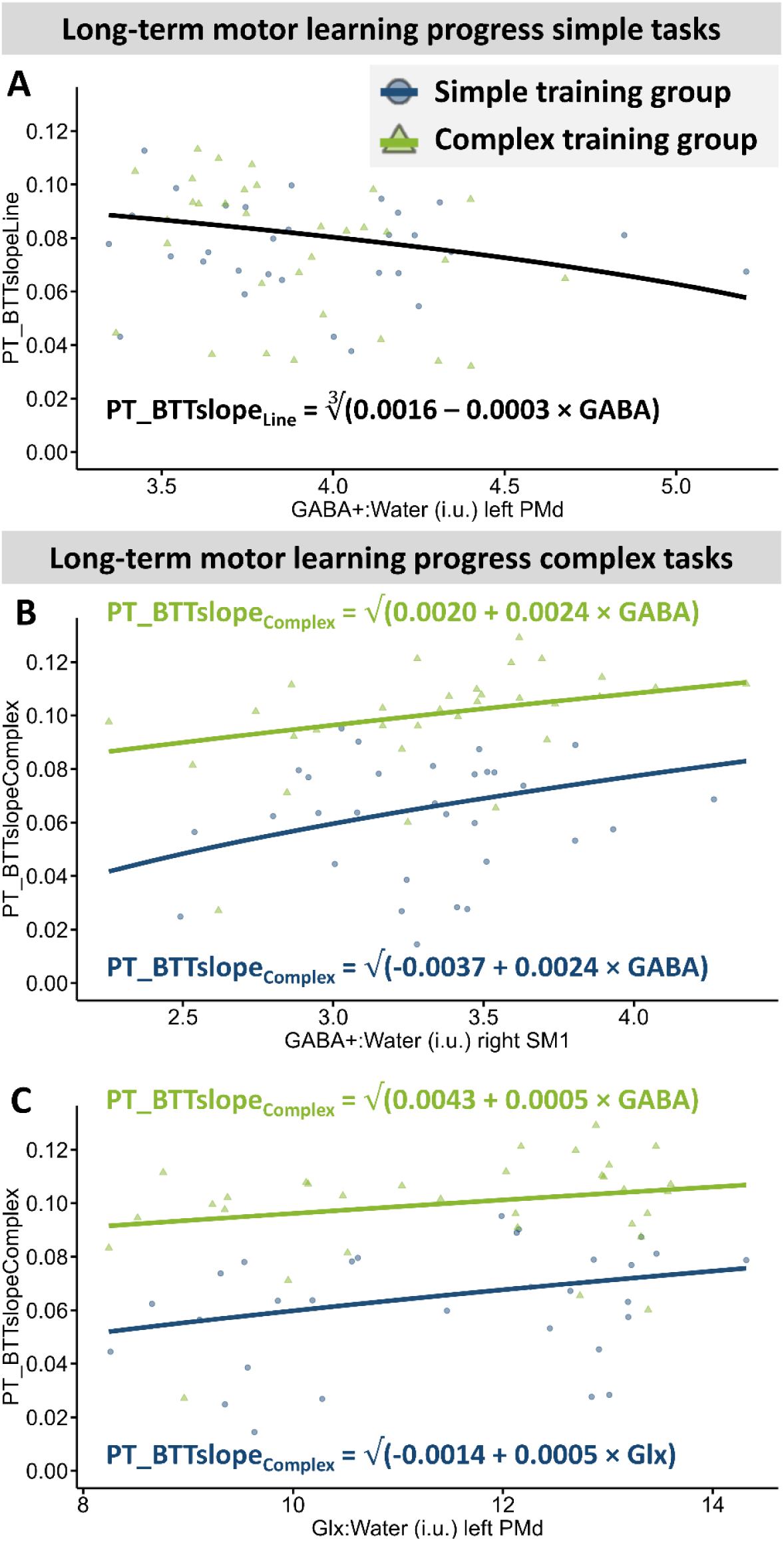
Effect of baseline neurometabolite levels on long-term motor learning progress. **[A]** Effect of GABA+ levels in left dorsal premotor cortex (PMd) on motor learning progress on simple tasks (PT_BTTslope_Line_). Since there was no effect of training group, only one regression line is displayed. **[B]** and [**C**] Effect of GABA+ levels in right primary sensorimotor cortex (SM1) (panel B) and Glx levels in left PMd (panel C) on motor learning progress on complex tasks (PT_BTTslope_Complex_). Since there was a significant effect of group, regressions for the simple (blue) and complex (green) learning groups are shown.

#### Long-term motor learning on complex tasks was positively associated with GABA+ levels in right SM1 and Glx levels in the left PMd

For the long-term motor learning progress on the complex task variants (PT_BTTslope_Complex_), GABA+ levels in right SM1 at baseline were positively associated with better motor learning (see **Figure 4B**). To comply with model assumptions, first a quadratic transformation of the dependent variable was conducted. Then, the model was simplified (stepwise removal of GABA_L-PMd×GROUP interaction [p = 0.8475], GABA_L-PMd main effect [p = 0.7069], GABA_L-SM1×GROUP interaction [p = 0.5192], GABA_L-SM1 main effect [p = 0.9515], GABA_R-SM1×GROUP interaction [p = 0.0979]). The final model (PT_BTTslope_Complex_)^2^ ∼ GABA_R-SM1 + GROUP (F_2,58_ = 35.810, p < 0.0001; see **Appendix 3B** for model details) showed a significant main effect of GROUP (GROUP[complex]: β = 0.00575, SE = 0.00073, t(58) = 7.856, p < 0.0001) and GABA_R-SM1 (β = 0.0024, SE = 0.00086, t(58) = 2.828, p = 0.0064). The main effect of GROUP indicates a steeper learning slope for the complex compared to the simple learning group (see **Figure 3D**). The main effect of GABA_R-SM1 indicates that higher GABA+ levels in right SM1 were linked to a steeper learning slope. Likewise, baseline Glx levels in the left PMd were positively associated with better motor learning (see **Figure 4C**). Again, a quadratic transformation of the dependent variable was necessary to comply with model assumptions. The model was simplified (removal of Glx_L-PMd×GROUP interaction [p = 0.7893], Glx_R-SM1×GROUP interaction [p = 0.4154], Glx_R-SM1 main effect [p = 0.8081], Glx_L-SM1×GROUP interaction [p = 0.1421], and Glx_L-SM1 main effect [p = 0.7846]), resulting in the final model (PT_BTTslope_Complex_)^2^ ∼ Glx_L-PMd + GROUP (F_2,59_ = 32.640, p < 0.0001; see **Appendix 3C** for model details). Motor learning progress on the complex task variants was significantly positively influenced by higher Glx levels in left PMd at baseline (β = 0.00050, SE = 0.00021, t(59) = 2.328, p = 0.0234), and was better for the complex as compared to the simple training group (GROUP[complex]: β = 0.00567, SE = 0.00074, t(59) = 7.655, p < 0.0001).

### 3.5 Changes in resting neurometabolite levels as a result of motor learning

#### Left SM1 GABA+ levels: no change

For the left SM1 VOI, resting GABA+ levels were not significantly influenced by motor learning. More specifically, first the GROUP×SESSION interaction (p = 0.9375) and then the SESSION main effect (p = 0.7678) were removed from the model, resulting in the final model GABA_L-SM1 ∼ GROUP + (1|SubjectID) (observations = 185, groups in random effects = 62) with a significant effect of GROUP (GROUP[complex]: β = 0.170, SE = 0.849, t(62.15) = 2.005, p = 0.0493). Since this result was not of interest but rather an overall group difference persisting over all three measurements, no further model details are being reported.

#### Left SM1 Glx levels: decrease over four weeks of training, but only for the complex training group

The Glx levels in the left SM1 VOI showed to be related to a GROUP×SESSION interaction (see **Figure 5A**). The model assumptions were fulfilled without any transformation and since the model yielded an interaction effect, no further simplification of the model was performed (final model: Glx_L-SM1 ∼ GROUP + SESSION + GROUP×SESSION + (1|SubjectID) (observations = 185, groups in random effects = 62). The interaction effect of GROUP×SESSION had a significant influence on Glx levels in left SM1 (GROUP×SESSION: F_2,123.13_ = 4.769, p = 0.0101; GROUP[complex]×SESSION[MID]: β = -0.604, SE = 0.310, t(123.24) = -1.945, p = 0.0540; GROUP[complex]×SESSION[POST]: β = -0.947, SE = 0.310, t(123.24) = -3.053, p = 0.0028), but neither the main effect GROUP (p = 0.4287) nor SESSION (p = 0.5505) had a significant influence. To explore this interaction effect, one LMM per GROUP was constructed according to the formula Glx_L-SM1 ∼ SESSION + (1|SubjectID). This model showed a significant effect of SESSION for the complex training group (SESSION: F_2,63.15_ = 3.673, p = 0.0310; SESSION[MID]: β = -0.208, SE = 0.218, t(63.26) = -0.955, p = 0.3435; SESSION[POST]: β = -0.581, SE = 0.218, t(63.26) = -2.669, p = 0.0097), but not the simple training group (p = 0.1441). Post hoc tests within the complex training group revealed, that the effect of SESSION on the Glx levels of the left SM1 resulted from a difference between PRE and POST measurement (p = 0.0135, t(30) = 2.624, mean difference = 0.607, 95% CI = 0.135–1.080; surviving Bonferroni correction α = 0.05/3 = 0.0167), but that neither PRE and MID (p = 0.3369) nor MID and POST (p = 0.0986) measurements significantly differed from each other, indicating that the Glx levels at rest in the left SM1 of the complex training group decreased significantly when looking at the full four weeks of motor training, but that the effect was not strong enough to reach significance for the intermediate comparisons of two weeks each (PRE vs. MID, MID vs. POST; for details on the LMMs and post hoc tests, see **Appendix 4A**).

**Figure 5.**
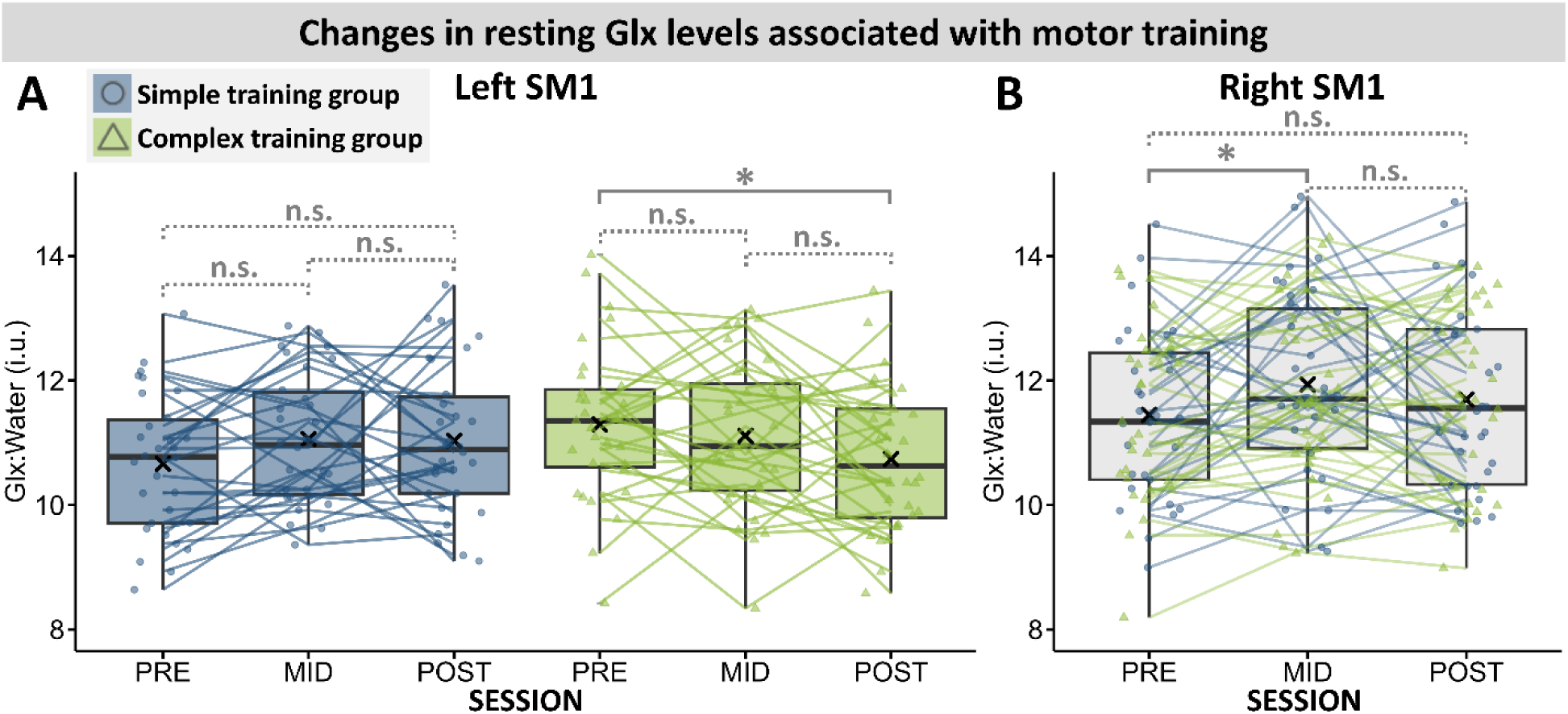
Changes in resting Glx levels associated with motor training intervention. **[A]** Changes in resting Glx levels in the left primary sensorimotor cortex (SM1). Groups are displayed separately due to the significant effect of group. **[B]** Changes in resting Glx levels in the right SM1, with means pooled for the two training groups. **Boxplots:** The box and horizontal bar indicate the interquartile range (Q1–median– Q3), the whiskers span over the min and max values (up to 1.5 × IQR, outliers marked by a dash), and “×” marks the mean. Asterisks indicate a significant difference in Glx levels as compared to non-significant (n.s.) results.

#### Right SM1 GABA+ levels: no change

For the right SM1, resting GABA+ levels did not change significantly with motor training. The GROUP×SESSION interaction (p = 0.7580), SESSION main effect (p = 0.7891) and GROUP main effect (p = 0.5102) showed no significant influence on GABA+ levels.

#### Right SM1 Glx levels: increase during the first two weeks of motor training

For the right SM1 VOI the Glx levels changed with motor training, independent of the group assignment (see **Figure 5B**). In a stepwise backwards simplification, the GROUP×SESSION interaction (p = 0.5380) and GROUP main effect (p = 0.6871) were removed from the model to obtain the final model Glx_R-SM1 ∼ SESSION + (1|SubjectID) (observations = 185, groups in random effects = 62). SESSION significantly influenced resting Glx_levels in right SM1 (SESSION: F_2,123.2_ = 4.159, p = 0.0179; SESSION[MID]: β = 0.474, SE = 0.165, t(123.28) = 2.884, p = 0.0046; SESSION[POST]: β = 0.234, SE = 0.165, t(123.28) = 1.421, p = 0.1578), and post hoc tests revealed a significant increase in Glx levels from PRE to MID session (p = 0.0043, t(60) = -2.968, mean difference = - 0.476, 95% CI = -0.797– -0.155; surviving Bonferroni correction α = 0.05/3 = 0.0167), but no significant differences between PRE and POST (p = 0.1499) or MID and POST (p = 0.1933) measurements. This indicates an increase in resting Glx levels of the right SM1 in the first two weeks of motor training, independent of the training group, that partially renormalized towards the POST measurement (for details on LMMs and post hoc tests, see **Appendix 4B**).

#### Left PMd GABA+ levels: no change

For the left PMd, resting GABA+ levels did not change significantly with four weeks of motor training (stepwise removed GROUP×SESSION interaction [p = 0.4378], SESSION main effect [p = 0.4170], and GROUP main effect [p = 0.3237] from LMM).

#### Left PMd Glx levels: no change

Resting Glx levels in left PMd did not change either as a result of the motor training intervention (stepwise removed GROUP×SESSION interaction [p = 0.2047], GROUP main effect [p = 0.794], and SESSION main effect [p = 0.3240] from LMM).

### 3.6 Modulation of left PMd neurometabolite levels during task execution and changes in modulation with motor learning

#### No modulation of the GABA+ or Glx levels in left PMd associated with the task performance in the scanner during the PRE measurement

For GABA+, the model was simplified (removal of GROUP×TASK interaction [p = 0.7351], TASK main effect [p = 0.6359], and GROUP main effect [p=0.5216]; final model with only Water_FWHM covariate of no interest [p = 0.0566]), and yielded neither an effect of the training paradigm, nor of the task execution in the scanner on GABA+ levels in left PMd during the PRE measurement. Similarly, Glx levels were not influenced by training group or task execution (stepwise removed GROUP×TASK interaction [p = 0.7603], TASK main effect [p = 0.8413], and GROUP main effect [p = 0.5814]; final model with only Water_FWHM covariate of no interest [p = 0.7769]).

#### No change in GABA+ or Glx modulation associated with motor training

Stepwise backwards regression using LMMs yielded no significant influence of measurement session, task execution, or training group assignment on GABA+ levels in left PMd when investigating data of all three measurement sessions (stepwise removal of GROUP×TASK interaction [p = 0.8245], SESSION×TASK interaction [p = 0.6450], TASK main effect [p = 0.7699], GROUP×SESSION interaction [p = 0.2710], SESSION main effect [p = 0.9326], GROUP [p = 0.6478]; final model with only Water_FWHM covariate of no interest [p = 0.3463]). Likewise, no influence of task execution on Glx levels in left PMd was found. However, the LMM yielded changes in Glx levels of left PMd when pooled over the four MRS acquisition blocks (BLOCK_RESTbefore, BLOCK_BTT1, BLOCK_BTT2, BLOCK_RESTafter; due to the removal of the TASK main and interaction effects). More specifically, stepwise removal of the SESSION×TASK interaction (p = 0.9914), GROUP×TASK interaction (p = 0.6790), and TASK main effect (p = 0.6112), resulted in the final model: Glx_L-PMd ∼ GROUP + SESSION + GROUP×SESSION + Water_FWHM + (1|SubjectID) (observations = 744, groups in random effects = 62). This model showed a significant influence of SESSION (p = 0.0005), the GROUP×SESSION interaction (p = 0.0017), and the covariate Water_FWHM (p = 0.0140), but not the main effect of GROUP (p = 0.5015). Since this result corresponds to a change in Glx levels (and more specifically, a decrease in Glx levels of the complex training group towards the POST measurement) when pooling the data over the four MRS acquisition blocks in the left PMd (which was not a main question of this research), the results of this LMM analysis including a visualization are reported in **Appendix 5A**. The task-related changes in GABA+ and Glx per subject and session are visualized in **Appendix 5B**.

### 3.7 Relationship between maximal modulation of neurometabolite levels and motor learning

#### The absolute maximal modulation at the PRE measurement is predictive of short-term but not long-term motor learning

Short-term motor learning within the PRE measurement session (MR_BTTslope_allLines_) was related to the absolute maximal modulation (|MAXmod|) of neurometabolites in left PMd. After removal of one outlier with a highly negative learning slope that distorted the residuals’ normality in a way that could not be corrected by transformations, no further model simplification was necessary: MR_BTTslope_allLines_ ∼ GABA_|MODmax| + Glx_|MODmax| + GABA_|MODmax|×Glx_|MODmax| (F_3,57_ = 3.645, p = 0.0178), with a significant influence of GABA_|MODmax| (β = 0.2451, SE = 0.0913, t(57) = 2.686, p = 0.0095), Glx_|MODmax| (β = 0.1662, SE = 0.0565, t(57) = 2.942, p = 0.0047), and the interaction GABA_|MODmax|×Glx_|MODmax| (β = -0.2442, SE = 0.1031, t(57) = -2.368, p = 0.0213). The interaction effect reflects that individuals with a high task-related absolute modulation of GABA+ or Glx at the PRE measurement had a steeper motor learning slope during the PRE measurement as compared to individuals showing a high modulation in both neurometabolites simultaneously or no modulation in either of the neurometabolites. Moreover, moderate modulation of both neurometabolites resulted in moderate motor learning slopes (see **Figure 6**; see **Appendix 5C** for model details). Interestingly, for the interaction between the two metabolites, a data-driven tipping point for each neurometabolite was estimated, at which the relationship between the remaining metabolite modulation and motor learning changed the sign of the slope (see **Figure 6B**). For GABA_|MAXmod| values below or above 0.68, Glx_|MAXmod| had, respectively, a positive or negative influence on short-term motor learning progress. Likewise, for Glx_|MAXmod| values below or above 1.00, GABA_|MAXmod| have a positive or negative association with short-term motor learning, respectively.

**Figure 6.**
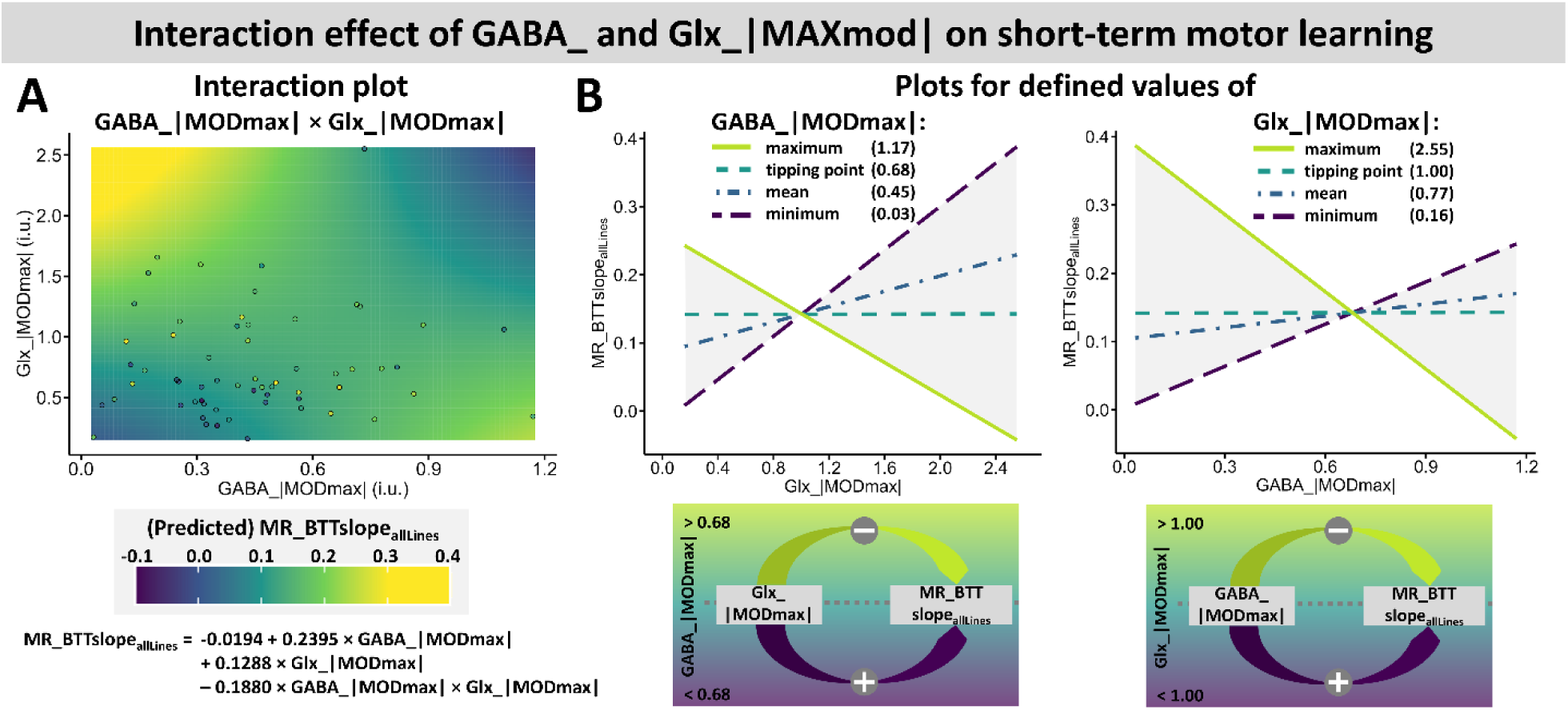
Effect of absolute maximal modulation interaction between GABA+ and Glx on short-term motor learning during PRE measurement. **[A]** Interaction plot and formula with individual data points. High absolute maximal modulation (|MAXmod|) of either GABA+ or Glx, but not both simultaneously or neither, predicts better motor learning, creating a balance when both are moderately modulated. **[B] Left:** Influence of Glx_|MAXmod| on short-term motor learning progress (MR_BTTslope_allLines_) for defined values of GABA_|MAXmod|. **Right:** Influence of GABA_|MAXmod| on short-term motor learning progress (MR_BTTslope_allLines_) for defined values of Glx_|MAXmod|. For both graphs, the minimum, mean, and maximum measured value for the neurometabolite absolute modulation was plotted. The “tipping point” value describes the value at which the influence of the neurometabolite on the x-axis on MR_BTTslope_allLines_ changes from a positive to a negative association or vice versa. Predictable gray areas are located between the minimal and maximal values that |MAXmod| assumed in the present study.

In contrast, long-term motor learning progress was not associated with the absolute maximal neurometabolite modulation. More specifically, there was no relationship between the absolute maximal modulation during the PRE measurement and the subsequent four-week motor learning for the learning progress on the simple (PT_BTTslope_Lines_; quadratic transformation, then stepwise removal of GABA_|MODmax|×Glx_|MODmax| [p = 0.2887], GABA_|MODmax| [p = 0.8863], and Glx_|MODmax| [p = 0.6816]) or the complex subtasks (PT_BTTslope_Complex_; quadratic transformation, then stepwise removal of GABA_|MODmax|×Glx_|MODmax| [p = 0.7262], Glx_|MODmax| [p = 0.9349], GABA_|MODmax| [p = 0.8672]).

#### No relationship between directional maximal modulation at PRE measurement and motor learning

The measure of the directional absolute modulation (MAXmod) within left PMd during the PRE measurement was neither predictive of short-term motor learning (MR_BTTslope_allLines_), nor of simple (PT_BTTslope_Lines_) or complex long-term motor learning (PT_BTTslope_Complex_). More specifically, for the regression of MR_BTTslope_allLines_ one outlier with a highly negative learning slope had to be removed from the analysis (same as for |MAXmod| regression). Subsequently, the GABA_MODmax×Glx_MODmax interaction (p = 0.8649), Glx_MODmax main effect (p = 0.9871), and the GABA_MODmax main effect (p = 0.7867) were stepwise removed from the model, resulting in an intercept-only model. For the regressions of PT_BTTslope_Lines_ and PT_BTTslope_Complex_, first the interaction GABA_MODmax×Glx_MODmax (p = 0.5068 and p = 0.1026, respectively), then the GABA_MODmax main effect (p = 0.2986 and p = 0.9864, respectively) and the Glx_MODmax main effect (p = 0.0978 and p = 0.9098, respectively) were stepwise removed from the model.

## 4 Discussion

We investigated the role of GABA+ and Glx levels in human motor learning and revealed four main results. First, the four-week training period on a complex bimanual coordination task led to increases in performance that were significantly higher for participants enrolled in a complex as compared to a simple task training paradigm, and the complex subtasks continued to improve until the end of the training period. Second, baseline GABA+ levels in right SM1 and Glx levels in left PMd showed a positive association with the learning progress on the complex subtasks of the BTT, whereas baseline GABA+ levels in left PMd were negatively associated with long-term motor learning progress on the simpler subtasks. There was no relationship between baseline neurometabolite levels and short-term motor learning. Third, left SM1 Glx levels at rest decreased over four weeks of motor training for the complex but not simple training group, whereas Glx levels in right SM1 increased during the first two weeks of motor training, independent of the training group. No change in resting GABA+ levels was reported in any of the VOIs. Likewise, for the left PMd, no changes in resting GABA+ or Glx levels were reported. Fourth, there was no task-related modulation in left PMd GABA+ or Glx levels at group level. However, the task-related absolute maximal modulation of GABA+ and Glx levels in left PMd, assessed at the initial stage of motor training intervention, showed a relationship with short-term but not long-term motor learning progress, indicating the importance of the ability to adapt or tune excitation and inhibition in function of the task requirements to facilitate motor learning.

### 4.1 Ongoing motor learning until the end of the training intervention

We implemented a four-week bimanual motor training regimen designed to induce learning and consequent neuroplasticity in task-related brain regions, and to investigate the role of excitatory and inhibitory neurotransmitters during this process. The motor training led to ongoing performance improvements up to the end of the four-week intervention period, equivalent to a total of approximately 11 hours spent on the BTT. Notably, each subsequent pair of progress tests up to the end of the intervention showed significant improvements for both training groups and on both subtask classifications, i.e., the simpler straight-line tasks and the complex tasks. This finding contrasts with earlier studies (Beets et al., 2015; Solesio-Jofre et al., 2018) employing only straight-line subtasks of the BTT, which reported a performance plateau after four hours of motor training. In contrast, in the current study, the relatively linear performance gains on the complex subtasks, that were sustained until the end of the training period, provide a solid behavioral foundation for examining neuroplastic changes associated with motor learning.

In contrast to the complex training group, the simple training group only practiced the iso-frequency (1:1 line) condition, a movement pattern that is part of the intrinsic motor repertoire (Wenderoth et al., 2005), and was only exposed to more difficult task variations during the progress tests (∼5 min each). Hence, we expected similar performance gains of both groups on the straight-line tasks, but smaller improvements in the simple as compared to the complex training group for the complex subtasks of the BTT. In line with this hypothesis, the results indicated no group difference in the motor learning progress on the straight-line tasks. However, for the complex subtasks of the BTT the complex training group outperformed the simple training group from the third progress test on (i.e., as soon as the actual training program started, since the first two progress tests were acquired before the first training week). This indicates faster and more profound learning in the complex as compared to the simple training group for the complex subtasks.

### 4.2 Baseline neurometabolite levels in right SM1 and left PMd predict long-term motor learning progress

Investigating the relationship between baseline neurometabolite levels and subsequent short- and long-term motor learning progress yielded no relationship between baseline GABA+ or Glx levels and short-term motor learning. In contrast, for long-term motor learning, significant associations with right SM1 GABA+ levels, as well as left PMd GABA+ and Glx levels were observed. More specifically, left PMd GABA+ levels were negatively associated with long-term motor learning progress on simpler subtasks, whereas the Glx levels in left PMd showed a positive association with the learning progress on complex subtasks of the BTT. Furthermore, baseline GABA+ levels in right SM1 also showed a positive relationship with long-term motor learning on the complex tasks.

#### Baseline GABA+ levels and long-term motor learning on the simple tasks

We found a significant association between lower GABA+ levels in left PMd and better long-term motor learning on the simple subtasks, and between higher GABA+ levels in right SM1 and better motor learning on the complex subtasks of the BTT. Although there is no previous evidence on the role of left PMd and right SM1 neurometabolite levels at baseline on the subsequent motor learning progress, there have been various reports on the role of GABA and Glx baseline levels in left SM1 for motor learning. However, this prior evidence only concerns shorter periods of motor learning up to five days. For baseline GABA levels in left SM1, results of Chalavi et al. (2018) were in line with the current findings. More specifically, GABA levels in the left SM1 were not related to the learning progress of a three-day BTT training, despite a negative association between GABA levels in left SM1 and initial performance (Chalavi et al., 2018). Similar results were reported for an SRTT, where baseline GABA levels in left SM1 were negatively correlated with initial performance, but showed no correlation with motor learning (Stagg et al., 2011). In contrast, other studies did show positive or negative associations between GABA levels in SM1 and motor learning. One study showed a positive association between baseline GABA levels in M1 (when no direct feedback was provided) or S1 (when direct visual feedback was given) and the initial learning progress on the BTT, but no association with the later learning progress over a five-day BTT training (Li et al., 2024). Another study, however, reported a negative association between baseline GABA levels in left M1 and subsequent motor learning within a 40-minute training session on a sequential reaction time task performed with the right hand (Kolasinski et al., 2019).

These findings are not surprising in light of the importance of GABA in motor learning. A single administration of baclofen, a GABA_B_ receptor agonist which reduces neuronal excitatbility, has been shown to impair subsequent visuomotor learning (Johnstone et al., 2021) and neuroplastic LTP-like neuroplastic processes (McDonnell et al., 2007). Along the same lines, another study reported a decrease in GABA-related inhibition to facilitate practice-dependent plasticity in the motor cortex both neurophysiologically and behaviorally, whereas an increase in GABA_A_-related inhibition via lorazepam administration depressed neuroplasticity (Ziemann et al., 2001). Hence, it could be hypothesized that lower GABA levels in motor-related brain regions at baseline may enable an increased communication between neurons during motor practice. This would facilitate LTP-like neuroplasticity (Kim et al., 2014), which in turn can lead to alterations in synaptic communication (Castillo et al., 2011; Sanes & Donoghue, 2000) and hence a better consolidation of motor skills (Harms et al., 2008). However, also higher baseline GABA levels have been linked to better motor learning (Li et al., 2024). One possible hypothesis for the underlying mechanism of these opposite results might be that higher baseline GABA levels could provide a greater range for learning-related decreases in GABA, allowing more room for modulation and, consequently, better motor learning (King et al., 2020). Furthermore, the effect might be region-specific. For example, the current study yielded lower GABA+ levels in left PMd to be associated with better long-term motor learning. This decreased inhibition might be in line with the increased activation of PMd during complex bimanual motor tasks (Van Ruitenbeek et al., 2023).

#### Baseline Glx levels and long-term motor learning on the complex tasks

In addition, a positive association between higher Glx levels in left PMd and subsequent long-term motor learning on the complex subtasks was shown. Again, there is no prior evidence on the link between the PMd’s neurometabolite levels and motor learning progress, and only results for shorter motor learning interventions are available. For the left SM1, however, previous work reported no relationship beween baseline Glx or Glu levels and subsequent motor learning. More specifically, one study reported no link between baseline Glx levels in left M1 or S1 and initial or later learning progress (Li et al., 2024). Similarly, for an SRTT, no relationship between left M1 Glu (Kolasinski et al., 2019) or left SM1 Glx levels (Stagg et al., 2011) at baseline and learning progress within the same session was reported. In contrast, one study did report a positive relationship between baseline Glx levels in left SM1 and motor learning of an SRTT (Bell et al., 2023).

As the main excitatory neurotransmitter in the mammalian brain, the role of glutamatergic neurotransmission for motor learning is vital, since it underlies the synaptic plasticity processes necessary for acquiring and refining motor skills (Zhou & Danbolt, 2014). This association has become more clear through animal research, in which the advantages of variable over constant practice were linked to a greater dependency on the glutamate receptors n-methyl-D-aspartate (NMDA) and more expression of the alfa-amino-3-hydroxy-5-methyl-4-isoxazolepropionic (AMPA) receptor has been shown (Apolinário-Souza et al., 2020). Furthermore, it has been repeatedly described that the induction of LTP- and LTD-like plasticity are dependent on the NMDA receptor (for review, see Lüscher & Malenka, 2012). Besides this link between neuroplasticity and glutamatergic neurotransmission, also a positive association between brain activation (as measured with fMRI) and Glu levels has been shown (Maruyama et al., 2021), which might indicate the importance of Glu to effectively activate a brain region.

#### Right vs. left SM1

This study yielded no links between motor learning for the baseline neurometabolite levels in the left SM1, but a positive association between baseline GABA+ levels in right SM1 and complex motor learning. There are several possible explanations as to why the baseline neurometabolite levels in the right (non-dominant) but not the left (dominant) SM1 were predictive of motor learning. First, in general, turning the BTT dial with the dominant hand might be easier compared to the non-dominant hand, leading to greater motor learning effects in, and greater importance of, the right non-dominant SM1. Behaviorally, this is supported by a BTT study that reported higher scores for straight-line tasks when the dominant hand had to move faster than the non-dominant hand, as compared to the opposite condition (Sisti et al., 2011). Second, the encoding of right/left movements via the right/left dial rotations using the dominant hand might be more straight-forward than the up/down encoding via the non-dominant hand, and hence might have required more learning to cope with this less compatible visuomotor transformation, making the effect of baseline neurometabolite levels in the non-dominant SM1 and especially GABA to shape motor output more influential.

#### Left PMd

Lower GABA+ and higher Glx levels in left PMd at baseline were associated with better long-term motor learning on simple or complex tasks of the BTT, respectively. These findings can potentially be explained by fMRI findings on the role of left PMd in motor tasks and motor learning. More specifically, task-related premotor activity has been shown to increase with BTT complexity (Van Ruitenbeek et al., 2023). The observed higher excitatory Glx levels in left PMd might hence be a prerequisite for a better task-related activation of left PMd, in turn allowing for better movement planning and control during complex BTT trials. Furthermore, left PMd activation has been associated with better motion smoothness (Sosnik et al., 2014), an important feature for obtaining high scores on the BTT especially when tracing straight lines. This might be linked to the observed correlation with lower baseline GABA levels, which might promote excitation, and hence allow a better activation of the left PMd to accomplish smooth dial rotations. Overall, the current findings are in support of the diverse literature that underlines the PMd’s central role in fine-tuning the motor output of M1 (Chouinard & Paus, 2006; Dum & Strick, 1991), planning of bimanual movements (Beets et al., 2015; Debaere et al., 2004; Fujiyama et al., 2016; Verstraelen et al., 2021), response selection (Chouinard & Paus, 2006; Crammond & Kalaska, 2000), and in learning a diversity of motor tasks (Hardwick et al., 2013).

### 4.3 Changes in resting left and right SM1 Glx levels associated with motor training

An interesting yet unresolved question is whether training does induce lasting changes in the baseline levels of neurometabolites or whether such changes are only transient and short-lived. Here, changes in resting Glx levels of left and right SM1 were observed over a four-week training paradigm. More specifically, left SM1 Glx levels decreased after four weeks of motor training in the complex but not the simple training group. In contrast, Glx levels in right SM1 increased during the first two weeks of motor training, independent of the training group. There were no changes in resting GABA+ levels in any of the VOIs, or in the resting GABA+ or Glx levels of the left PMd.

Previous evidence on changes in resting neurometabolite levels with long-term motor learning interventions are very scarce. Only in one study, left SM1 neurotransmitter levels were investigated over a longer period, i.e., a six-week juggling training intervention. A decrease in GABA was reported for a low-intensity (i.e., 15 min, 5 ×/week), but not high-intensity (i.e., 30 min, 5 ×/week) training group (Sampaio-Baptista et al., 2015). This result is in contrast with the current study, where no changes in resting GABA+ levels were reported for any of the VOIs after two or four weeks of motor training. However, more in line with our findings, in a study employing a shorter three-day BTT training intervention, no changes in left SM1 GABA levels were reported (Chalavi et al., 2018). Changes in measurement timings, training intensity, and task requirements could have resulted in these discordant results. Unfortunately, neither of these studies (Chalavi et al., 2018; Sampaio-Baptista et al., 2015) investigated changes in Glu or Glx.

Of note is the opposite effect of the motor training intervention on Glx levels of the left and right SM1, with a decrease in Glx only in the complex training group after four weeks of training for the left SM1, and an increase in Glx for both groups after two weeks of training, with a partial (non-significant) renormalization after four weeks of motor training. Potentially, effects of lateralization and the frequency of hand-use might play a role in this context. More specifically, the bimanual motor training might have led to an increase in excitation in the non-dominant (right) SM1 due to the increased use and refinement of left hand control as compared to baseline. In contrast, the dominant (left) SM1 in control of the right hand is by default exposed to high dexterity demands. This might have resulted in a lack of changes in the simple training group, and a decrease in Glx in the complex training group in order to shift the laterality in favor of the non-dominant hemisphere, a requirement which might be more prominent during complex rather than simple task training. However, to date motor learning studies reporting neurochemical levels in both SM1s simultaneously are lacking, and hence this hypothesis is speculative. Evidence from fMRI studies has shown partially dissimilar effects of five days of BTT training on brain activity of the right and left SM1. For example, one study reported a decreased activation of left M1 and right S1 during the planning phase of the BTT, whereas during the task execution phase of the BTT bilateral S1 and M1 were less activated after the training intervention as compared to baseline (Beets et al., 2015). Furthermore, an increased intra-hemispheric connectivity within the motor network of the right but not left hemisphere has been shown after a five-day BTT training intervention as compared to baseline (Solesio-Jofre et al., 2018). Taken together, bimanual motor training might lead to dissimilar effects in both hemispheres, potentially reinforced by slightly different task demands for the left versus the right hand (as discussed in the previous section) that might have led to the differential changes in resting Glx levels of the left and right SM1.

### 4.4 No group-level modulation of neurometabolite levels in left PMd during task execution, but link between individual modulatory ability of left PMd neurometabolites and short-term motor learning

Lastly, this research tackled the question of whether there is a modulation of left PMd neurometabolite levels during task execution and how this relates to motor learning. First, we investigated the differences between resting or task-related acquisition blocks, and the changes in this modulation with four weeks of motor learning. This yielded no modulation in GABA+ or Glx levels at the group level. However, at the level of the individual, the task-related absolute maximal modulation of GABA+ and Glx levels in left PMd during the PRE measurement session was associated with short-term but not long-term motor learning progress. More specifically, there was an interaction effect of GABA+ and Glx absolute modulation on short-term motor learning during the PRE measurement that indicated best learning progress for individuals with either a high GABA+ or a high Glx modulation, medium learning progress for individuals with simultaneous GABA+ and Glx modulation in a balanced manner, and least learning progress when neither of the neurometabolites were modulated or both showed a strong modulation. This finding points towards the importance of the ability to adapt or tune excitation and inhibition in function of the task requirements to facilitate motor learning.

#### Glx modulations

A meta-analysis on task-related modulation of Glu/Glx yielded increased Glu and Glx levels in task-related brain regions during task execution for various task domains (Pasanta et al., 2022). For the motor domain, this analysis on SM1 data included studies investigating finger-tapping (Schaller et al., 2014) and rhythmic hand clenching (Chen et al., 2017; Volovyk & Tal, 2020). In the present study, we could not replicate these findings for the left PMd, which was more in line with a report of no change in SM1 Glu levels with motor learning (Bell et al., 2023). There are several possible explanations for this finding. First, the task employed in the present study has higher cognitive and coordinative demands than the tasks investigated in the previous reports, with hand clenching specifically requiring force rather than fine precision motor control (as required for the BTT). These different task requirements might have influenced the results. Second, although the meta-analysis revealed Glu/Glx changes across various brain areas including visual, cognitive, and learning-related task domains, there are no prior results on the premotor region. Third, MRS acquisition and quantification of Glu/Glx can be obtained using various approaches that influence the final quantification. In this study, a MEGA-PRESS sequence was used to acquire MRS spectra in order to study changes in GABA+ levels. Furthermore, the fitting algorithm included in the Gannet toolbox is specifically optimized for the quantification of GABA signals, with the Glx quantification rather being a byproduct, since neither the acquisition parameters nor the analysis is perfectly tailored to it. This also results in the pooling of Glu and glutamine signals into the Glx signal, which might conceal changes in Glu levels due to opposing changes in glutamine signals. Lastly, functional MRS can be analyzed either blockwise (i.e., per acquisition block), which has limited temporal resolution, or in an event-related fashion, which can be more sensitive to transient changes in neurometabolites (Koolschijn et al., 2023). The blockwise analysis employed in the current study has a coarse temporal resolution of 11 minutes per block, potentially concealing short-term neurotransmitter modulations. However, detecting small metabolites such as GABA at 3T requires sufficient scanning time to achieve a sufficiently high SNR, limiting the feasibility of event-related designs at commonly available magnetic field strengths.

#### GABA+ modulations

For GABA, a meta-analysis (Pasanta et al., 2022) reported a non-significant tendency towards task-related decreases in GABA levels. However, for the motor domain, only two studies (Chen et al., 2017; Kolasinski et al., 2019) examining GABA modulation were included in this meta-analysis. These studies reported a decrease in GABA levels after several minutes of continuous rhythmic hand clenching (Chen et al., 2017) and during motor learning on an SRTT (Kolasinski et al., 2019). Notably, both studies were conducted using a 7T MR scanner and hence have a better spectral resolution than the 3T results presented here. However, also one study conducted at 3T was able to show GABA reductions with learning of a serial force production task (Floyer-Lea et al., 2006). In contrast, other 3T and 7T studies have reported no motor task-related GABA modulation for finger-tapping (Schaller et al., 2014), motor learning on an SRTT (Bell et al., 2023; Eisenstein et al., 2023, 2024), or during task transfer after three days of motor learning (Rasooli et al., 2024). Comparison with the here presented finding of no modulation in GABA+ levels during bimanual motor learning in the left PMd should, however, be done with care, as previous evidence is based on results from SM1, not PMd.

#### Interaction effect of absolute GABA+ and Glx modulation on short-term motor learning

Although the present study yielded no modulation in GABA+ or Glx levels at the group level with motor learning, there was an interaction between the inter-individual amount of absolute GABA+ and Glx modulation and short-term motor learning during the PRE measurement. Several studies support the notion that the ability to modulate brain metabolites is essential for successful motor learning. One study employing anodal tDCS yielded that the ability to modulate (i.e., decrease) GABA levels in left M1 was associated with better motor learning on an SRTT (Stagg et al., 2011). Furthermore, greater decreases in the GABA/Glu ratio within a single session of an SRTT training were correlated with higher performance gains (Maruyama et al., 2021). Additionally, higher Glu increases after SRTT motor learning were predictive of higher overnight offline learning gains (Eisenstein et al., 2024). Taken together, these results point towards the importance of the brain’s flexibility to modulate neurometabolites efficiently for better task performance and learning success, and more specifically shift the E-I balance in the direction of less inhibition and more excitation. It also appears that the process of motor learning and its neural correlates is characterized by high inter-individual differences, that might be too subtle or too diverse to be revealed at the group level.

## 5 Conclusion

In this study, we aimed to address the gap in evidence regarding neurometabolic changes associated with long-term motor learning. We investigated the relationship between inhibitory and excitatory neurometabolites and the motor learning process. This study yielded that an initially more excitatory neurometabolite profile in left PMd, and increased inhibition in non-dominant SM1 predicted long-term motor training success. Furthermore, after two weeks of motor training, the resting excitatory neurometabolite levels in the non-dominant SM1 showed a transient increase regardless of training complexity, which partially renormalized after four weeks. For the dominant SM1, however, the excitatory neurometabolites gradually decreased over the four-week training period, but only for complex but not simple motor training. Lastly, neurometabolites in left PMd showed no task-related modulation at the group level. However, the individual interplay between excitatory and inhibitory neurotransmitter modulation in left PMd during initial motor learning influenced short-term training success.

## Acknowledgements

This work was supported by the Research Fund KU Leuven (C16/15/070), the Research Foundation Flanders grant (G089818N, G039821N), the Excellence of Science grant (EOS 30446199, MEMODYN) and the Hercules fund AUHL/11/01 (R-3987) and I005018N. Melina Hehl is funded by a fellowship grant from Research Foundation Flanders (11F6921N) and a KU Leuven Special Research Fund (PDMT2/24/077). Shanti Van Malderen is funded by a fellowship grant from Research Foundation Flanders (11L9322N) and an UHasselt Special Research Fund (BOF21INCENT15). Svitlana Blashchuk is funded by an UHasselt Special Research Fund (BOF24DOC13). The authors declare no competing financial interests. The funders had no role in study design, data collection and analysis, decision to publish, or preparation of the manuscript.

## Author contributions

MH, KC and SPS designed the study; MH performed the data acquisition with help from SVM and KC; MH analyzed the data; MH wrote the manuscript with input from all authors (SVM, SB, SS, RAEE, SPS, KC).

## Data availability statement

The data that support the findings of this study are available on request from the corresponding author. The data are not publicly available due to privacy or ethical restrictions.

## Abbreviations

BLOCK_BTT1/BTT2: MRS acquisition in left PMd during execution of the BTT (first/last 11 min, respectively)
BLOCK_RESTafter: MRS acquisition in left PMd at rest after task execution
BLOCK_RESTbefore: MRS acquisition in left PMd at rest before task execution
BTT: bimanual tracking task
CSF: cerebrospinal fluid
E-I balance: excitatory-inhibitory balance
fMRI: functional magnetic resonance imaging
GABA: gamma-aminobutyric acid
GABA+: gamma-aminobutyric acid plus co-edited macromolecules
Glu: glutamate
Glx: compound measure of glutamate and glutamine
GM: gray matter
LMM: linear mixed model
LTD: long-term depression
LTP: long-term potentiation
M1: primary motor cortex
MEGA-PRESS: Mescher-Garwood Point Resolved Spectroscopy
MID: measurement session after two weeks of motor training
MOIST: Multiply Optimized Insensitive Suppression Train
MRI: magnetic resonance imaging
MRS: magnetic resonance spectroscopy
NM: neurometabolite
PET: positron emission tomography
PMd: dorsal premotor cortex
PMv: ventral premotor cortex
POST: measurement session after four weeks of motor learning
PRE: measurement session at baseline
PT: progress test
PT_BTT: progress tests of the bimanual tracking task
S1: primary somatosensory cortex
SM1: primary sensorimotor cortex
SMA: supplementary motor area
SNR: signal-to-noise ratio
SRTT: serial reaction time task
tDCS: transcranial direct current stimulation
TE: echo time
TMS: transcranial magnetic stimulation
TR: repetition time
VOI: volume of interest
WM: white matter

