## Appendix/Supplement for "The reciprocal relationship between short- and long-term motor learning and neurometabolites"

*(Running title: **Interplay of GABA, Glx & Motor Learning**)*

Melina Hehl[1,2,3,4], Shanti Van Malderen[1,2,3], Svitlana Blashchuk[3], Stefan Sunaert[4], Richard A. E. Edden[5,6], Stephan P. Swinnen[1,2], & Koen Cuypers[1,2,3]

[1]Movement Control & Neuroplasticity Research Group, Department of Movement Sciences, Group Biomedical Sciences, KU Leuven, Heverlee, Belgium

[2]KU Leuven, Leuven Brain Institute (LBI), Leuven, Belgium

[3]Neuroplasticity and Movement Control Research Group, Rehabilitation Research Institute (REVAL), Hasselt University, Diepenbeek, Belgium

[4]Department of Imaging and Pathology, Group Biomedical Sciences, KU Leuven, Leuven, Belgium

[5]Russell H. Morgan Department of Radiology and Radiological Science, The Johns Hopkins University School of Medicine, Baltimore, MD, USA

[6]F. M. Kirby Research Center for Functional Brain Imaging, Kennedy Krieger Institute, Baltimore, MD, USA

* Corresponding author:

Melina Hehl

Movement Control & Neuroplasticity Research Group,

Department of Movement Sciences, Group Biomedical Sciences,

KU Leuven,

Tervuursevest 101 – box 1501, 3001 Heverlee, Belgium.

**Appendix 1. Detailed description of the Bimanual Tracking Task (BTT)**

***The Bimanual Tracking Task***

The goal of the BTT was to accurately track a target moving along a target line presented on a screen with a cursor by rotating two dials to control the vertical and horizontal movements with the left and right index finger, respectively (see main manuscript **Figure 2B**) (Fujiyama et al., 2016; Sisti et al., 2011; Zivari Adab et al., 2020). Consequently, moving both dials at the exact same speed (frequency ratio L:R of 1:1, both hands rotating rightwards) would result in a line tilted 45° to the upper right corner, whereas other frequency ratios would lead to steeper lines (e.g., 3:1) or flatter lines (e.g., 1:3). The BTT was performed while participants were seated in an upright position in front of the task setup and a 15-inch laptop placed on a table in front of them (see main manuscript, **Figure 2A**). The task setup consisted of two hand rests spaced approximately 48 cm apart, and circular dials (diameter = 6 cm) in front of them with a hollow (diameter = 1.5 cm), in which participants placed the fingertip of their index fingers after adjusting the distance between handle and dial to each individual's hand size.

***The Training Paradigm***

The 4-week training paradigm was compromised of 4 training sessions per week lasting 30 minutes each, separated into 6 blocks of 5 minutes with a brief self-paced break between blocks to avoid fatigue. Trials lasted 9 to 54 sec consisting of 2 sec with display of the target line, 5–50 sec of movement execution (depending on the trial), and 2 sec in which the feedback was displayed. Trials were compiled in a way to always add up to 5-minute training blocks. After thorough instruction and a 2-minute familiarization session, participants performed the total of 16 training sessions in their home environment with daily online follow-up and occasional contact by phone to ensure training adherence. After screening, participants were stratified by sex and pseudo-randomly assigned to either the complex training group or the simple training group.

***Complex Training Group***

Participants in the complex training group would constantly be kept challenged throughout the training paradigm by a staircase-level-design with participants being exposed to progressively more difficult tasks with increasing level number (280 different tasks, combined and arranged to 5-minute levels in the coarse order: straight lines [in-phase → anti-phase] → simple curves → zig-zags → waves → complex figures with various combinations of the stated conditions; also the speed of the tracing would vary between trials). Starting in the first level with the simplest task variant (1:1 lines), participants would progress one level (>65%), stay on the same level (40–65%), or regress one level (<40%) based on the average score of all trials in the previous 5-minute block.

***Simple Training Group***

Participants in the simple training group were only exposed to the simplest task condition, the 1:1 frequency ratio line in all four possible directions (2 sec of preparation, 10 sec of movement, 2 sec of feedback), with both hands moving at the same speed. The plasticity induced by the complex bimanual task training program was compared with this practice of a simple task variant that is considered to be part of the intrinsic motor repertoire and does not require learning (control group).

***Progress Tests***

To ensure comparability of the training progress between individuals despite individualized training program and two different training paradigms (complex vs. simple), participants were presented with a 5-minute standardized progress test at six different times throughout the 4-week training paradigm: at baseline, at the beginning of each of the four training weeks (when it would replace the first 5-minute block of the training day) and after finalizing the training paradigm. This progress test consisted of a subset of trained tasks (1:1, 1:3, zig-zag, waves) and untrained tasks (complex line-art flamingo with several straight and bended lines) (see main manuscript, **Figure 2D**, as well as mirrored counterparts, total of 18 tasks). The first and the last progress test were performed under supervision in the laboratory.

***Score Calculation***

Participants' performance was scored on each trial with a percentage ranging from 0–100%, reflecting their overall accuracy in terms of speed, movement direction, and distance from the target. Participants received on-screen feedback of this score after each trial. The score was calculated online as described earlier (see Zivari Adab et al. (2020), their Fig. 1D, here with a trajectory sampling rate of 20 Hz) to obtain the preliminary percentage $P$, but was additionally multiplied with a distance factor $D$ to obtain the final score $S$. This was done to penalize perfectly parallel but distant tracking of the target line and therefore encourage participant's overall accuracy and offer more room for improvement. The distance factor $D$ was determined by calculating the distance between the participant's cursor and the target line for each data point (see Zivari Adab et al. (2020), their Fig. 1D) and averaging this distance over the whole trial as average distance $\bar{d}$ (expressed in units, with 1 unit = distance that can be covered in 200 ms). This average distance was then divided by 5 units (distance that can be covered in 1000 ms), a value that showed in pilot experiments to have the best sensitivity to progress while offering reasonably small bottom and ceiling effects, and subtracted from 1:

$$D = \left(1 - \frac{\bar{d}}{5}\right)$$

Finally, the distance factor $D$ was multiplied with the percentage of points $P$ that were reached in the correct order (see score calculation in see Zivari Adab et al. (2020), (Zivari Adab et al., 2020)(Zivari Adab et al., 2020)(Zivari Adab et al., 2020)(Zivari Adab et al., 2020)(Zivari Adab et al., 2020)(Zivari Adab et al., 2020)their Fig. 1D) to obtain the final score:

$$S = P \cdot D$$

Consequently, very small average distances $\bar{d}$ would lead to a distance factor $D$ close to 1, resulting to nearly no deduction from the obtained percentage $P$, whereas very large distances $\bar{d}$ would lead to a distance factor $D$ close to 0, and would therefore considerably lower the obtained percentage P. Lastly, since participants should not be discouraged during early learning, the final distance factor was limited to range between 1 (maximum) and 0.1 (minimum).

***Slope of Linear Regression as Overall Learning Outcome***

The learning progress was quantified as the linear slope of the progress test scores over time. This was done by estimating a linear regression using the average scores at each of the progress tests as dependent variable and the time spent on the BTT ($BTT_{time}$; during trainings and measurements, measured in minutes, see **Figure A1**) as the independent variable. Since participants in the here presented sample also participated in magnetic resonance imaging (MRI) and transcranial magnetic stimulation (TMS) experiments where they were presented with 1:1, 1:3 and 3:1 lines for functional measurements, exposure to the BTT during measurements has been included in the independent variable. An overview of the training weeks, points of measurement, training sessions (30 min each) and progress tests can be seen in **Figure A1**. The slope of a linear regression was chosen since it takes into account the initial task performance as well as the progress in performance as the training advances. Furthermore, the linear regression was chosen over a logarithmic or power regression due to its much better ability to fit the data of this learning process over an extended period of time. To obtain a more detailed picture of the skill level and coordinative requirements of different subsets of tasks, linear slopes were calculated for the average scores on specified sets of BTT subtasks (e.g., all lines: $BTTslope_{allLines}$ derived from $BTTscore_{allLines}$; complex task variants: $BTTslope_{complex}$ derived from $BTTscore_{complex}$).

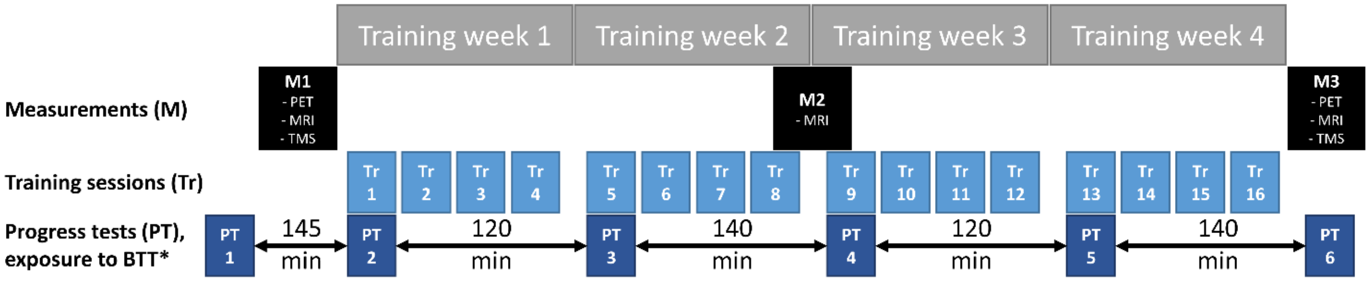

***Figure A1.*** *Overview of the four bimanual tracking task (BTT) training weeks. Each training session (Tr1–16) lasted 30 minutes, and trainings 1, 5, 9, and 13 started with a progress test instead of the first 5-minute block. During the measurements (M1–3), participants were exposed to the 1:1, 1:3 and 3:1 frequency ratio task variant during the magnetic resonance imaging (MRI, 20 min of BTT) and transcranial magnetic stimulation (TMS, 120 min of BTT), but not during the positron emission tomography (PET) measurement. Furthermore, participants were not shown any more difficult task variants during these times. Therefore, the exposure to the BTT in minutes is approximately equally spaced between progress tests (PT1–6).*

### Appendix 2. Detailed results of the behavioral analysis.

Appendix 2A: Behavioral analysis of PT_BTTscore$_{Lines}$ by time spent on the BTT.

#### Model 1: Full linear mixed model

Formula: PT_BTTScoreLines ~ BTTtime + GROUP + BTTtime * GROUP + (1 | SubjectID)

| AIC | BIC | logLik | deviance | df.resid |
|---|---|---|---|---|
| 2929.1 | 2952.6 | -1458.5 | 2917.1 | 366 |

Random effects:

| Groups | Name | Variance | Std.Dev. |
|---|---|---|---|
| **SubjectID** | (Intercept) | 68.13 | 8.254 |
| **Residual** | | 115.82 | 10.762 |

*Number of obs: 372, groups: SubjectID, 62*

Fixed effects:

| | Estimate | Std. Error | df | t-value | Pr(>\|t\|) | |
|---|---|---|---|---|---|---|
| **(Intercept)** | 32.1000 | 2.0820 | 131.00 | 15.414 | <0.0001 | *** |
| **BTTtime** | 0.0759 | 0.0036 | 310.00 | 21.260 | <0.0001 | *** |
| **GROUP[complex]** | 2.7690 | 2.8990 | 131.00 | 0.955 | 0.3410 | |
| **BTTtime:GROUP[complex]** | 0.0005 | 0.0050 | 310.00 | 0.103 | 0.9180 | |

*Signif. codes: 0 '***' 0.001 '**' 0.01 '*' 0.05 '.' 0.1 ' ' 1*

Type III Analysis of Variance Table with Satterthwaite's method:

| | Sum Sq | Mean Sq | NumDF | DenDF | F value | Pr(>F) | |
|---|---|---|---|---|---|---|---|
| **BTTtime** | 108802 | 108802 | 1 | 310.00 | 939.414 | <0.0001 | *** |
| **GROUP** | 106 | 106 | 1 | 131.04 | 0.912 | 0.3412 | |
| **BTTtime:GROUP** | 1 | 1 | 1 | 310.00 | 0.011 | 0.9178 | |

*Signif. codes: 0 '***' 0.001 '**' 0.01 '*' 0.05 '.' 0.1 ' ' 1*

Normality of residuals:

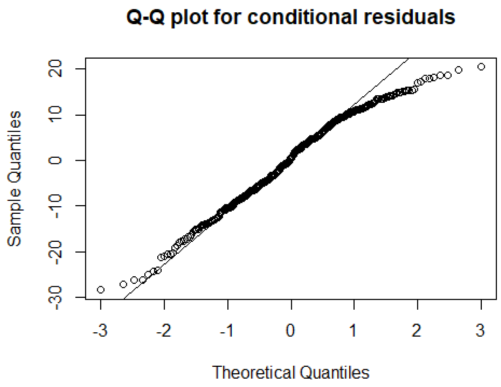

Shapiro-Wilk normality test: p = <0.0001

### Model 2: Full linear mixed model with cubic transformation

Formula: $(PT\_BTTScoreLines)^3$ ~ BTTtime + GROUP + BTTtime * GROUP + (1 | SubjectID)

| AIC | BIC | logLik | deviance | df.resid |
|---|---|---|---|---|
| 9521.2 | 9544.7 | -4754.6 | 9509.2 | 366 |

Random effects:

| Groups | Name | Variance | Std.Dev. |
|---|---|---|---|
| **SubjectID** | (Intercept) | $6252 \times 10^9$ | 79072.0 |
| **Residual** | | $5210 \times 10^9$ | 72180.0 |

*Number of obs: 372, groups: SubjectID, 62*

Fixed effects:

| | Estimate | Std. Error | df | t-value | Pr(>\|t\|) | |
|---|---|---|---|---|---|---|
| **(Intercept)** | 42546.13 | 17358.95 | 98.50 | 2.451 | 0.0160 | * |
| **BTTtime** | 653.41 | 23.94 | 310.00 | 27.299 | <0.0001 | *** |
| **GROUP[complex]** | 31787.72 | 24162.64 | 98.50 | 1.316 | 0.1910 | |
| **BTTtime:GROUP[complex]** | 14.14 | 33.32 | 310.00 | 0.425 | 0.6710 | |

*Signif. codes: 0 '***' 0.001 '**' 0.01 '*' 0.05 '.' 0.1 ' ' 1*

Type III Analysis of Variance Table with Satterthwaite's method:

| | Sum Sq | Mean Sq | NumDF | DenDF | F value | Pr(>F) | |
|---|---|---|---|---|---|---|---|
| **BTTtime** | $8.91 \times 10^{12}$ | $8.91 \times 10^{12}$ | 1 | 310.00 | 1571.981 | <0.0001 | *** |
| **GROUP** | $9.02 \times 10^9$ | $9.02 \times 10^9$ | 1 | 98.50 | 1.731 | 0.1914 | |
| **BTTtime:GROUP** | $9.39 \times 10^8$ | $9.39 \times 10^8$ | 1 | 310.00 | 0.180 | 0.6715 | |

*Signif. codes: 0 '***' 0.001 '**' 0.01 '*' 0.05 '.' 0.1 ' ' 1*

Normality of residuals:

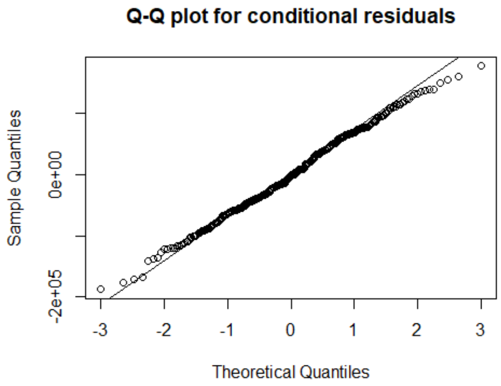

Shapiro-Wilk normality test: p = 0.3747

### Model 3: Removing BTTtime×GROUP interaction (based on p = 0.6715 in Model 2)

Formula: $(PT\_BTTScoreLines)^3$ ~ BTTtime + GROUP + (1 | SubjectID)

| AIC | BIC | logLik | deviance | df.resid |
|---|---|---|---|---|
| 9519.4 | 9539 | -4754.7 | 9509.4 | 367 |

Random effects:

| Groups | Name | Variance | Std.Dev. |
|---|---|---|---|
| **SubjectID** | (Intercept) | $6.25\times10^{9}$ | 79069 |
| **Residual** | | $5.21\times10^{9}$ | 72201 |

***Number of obs: 372, groups: SubjectID, 62***

Fixed effects:

| | Estimate | Std. Error | df | t-value | Pr(>\|t\|) | |
|---|---|---|---|---|---|---|
| **(Intercept)** | 40106.60 | 16380.82 | 78.97 | 2.448 | 0.0166 | * |
| **BTTtime** | 660.71 | 16.65 | 310.00 | 39.671 | <0.0001 | *** |
| **GROUP[complex]** | 36514.31 | 21444.81 | 62.00 | 1.703 | 0.0936 | . |

***Signif. codes: 0 '***' 0.001 '**' 0.01 '*' 0.05 '.' 0.1 ' ' 1***

Type III Analysis of Variance Table with Satterthwaite's method:

| | Sum Sq | Mean Sq | NumDF | DenDF | F value | Pr(>F) | |
|---|---|---|---|---|---|---|---|
| **BTTtime** | $8.20\times10^{12}$ | $8.20\times10^{12}$ | 1 | 310.00 | 1573.791 | <0.0001 | *** |
| **GROUP[complex]** | $1.51\times10^{10}$ | $1.51\times10^{10}$ | 1 | 62.00 | 2.899 | 0.0936 | . |

***Signif. codes: 0 '***' 0.001 '**' 0.01 '*' 0.05 '.' 0.1 ' ' 1***

### Model 4: Removing GROUP main effect (based on p = 0.09363 in Model 3) → **FINAL MODEL**

Formula: $(PT\_BTTScoreLines)^3$ ~ BTTtime + (1 | SubjectID)

| AIC | BIC | logLik | deviance | df.resid |
|---|---|---|---|---|
| 9520.2 | 9535.9 | -4756.1 | 9512.2 | 368 |

Random effects:

| Groups | Name | Variance | Std.Dev. |
|---|---|---|---|
| **SubjectID** | (Intercept) | $6.59\times10^{9}$ | 81147 |
| **Residual** | | $5.21\times10^{9}$ | 72201 |

*Number of obs: 372, groups: PatientID, 62*

Fixed effects:

| | Estimate | Std. Error | df | t-value | Pr(>\|t\|) | |
|---|---|---|---|---|---|---|
| **(Intercept)** | 58952.69 | 12296.14 | 96.78 | 4.794 | <0.0001 | *** |
| **BTTtime** | 660.71 | 16.65 | 310.00 | 39.671 | <0.0001 | *** |

*Signif. codes: 0 '***' 0.001 '**' 0.01 '*' 0.05 '.' 0.1 ' ' 1*

Type III Analysis of Variance Table with Satterthwaite's method:

| | Sum Sq | Mean Sq | NumDF | DenDF | F value | Pr(>F) | |
|---|---|---|---|---|---|---|---|
| **BTTtime** | $8.20{\times}10^{12}$ | $8.20{\times}10^{12}$ | 1 | 310.00 | 1573.8 | <0.0001 | *** |

*Signif. codes: 0 '***' 0.001 '**' 0.01 '*' 0.05 '.' 0.1 ' ' 1*

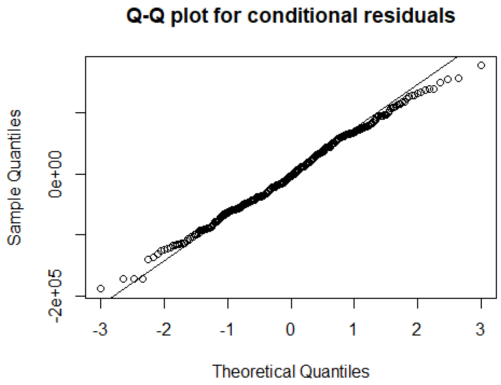

Shapiro-Wilk normality test: p = 0.334

Appendix 2B: Differences between subsequent progress tests for PT_BTTscore$_{Lines}$.

Results from paired one-sided t-tests with $H_A$: $\Delta\mu < 0$, with $\Delta\mu$ being the difference in mean score between the two progress tests (PTs). Significant results indicate an increase in performance from the earlier to the later PT. Fitting with the one-sided hypothesis, only an upper limit (UL) of the confidence interval (CI) is reported.

| contrast | mean difference | 95% CI UL | t-value | df | p-value | significance |
|---|---|---|---|---|---|---|
| **PT1 - PT2** | -16.49 | -13.74 | -10.038 | 61 | < 0.0001 | * |
| **PT2 - PT3** | -20.61 | -17.91 | -12.740 | 61 | < 0.0001 | * |
| **PT3 - PT4** | -7.98 | -6.61 | -9.725 | 61 | < 0.0001 | * |
| **PT4 - PT5** | -2.02 | -1.03 | -3.384 | 61 | 0.0006 | * |
| **PT5 - PT6** | -3.02 | -2.04 | -5.134 | 61 | < 0.0001 | * |

***Asterisks indicate results p < α, with Bonferroni correction for 5 tests (α = 0.05/5 = 0.01).***
***Abbreviations:** CI = confidence interval; df = degrees of freedom; PT = progress test; UL = upper limit.*

### Appendix 2C: Behavioral analysis of PT_BTTscore$_{Complex}$ by time spent on the BTT.

### Model 1: Full linear mixed model → **FINAL MODEL**

Formula: PT_BTTScoreComplex ~ BTTtime + GROUP + BTTtime * GROUP + (1 | SubjectID)

| AIC | BIC | logLik | deviance | df.resid |
|---|---|---|---|---|
| 2770.1 | 2793.6 | -1379 | 2758.1 | 366 |

Random effects:

| Groups | Name | Variance | Std.Dev. |
|---|---|---|---|
| **PatientID** | (Intercept) | 102.82 | 10.14 |
| **Residual** | | 65.78 | 8.11 |

*Number of obs: 372, groups: PatientID, 62*

Fixed effects:

| | Estimate | Std. Error | df | t-value | Pr(>\|t\|) | |
|---|---|---|---|---|---|---|
| **(Intercept)** | 4.7520 | 2.1450 | 90.40 | 2.216 | 0.0292 | * |
| **BTTtime** | 0.0618 | 0.0027 | 310.00 | 22.967 | <0.0001 | *** |
| **GROUP[complex]** | 0.6478 | 2.9860 | 90.40 | 0.217 | 0.8287 | |
| **BTTtime:GROUP[complex]** | 0.0363 | 0.0037 | 310.00 | 9.683 | <0.0001 | *** |

*Signif. codes: 0 '***' 0.001 '**' 0.01 '*' 0.05 '.' 0.1 ' ' 1*

Type III Analysis of Variance Table with Satterthwaite's method:

| | Sum Sq | Mean Sq | NumDF | DenDF | F value | Pr(>F) | |
|---|---|---|---|---|---|---|---|
| **BTTtime** | 119836 | 119836 | 1 | 310.00 | 1821.807 | <0.0001 | *** |
| **GROUP** | 3 | 3 | 1 | 90.40 | 0.047 | 0.8287 | |
| **BTTtime:GROUP** | 6168 | 6168 | 1 | 310.00 | 93.766 | <0.0001 | *** |

*Signif. codes: 0 '***' 0.001 '**' 0.01 '*' 0.05 '.' 0.1 ' ' 1*

Normality of residuals:

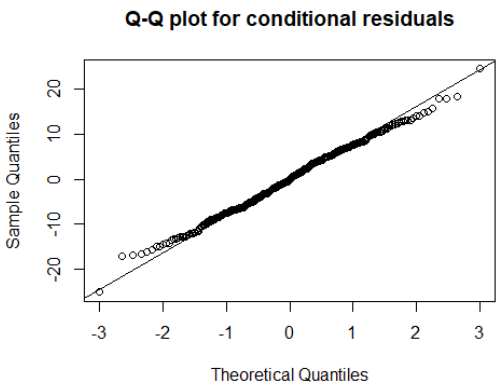

Shapiro-Wilk normality test: p = 0.4953

Appendix 2D: Differences between subsequent progress tests for PT_BTTscore$_{Complex}$, and differences in performance between the two training groups per progress test.

Due to the interaction with GROUP, both training groups have been analyzed individually, and progress test results have additionally been compared between groups.

Results from two-sided Welch two-sample t-tests with $H_A$: $\mu_{Simple} \neq \mu_{Complex}$, with μ being the mean score of the progress test (PTs) to test the group differences per PT.

| contrast | mean difference | 95% CI | t-value | df | p-value | significance |
|---|---|---|---|---|---|---|
| **(GROUP_simple - GROUP_complex) \| PT1** | -0.81 | -3.06 – 1.44 | -0.722 | 54.8 | 0.4733 | |
| **(GROUP_simple - GROUP_complex) \| PT2** | -2.11 | -6.55 – 2.34 | -0.950 | 57.4 | 0.3462 | |
| **(GROUP_simple - GROUP_complex) \| PT3** | -11.58 | -19.19 – -3.98 | -3.046 | 60.0 | 0.0034 | * |
| **(GROUP_simple - GROUP_complex) \| PT4** | -18.99 | -27.31 – -10.67 | -4.564 | 60.0 | < 0.0001 | * |
| **(GROUP_simple - GROUP_complex) \| PT5** | -22.41 | -30.08 – -14.73 | -5.850 | 56.2 | < 0.0001 | * |
| **(GROUP_simple - GROUP_complex) \| PT6** | -20.67 | -27.41 – -13.93 | -6.142 | 56.3 | < 0.0001 | * |

***Asterisks indicate results p < α, with Bonferroni correction for 5 tests (α = 0.05/6 = 0.0083).***
***Abbreviations:** CI = confidence interval; df = degrees of freedom; PT = progress test.*

Results from paired one-sided t-tests with $H_A$: $\Delta\mu < 0$, with Δμ being the difference in mean score between the two progress tests (PTs). Significant results indicate an increase in performance from the earlier to the later PT. Fitting with the one-sided hypothesis, only an upper limit (UL) of the confidence interval (CI) is reported.

| contrast | mean difference | 95% CI UL | t-value | df | p-value | significance |
|---|---|---|---|---|---|---|
| **(PT1 - PT2) \| GROUP_simple** | -5.03 | -3.14 | -4.526 | 29 | < 0.0001 | * |
| **(PT2 - PT3) \| GROUP_simple** | -10.11 | -7.23 | -5.971 | 29 | < 0.0001 | * |
| **(PT3 - PT4) \| GROUP_simple** | -8.96 | -6.73 | -6.843 | 29 | < 0.0001 | * |
| **(PT4 - PT5) \| GROUP_simple** | -7.74 | -5.88 | -7.082 | 29 | < 0.0001 | * |
| **(PT5 - PT6) \| GROUP_simple** | -7.28 | -5.12 | -5.744 | 29 | < 0.0001 | * |

***Asterisks indicate results p < α, with Bonferroni correction for 5 tests (α = 0.05/5 = 0.01).***
***Abbreviations:** CI = confidence interval; df = degrees of freedom; PT = progress test; UL = upper limit.*

| contrast | mean difference | 95% CI UL | t-value | df | p-value | significance |
|---|---|---|---|---|---|---|
| **(PT1 - PT2) \| GROUP_complex** | -6.33 | -4.65 | -6.405 | 31 | < 0.0001 | * |
| **(PT2 - PT3) \| GROUP_complex** | -19.59 | -16.30 | -10.089 | 31 | < 0.0001 | * |
| **(PT3 - PT4) \| GROUP_complex** | -16.36 | -14.23 | -13.023 | 31 | < 0.0001 | * |
| **(PT4 - PT5) \| GROUP_complex** | -11.15 | -8.75 | -7.869 | 31 | < 0.0001 | * |
| **(PT5 - PT6) \| GROUP_complex** | -5.54 | -4.11 | -6.562 | 31 | < 0.0001 | * |

***Asterisks indicate results p < α, with Bonferroni correction for 5 tests (α = 0.05/5 = 0.01).***
***Abbreviations:** CI = confidence interval; df = degrees of freedom; PT = progress test; UL = upper limit.*

Appendix 2E: Analysis of learning progress during the magnetic resonance spectroscopy scans.

Simple training group (one-sided paired t-test with $H_A$: $\mu_{\text{earlier session}} > \mu_{\text{later session}}$):

| Test (A - B) | t-value | df | p-value | A mean ± SD | B mean ± SD | 95% CI LL | Estimate difference |
|---|---|---|---|---|---|---|---|
| **MR_BTTslope$_{allLines}$** | | | | | | | |
| PRE - MID | 2.809 | 29 | **0.0044**** | 0.134 ± 0.091 | 0.108 ± 0.118 | 0.035 | 0.088 |
| MID - POST | 0.897 | 29 | 0.1885 | 0.046 ± 0.132 | 0.011 ± 0.082 | -0.022 | 0.025 |
| PRE - POST | 5.802 | 29 | **<0.0001**** | 0.020 ± 0.064 | -0.011 ± 0.078 | 0.080 | 0.114 |
| **MR_BTTslope$_{11}$** | | | | | | | |
| PRE - MID | 2.802 | 29 | **0.0045**** | 0.127 ± 0.139 | 0.094 ± 0.157 | 0.050 | 0.126 |
| MID - POST | -0.057 | 29 | 0.5226 | 0.001 ± 0.172 | 0.017 ± 0.090 | -0.060 | -0.002 |
| PRE - POST | 4.929 | 29 | **<0.0001**** | 0.003 ± 0.056 | -0.009 ± 0.069 | 0.081 | 0.124 |
| **MR_BTTslope$_{13,31}$** | | | | | | | |
| PRE - MID | 2.239 | 29 | **0.0165**** | 0.140 ± 0.091 | 0.117 ± 0.122 | 0.016 | 0.067 |
| MID - POST | 1.475 | 29 | 0.0754 | 0.073 ± 0.126 | 0.008 ± 0.085 | -0.006 | 0.041 |
| PRE - POST | 4.967 | 29 | **<0.0001**** | 0.032 ± 0.076 | -0.012 ± 0.089 | 0.071 | 0.108 |

*** indicate significant results surviving Bonferroni correction for 3 tests (α = 0.05/3 = 0.0167).*
*<u>Abbreviations:</u> CI = confidence interval; df = degrees of freedom; LL = lower limit; SD = standard deviation.*

Complex training group (one-sided paired t-test with $H_A$: $\mu_{\text{earlier session}} > \mu_{\text{later session}}$):

| Test (A - B) | t-value | df | p-value | A mean ± SD | B mean ± SD | 95% CI LL | Estimate difference |
|---|---|---|---|---|---|---|---|
| **MR_BTTslope$_{allLines}$** | | | | | | | |
| PRE - MID | 3.758 | 31 | **0.0004**** | 0.134 ± 0.091 | 0.108 ± 0.118 | 0.053 | 0.097 |
| MID - POST | 1.153 | 31 | 0.1288 | 0.046 ± 0.132 | 0.011 ± 0.082 | -0.010 | 0.022 |
| PRE - POST | 5.008 | 31 | **<0.0001**** | 0.020 ± 0.064 | -0.011 ± 0.078 | 0.079 | 0.119 |
| **MR_BTTslope$_{11}$** | | | | | | | |
| PRE - MID | 2.570 | 31 | **0.0076**** | 0.127 ± 0.139 | 0.094 ± 0.157 | 0.026 | 0.077 |
| MID - POST | 1.470 | 31 | 0.0758 | 0.001 ± 0.172 | 0.017 ± 0.090 | -0.004 | 0.026 |
| PRE - POST | 3.375 | 31 | **0.0010**** | 0.003 ± 0.056 | -0.009 ± 0.069 | 0.051 | 0.103 |
| **MR_BTTslope$_{13,31}$** | | | | | | | |
| PRE - MID | 3.897 | 31 | **0.0002**** | 0.140 ± 0.091 | 0.117 ± 0.122 | 0.061 | 0.108 |
| MID - POST | 0.949 | 31 | 0.1750 | 0.073 ± 0.126 | 0.008 ± 0.085 | -0.016 | 0.021 |
| PRE - POST | 5.231 | 31 | **<0.0001**** | 0.032 ± 0.076 | -0.012 ± 0.089 | 0.087 | 0.129 |

*** indicate significant results surviving Bonferroni correction for 3 tests (α = 0.05/3 = 0.0167).*
*<u>Abbreviations:</u> CI = confidence interval; df = degrees of freedom; SD = standard deviation.*

Simple (S) vs. complex (C) training group per session (independent Welch t-test with $H_A$: $\mu_{\text{Simple}} \neq \mu_{\text{Complex}}$):

| Test | t-value | df | p-value | S mean ± SD | C mean ± SD | 95% CI |
|---|---|---|---|---|---|---|
| **MR_BTTslope$_{allLines}$** | | | | | | |
| PRE (S - C) | 0.993 | 58 | 0.3250 | 0.134 ± 0.091 | 0.108 ± 0.118 | -0.027–0.080 |
| MID (S - C) | 1.236 | 48 | 0.2224 | 0.046 ± 0.132 | 0.011 ± 0.082 | -0.022–0.091 |
| POST (S - C) | 1.763 | 59 | 0.0831 | 0.020 ± 0.064 | -0.011 ± 0.078 | -0.004–0.068 |
| **MR_BTTslope$_{11}$** | | | | | | |
| PRE (S - C) | 0.876 | 60 | 0.3844 | 0.127 ± 0.139 | 0.094 ± 0.157 | -0.042–0.108 |
| MID (S - C) | -0.465 | 43 | 0.6441 | 0.001 ± 0.172 | 0.017 ± 0.090 | -0.087–0.055 |
| POST (S - C) | 0.727 | 59 | 0.4700 | 0.003 ± 0.056 | -0.009 ± 0.069 | -0.020–0.044 |
| **MR_BTTslope$_{13,31}$** | | | | | | |
| PRE (S - C) | 0.853 | 57 | 0.3974 | 0.140 ± 0.091 | 0.117 ± 0.122 | -0.031–0.078 |
| MID (S - C) | 2.344 | 50 | **0.0231*** | 0.073 ± 0.126 | 0.008 ± 0.085 | 0.009–0.119 |
| POST (S - C) | 2.102 | 60 | **0.0398*** | 0.032 ± 0.076 | -0.012 ± 0.089 | 0.002–0.086 |

** indicate results p < α = 0.05, not surviving Bonferroni correction for 3 tests (α = 0.05/3 = 0.0167).*
*<u>Abbreviations:</u> CI = confidence interval; df = degrees of freedom; SD = standard deviation.*

### Appendix 3. Analysis details regarding baseline neurometabolite levels to predict motor learning.

#### Appendix 3A: Multiple linear regression details for the prediction of long-term motor learning on simple tasks (PT_BTTslope$_{Lines}$) by GABA+ levels in motor-related brain regions.

#### Model 1: Full model multiple linear regression

Formula: PT_BTTslopeLines ~ GABA_R-SM1 + GABA_L-SM1 + GABA_L-PMd + GROUP + GABA_R-SM1:GROUP + GABA_L-SM1:GROUP + GABA_L-PMd:GROUP

Coefficients:

| | Estimate | Std. Error | t-value | Pr(>\|t\|) |
|---|---|---|---|---|
| **(Intercept)** | 0.097 | 0.061 | 1.598 | 0.1160 |
| **GABA_R-SM1** | -0.001 | 0.010 | -0.119 | 0.9050 |
| **GABA_L-SM1** | 0.000 | 0.010 | 0.034 | 0.9730 |
| **GABA_L-PMd** | -0.005 | 0.010 | -0.487 | 0.6280 |
| **GROUPcomp** | 0.072 | 0.086 | 0.833 | 0.4090 |
| **GABA_R-SM1:GROUPcomp** | 0.015 | 0.014 | 1.102 | 0.2750 |
| **GABA_L-SM1:GROUPcomp** | -0.013 | 0.013 | -0.976 | 0.3340 |
| **GABA_L-PMd:GROUPcomp** | -0.018 | 0.015 | -1.179 | 0.2440 |

***Residual standard error: 0.0211 on 53 degrees of freedom (1 observation deleted due to missingness)***
***Multiple R-squared: 0.1318, Adjusted R-squared: 0.01713***
***F-statistic: 1.149 on 7 and 53 DF, p-value: 0.3474***

Anova Table (Type III tests)

| | Sum Sq | DF | F value | Pr(>F) |
|---|---|---|---|---|
| **(Intercept)** | 0.0011365 | 1 | 2.5522 | 0.1161 |
| **GABA_R-SM1** | 0.0000063 | 1 | 0.0142 | 0.9055 |
| **GABA_L-SM1** | 0.0000005 | 1 | 0.0012 | 0.9726 |
| **GABA_L-PMd** | 0.0001055 | 1 | 0.2369 | 0.6284 |
| **GROUP** | 0.0003087 | 1 | 0.6932 | 0.4088 |
| **GABA_R-SM1:GROUP** | 0.0005408 | 1 | 1.2143 | 0.2755 |
| **GABA_L-SM1:GROUP** | 0.000424 | 1 | 0.9522 | 0.3336 |
| **GABA_L-PMd:GROUP** | 0.0006193 | 1 | 1.3906 | 0.2436 |
| **Residuals** | 0.0236022 | 53 | | |

*Signif. codes: 0 '***' 0.001 '**' 0.01 '*' 0.05 '.' 0.1 ' ' 1*

Check assumptions:

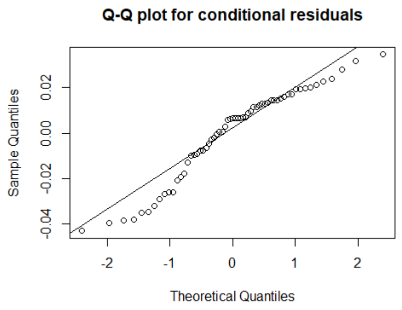

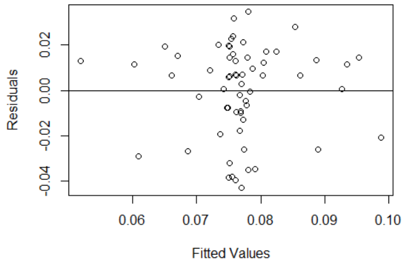

Shapiro-Wilk normality test: p = <0.0001

studentized Breusch-Pagan test: p = 0.5367

### Model 2: Full model multiple linear regression with cubic transformation

Formula: $(PT\_BTTslopeLines)^3$ ~ GABA_R-SM1 + GABA_L-SM1 + GABA_L-PMd + GROUP + GABA_R-SM1:GROUP + GABA_L-SM1:GROUP + GABA_L-PMd:GROUP

Coefficients:

| | Estimate | Std. Error | t-value | Pr(>\|t\|) |
|---|---|---|---|---|
| **(Intercept)** | 0.00077 | 0.00103 | 0.751 | 0.4560 |
| **GABA_R-SM1** | 0.00003 | 0.00017 | 0.191 | 0.8490 |
| **GABA_L-SM1** | 0.00003 | 0.00016 | 0.205 | 0.8390 |
| **GABA_L-PMd** | -0.00013 | 0.00016 | -0.778 | 0.4400 |
| **GROUPcomp** | 0.00173 | 0.00145 | 1.19 | 0.2390 |
| **GABA_R-SM1:GROUPcomp** | 0.00021 | 0.00023 | 0.891 | 0.3770 |
| **GABA_L-SM1:GROUPcomp** | -0.00029 | 0.00022 | -1.295 | 0.2010 |
| **GABA_L-PMd:GROUPcomp** | -0.00032 | 0.00026 | -1.208 | 0.2320 |

*Residual standard error: 0.0003559 on 53 degrees of freedom (1 observation deleted due to missingness)*
*Multiple R-squared: 0.1778, Adjusted R-squared: 0.06915*
*F-statistic: 1.637 on 7 and 53 DF, p-value: 0.1454*

Anova Table (Type III tests)

| | Sum Sq | DF | F value | Pr(>F) |
|---|---|---|---|---|
| **(Intercept)** | 7.1E-08 | 1 | 0.563 | 0.4563 |
| **GABA_R-SM1** | 4.6E-09 | 1 | 0.036 | 0.8494 |
| **GABA_L-SM1** | 5.3E-09 | 1 | 0.042 | 0.8385 |
| **GABA_L-PMd** | 7.7E-08 | 1 | 0.605 | 0.4403 |
| **GROUP** | 1.8E-07 | 1 | 1.416 | 0.2393 |
| **GABA_R-SM1:GROUP** | 1.0E-07 | 1 | 0.793 | 0.3772 |
| **GABA_L-SM1:GROUP** | 2.1E-07 | 1 | 1.677 | 0.2009 |
| **GABA_L-PMd:GROUP** | 1.9E-07 | 1 | 1.460 | 0.2323 |
| **Residuals** | 6.7E-06 | 53 | | |

*Signif. codes: 0 '***' 0.001 '**' 0.01 '*' 0.05 '.' 0.1 ' ' 1*

Check assumptions:

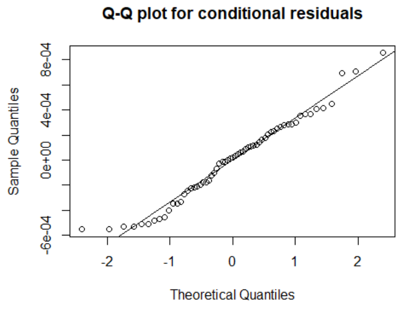

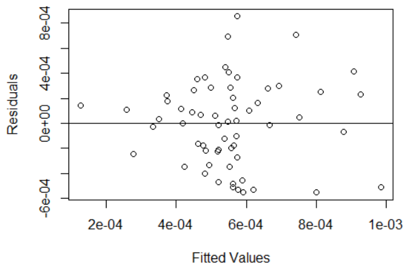

Shapiro-Wilk normality test: p = 0.1907

studentized Breusch-Pagan test: p = 0.4057

### Model 3: Removing GABA_R-SM1:GROUP interaction (based on p = 0.3772 in Model 2)

Formula: $(PT\_BTTslopeLines)^3$ ~ GABA_R-SM1 + GABA_L-SM1 + GABA_L-PMd + GROUP + GABA_L-SM1:GROUP + GABA_L-PMd:GROUP

Anova Table (Type III tests)

| | Sum Sq | DF | F value | Pr(>F) | |
|---|---|---|---|---|---|
| **(Intercept)** | 1.78E-08 | 1 | 0.1412 | 0.7086 | |
| **GABA_R-SM1** | 2.1E-07 | 1 | 1.656 | 0.2037 | |
| **GABA_L-SM1** | 3.5E-09 | 1 | 0.028 | 0.8689 | |
| **GABA_L-PMd** | 5.6E-08 | 1 | 0.441 | 0.5094 | |
| **GROUP** | 4.7E-07 | 1 | 3.716 | 0.0592 | . |
| **GABA_L-SM1:GROUP** | 1.6E-07 | 1 | 1.278 | 0.2633 | |
| **GABA_L-PMd:GROUP** | 2.4E-07 | 1 | 1.897 | 0.1741 | |
| **Residuals** | 6.8E-06 | 54 | | | |

*Signif. codes: 0 '***' 0.001 '**' 0.01 '*' 0.05 '.' 0.1 ' ' 1*

### Model 4: Removing GABA_L-SM1:GROUP interaction (based on p = 0.2633 in Model 3)

Formula: $(PT\_BTTslopeLines)^3$ ~ GABA_R-SM1 + GABA_L-SM1 + GABA_L-PMd + GROUP + GABA_L-PMd:GROUP

Anova Table (Type III tests)

| | Sum Sq | DF | F value | Pr(>F) |
|---|---|---|---|---|
| **(Intercept)** | 1.48E-07 | 1 | 1.165 | 0.2852 |
| **GABA_R-SM1** | 1.5E-07 | 1 | 1.201 | 0.2779 |
| **GABA_L-SM1** | 1.3E-07 | 1 | 1.020 | 0.3171 |
| **GABA_L-PMd** | 3.8E-08 | 1 | 0.302 | 0.5847 |
| **GROUP** | 3.1E-07 | 1 | 2.426 | 0.1251 |
| **GABA_L-PMd:GROUP** | 2.7E-07 | 1 | 2.158 | 0.1475 |
| **Residuals** | 7.0E-06 | 55 | | |

*Signif. codes: 0 '***' 0.001 '**' 0.01 '*' 0.05 '.' 0.1 ' ' 1*

### Model 5: Removing GABA_L-SM1 main effect (based on p = 0.3171 in Model 4)

Formula: $(PT\_BTTslopeLines)^3$ ~ GABA_R-SM1 + GABA_L-PMd + GROUP + GABA_L-PMd:GROUP

Anova Table (Type III tests)

| | Sum Sq | DF | F value | Pr(>F) |
|---|---|---|---|---|
| **(Intercept)** | 8.91E-08 | 1 | 0.702 | 0.4055 |
| **GABA_R-SM1** | 9.3E-08 | 1 | 0.735 | 0.3951 |
| **GABA_L-PMd** | 6.2E-08 | 1 | 0.490 | 0.4867 |
| **GROUP** | 2.8E-07 | 1 | 2.182 | 0.1452 |
| **GABA_L-PMd:GROUP** | 2.5E-07 | 1 | 1.977 | 0.1653 |
| **Residuals** | 7.1E-06 | 56 | | |

*Signif. codes: 0 '***' 0.001 '**' 0.01 '*' 0.05 '.' 0.1 ' ' 1*

### Model 6: Removing GABA_R-SM1 main effect (based on p = 0.3951 in Model 5)

Formula: $(PT\_BTTslopeLines)^3$ ~ GABA_R-SM1 + GABA_L-PMd + GROUP + GABA_L-PMd:GROUP

Anova Table (Type III tests)

| | Sum Sq | DF | F value | Pr(>F) |
|---|---|---|---|---|
| **(Intercept)** | 3.26E-07 | 1 | 2.526 | 0.1175 |
| **GABA_L-PMd** | 8.1E-08 | 1 | 0.629 | 0.4309 |
| **GROUP** | 2.8E-07 | 1 | 2.160 | 0.1471 |
| **GABA_L-PMd:GROUP** | 2.6E-07 | 1 | 1.999 | 0.1628 |
| **Residuals** | 7.5E-06 | 58 | | |

*Signif. codes: 0 '***' 0.001 '**' 0.01 '*' 0.05 '.' 0.1 ' ' 1*

### Model 7: Removing GABA_L-PMd:GROUP interaction (based on p = 0.1628 in Model 6)

Formula: $(PT\_BTTslopeLines)^3$ ~ GABA_R-SM1 + GABA_L-PMd + GROUP

Anova Table (Type III tests)

| | Sum Sq | DF | F value | Pr(>F) | |
|---|---|---|---|---|---|
| **(Intercept)** | 1.24E-06 | 1 | 9.462 | 0.0032 | ** |
| **GABA_L-PMd** | 5.7E-07 | 1 | 4.372 | 0.0408 | * |
| **GROUP** | 6.0E-08 | 1 | 0.456 | 0.5022 | |
| **Residuals** | 7.8E-06 | 59 | | | |

*Signif. codes: 0 '***' 0.001 '**' 0.01 '*' 0.05 '.' 0.1 ' ' 1*

### Model 8: Removing GROUP main effect (based on p = 0.5022 in Model 7) → **FINAL MODEL**

Formula: $(PT\_BTTslopeLines)^3$ ~ GABA_R-SM1 + GABA_L-PMd

Coefficients:

| | Estimate | Std. Error | t-value | Pr(>\|t\|) | |
|---|---|---|---|---|---|
| **(Intercept)** | 0.00160 | 0.00049 | 3.255 | 0.0019 | ** |
| **GABA_L-PMd** | -0.00027 | 0.00013 | -2.160 | 0.0348 | * |

*Residual standard error: 0.0003609 on 60 degrees of freedom*
*Multiple R-squared: 0.07217, Adjusted R-squared: 0.05671*
*F-statistic: 4.667 on 1 and 60 DF, p-value: 0.03475*

Anova Table (Type III tests)

| | Sum Sq | DF | F value | Pr(>F) | |
|---|---|---|---|---|---|
| **(Intercept)** | 1.38E-06 | 1 | 10.598 | 0.0019 | ** |
| **GABA_L-PMd** | 6.1E-07 | 1 | 4.667 | 0.0347 | * |
| **Residuals** | 7.8E-06 | 60 | | | |

*Signif. codes: 0 '***' 0.001 '**' 0.01 '*' 0.05 '.' 0.1 ' ' 1*

Check assumptions:

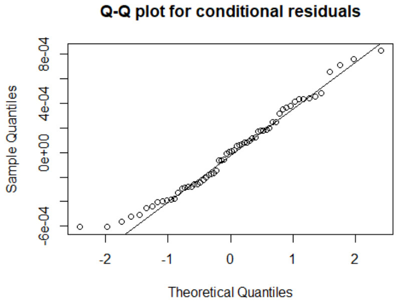

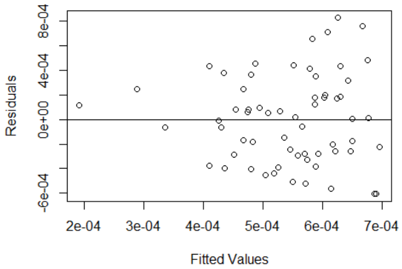

Shapiro-Wilk normality test: p = 0.2029

studentized Breusch-Pagan test: p = 0.0155

### Appendix 3B: Multiple linear regression details for the prediction of long-term motor learning on complex tasks (PT_BTTslope$_{Complex}$) by GABA+ levels in motor-related brain regions.

### Model 1: Full model multiple linear regression

Formula: PT_BTTslopeComplex ~ GABA_R-SM1 + GABA_L-SM1 + GABA_L-PMd + GROUP + GABA_R-SM1:GROUP + GABA_L-SM1:GROUP + GABA_L-PMd:GROUP

Coefficients:

| | Estimate | Std. Error | t-value | Pr(>\|t\|) |
|---|---|---|---|---|
| **(Intercept)** | 0.02253 | 0.06015 | 0.375 | 0.7090 |
| **GABA_R-SM1** | 0.00670 | 0.01008 | 0.664 | 0.5090 |
| **GABA_L-SM1** | 0.00710 | 0.00960 | 0.739 | 0.4630 |
| **GABA_L-PMd** | -0.00225 | 0.00952 | -0.237 | 0.8140 |
| **GROUPcomp** | 0.02181 | 0.08513 | 0.256 | 0.7990 |
| **GABA_R-SM1:GROUPcomp** | 0.01606 | 0.01351 | 1.189 | 0.2400 |
| **GABA_L-SM1:GROUPcomp** | -0.01229 | 0.01306 | -0.941 | 0.3510 |
| **GABA_L-PMd:GROUPcomp** | 0.00179 | 0.01530 | 0.117 | 0.9070 |

*Residual standard error: 0.02088 on 53 degrees of freedom (1 observation deleted due to missingness)*
*Multiple R-squared: 0.5096, Adjusted R-squared: 0.4449*
*F-statistic: 7.869 on 7 and 53 DF, p-value: 1.624e-06*

Anova Table (Type III tests)

| | Sum Sq | DF | F value | Pr(>F) |
|---|---|---|---|---|
| **(Intercept)** | 6.12E-05 | 1 | 0.140 | 0.7095 |
| **GABA_R-SM1** | 1.9E-04 | 1 | 0.441 | 0.5093 |
| **GABA_L-SM1** | 2.4E-04 | 1 | 0.546 | 0.4630 |
| **GABA_L-PMd** | 2.4E-05 | 1 | 0.056 | 0.8139 |
| **GROUP** | 2.9E-05 | 1 | 0.066 | 0.7988 |
| **GABA_R-SM1:GROUP** | 6.2E-04 | 1 | 1.413 | 0.2398 |
| **GABA_L-SM1:GROUP** | 3.9E-04 | 1 | 0.886 | 0.3508 |
| **GABA_L-PMd:GROUP** | 6.0E-06 | 1 | 0.014 | 0.9074 |
| **Residuals** | 2.3E-02 | 53 | | |

*Signif. codes: 0 ‘***’ 0.001 ‘**’ 0.01 ‘*’ 0.05 ‘.’ 0.1 ‘ ’ 1*

Check assumptions:

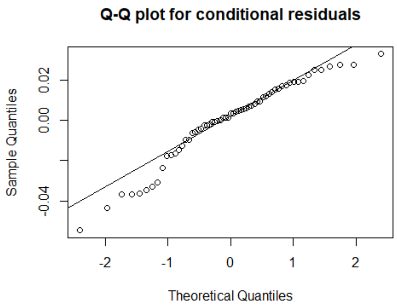

Shapiro-Wilk normality test: p = 0.0097

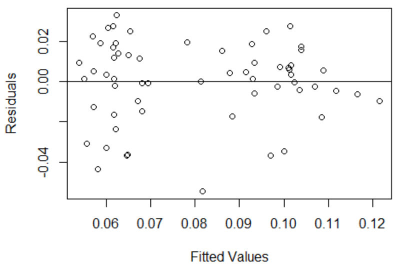

studentized Breusch-Pagan test: p = 0.3352

### Model 2: Full model multiple linear regression with quadratic transformation

Formula: $(PT\_BTTslopeComplex)^2$ ~ GABA_R-SM1 + GABA_L-SM1 + GABA_L-PMd + GROUP + GABA_R-SM1:GROUP + GABA_L-SM1:GROUP + GABA_L-PMd:GROUP

Coefficients:

| | Estimate | Std. Error | t-value | Pr(>\|t\|) | |
|---|---|---|---|---|---|
| **(Intercept)** | 0.00177 | 0.00835 | 0.212 | 0.8331 | |
| **GABA_R-SM1** | 0.00057 | 0.00140 | 0.406 | 0.6865 | |
| **GABA_L-SM1** | 0.00076 | 0.00133 | 0.572 | 0.5700 | |
| **GABA_L-PMd** | -0.00055 | 0.00132 | -0.414 | 0.6809 | |
| **GROUPcomp** | -0.00189 | 0.01181 | -0.16 | 0.8732 | |
| **GABA_R-SM1:GROUPcomp** | 0.00321 | 0.00188 | 1.71 | 0.0931 | . |
| **GABA_L-SM1:GROUPcomp** | -0.00124 | 0.00181 | -0.682 | 0.4984 | |
| **GABA_L-PMd:GROUPcomp** | 0.00041 | 0.00212 | 0.193 | 0.8475 | |

*Signif. codes: 0 '***' 0.001 '**' 0.01 '*' 0.05 '.' 0.1 ' ' 1*
*Residual standard error: 0.002898 on 53 degrees of freedom (1 observation deleted due to missingness)*
*Multiple R-squared: 0.5784, Adjusted R-squared: 0.5227*
*F-statistic: 10.39 on 7 and 53 DF, p-value: 3.98e-08*

Anova Table (Type III tests)

| | Sum Sq | DF | F value | Pr(>F) | |
|---|---|---|---|---|---|
| **(Intercept)** | 3.80E-07 | 1 | 0.045 | 0.8331 | |
| **GABA_R-SM1** | 1.4E-06 | 1 | 0.165 | 0.6865 | |
| **GABA_L-SM1** | 2.7E-06 | 1 | 0.327 | 0.5700 | |
| **GABA_L-PMd** | 1.4E-06 | 1 | 0.171 | 0.68088 | |
| **GROUP** | 2.2E-07 | 1 | 0.026 | 0.87317 | |
| **GABA_R-SM1:GROUP** | 2.5E-05 | 1 | 2.924 | 0.09313 | . |
| **GABA_L-SM1:GROUP** | 3.9E-06 | 1 | 0.465 | 0.49835 | |
| **GABA_L-PMd:GROUP** | 3.1E-07 | 1 | 0.037 | 0.84754 | |
| **Residuals** | 4.5E-04 | 53 | | | |

*Signif. codes: 0 '***' 0.001 '**' 0.01 '*' 0.05 '.' 0.1 ' ' 1*

Check assumptions:

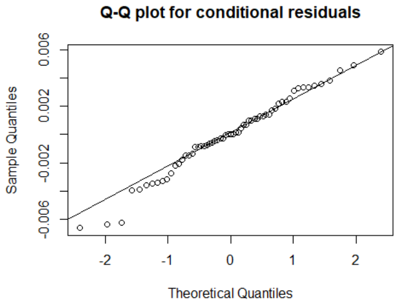

Shapiro-Wilk normality test: p = 0.5039

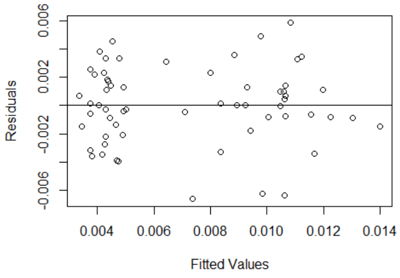

studentized Breusch-Pagan test: p = 0.3804

### Model 3: Removing GABA_L-PMd:GROUP interaction (based on p = 0.8475 in Model 2)

Formula: $(PT\_BTTslopeComplex)^2$ ~ GABA_R-SM1 + GABA_L-SM1 + GABA_L-PMd + GROUP + GABA_R-SM1:GROUP + GABA_L-SM1:GROUP

Anova Table (Type III tests)

| | Sum Sq | DF | F value | Pr(>F) | |
|---|---|---|---|---|---|
| **(Intercept)** | 1.90E-07 | 1 | 0.023 | 0.8814 | |
| **GABA_R-SM1** | 1.6E-06 | 1 | 0.189 | 0.6658 | |
| **GABA_L-SM1** | 2.6E-06 | 1 | 0.312 | 0.5786 | |
| **GABA_L-PMd** | 1.2E-06 | 1 | 0.143 | 0.7069 | |
| **GROUP** | 1.0E-08 | 1 | 0.001 | 0.9762 | |
| **GABA_R-SM1:GROUP** | 2.4E-05 | 1 | 2.947 | 0.0918 | . |
| **GABA_L-SM1:GROUP** | 3.7E-06 | 1 | 0.449 | 0.5056 | |
| **Residuals** | 4.5E-04 | 54 | | | |

*Signif. codes: 0 '***' 0.001 '**' 0.01 '*' 0.05 '.' 0.1 ' ' 1*

### Model 4: Removing GABA_L-PMd main effect (based on p = 0.7069 in Model 3)

Formula: $(PT\_BTTslopeComplex)^2$ ~ GABA_R-SM1 + GABA_L-SM1 + GROUP + GABA_R-SM1:GROUP + GABA_L-SM1:GROUP

Anova Table (Type III tests)

| | Sum Sq | DF | F value | Pr(>F) | |
|---|---|---|---|---|---|
| **(Intercept)** | 3.00E-08 | 1 | 0.004 | 0.9533 | |
| **GABA_R-SM1** | 2.0E-06 | 1 | 0.248 | 0.6208 | |
| **GABA_L-SM1** | 2.2E-06 | 1 | 0.266 | 0.6081 | |
| **GROUP** | 1.0E-08 | 1 | 0.002 | 0.9696 | |
| **GABA_R-SM1:GROUP** | 2.4E-05 | 1 | 2.944 | 0.0919 | . |
| **GABA_L-SM1:GROUP** | 3.4E-06 | 1 | 0.421 | 0.5192 | |
| **Residuals** | 4.5E-04 | 55 | | | |

*Signif. codes: 0 '***' 0.001 '**' 0.01 '*' 0.05 '.' 0.1 ' ' 1*

### Model 5: Removing GABA_L-SM1:GROUP interaction (based on p = 0.5192 in Model 4)

Formula: $(PT\_BTTslopeComplex)^2$ ~ GABA_R-SM1 + GABA_L-SM1 + GROUP + GABA_R-SM1:GROUP

Anova Table (Type III tests)

| | Sum Sq | DF | F value | Pr(>F) |
|---|---|---|---|---|
| **(Intercept)** | 8.80E-07 | 1 | 0.109 | 0.7421 |
| **GABA_R-SM1** | 2.1E-06 | 1 | 0.264 | 0.6094 |
| **GABA_L-SM1** | 3.0E-08 | 1 | 0.004 | 0.9515 |
| **GROUP** | 3.4E-06 | 1 | 0.419 | 0.5202 |
| **GABA_R-SM1:GROUP** | 2.1E-05 | 1 | 2.647 | 0.1094 |
| **Residuals** | 4.5E-04 | 56 | | |

*Signif. codes: 0 '***' 0.001 '**' 0.01 '*' 0.05 '.' 0.1 ' ' 1*

### Model 6: Removing GABA_L-SM1 main effect (based on p = 0.9515 in Model 5)

Formula: $(PT\_BTTslopeComplex)^2$ ~ GABA_R-SM1 + GROUP + GABA_R-SM1:GROUP

Anova Table (Type III tests)

| | Sum Sq | DF | F value | Pr(>F) | |
|---|---|---|---|---|---|
| **(Intercept)** | 1.60E-06 | 1 | 0.203 | 0.6544 | |
| **GABA_R-SM1** | 2.1E-06 | 1 | 0.270 | 0.6053 | |
| **GROUP** | 3.6E-06 | 1 | 0.451 | 0.5046 | |
| **GABA_R-SM1:GROUP** | 2.2E-05 | 1 | 2.832 | 0.0979 | . |
| **Residuals** | 4.5E-04 | 57 | | | |

*Signif. codes: 0 '***' 0.001 '**' 0.01 '*' 0.05 '.' 0.1 ' ' 1*

### Model 7: Removing GABA_R-SM1:GROUP interaction (based on p = 0.0979 in Model 6) → **FINAL MODEL**

Formula: $(PT\_BTTslopeComplex)^2$ ~ GABA_R-SM1 + GROUP

Coefficients:

| | Estimate | Std. Error | t-value | Pr(>\|t\|) | |
|---|---|---|---|---|---|
| **(Intercept)** | -0.00373 | 0.00288 | -1.296 | 0.2002 | |
| **GABA_R-SM1** | 0.00243 | 0.00086 | 2.828 | 0.0064 | ** |
| **GROUPcomp** | 0.00575 | 0.00073 | 7.856 | <0.0001 | *** |

*Signif. codes: 0 '***' 0.001 '**' 0.01 '*' 0.05 '.' 0.1 ' ' 1*
*Residual standard error: 0.002854 on 58 degrees of freedom (1 observation deleted due to missingness)*
*Multiple R-squared: 0.5525, Adjusted R-squared: 0.5371*
*F-statistic: 35.81 on 2 and 58 DF, p-value: 7.441e-11*

Anova Table (Type III tests)

| | Sum Sq | DF | F value | Pr(>F) | |
|---|---|---|---|---|---|
| **(Intercept)** | 1.37E-05 | 1 | 1.679 | 0.2002 | |
| **GABA_R-SM1** | 6.5E-05 | 1 | 7.999 | 0.0064 | ** |
| **GROUP** | 5.0E-04 | 1 | 61.710 | <0.0001 | *** |
| **Residuals** | 4.7E-04 | 58 | | | |

*Signif. codes: 0 '***' 0.001 '**' 0.01 '*' 0.05 '.' 0.1 ' ' 1*

Check assumptions:

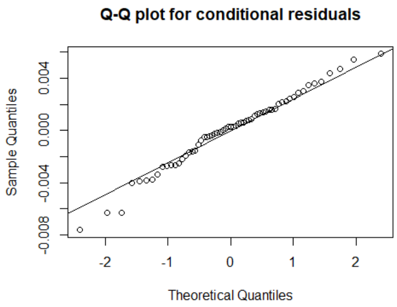

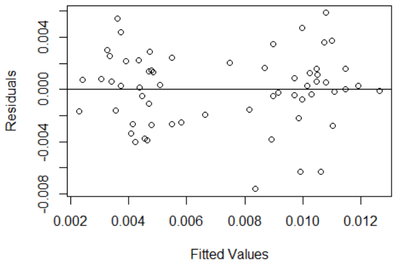

Shapiro-Wilk normality test: p = 0.2029

studentized Breusch-Pagan test: p = 0.3899

### Appendix 3C: Multiple linear regression details for the prediction of long-term motor learning on complex tasks ($PT\_BTTslope_{Complex}$) by Glx levels in motor-related brain regions.

### Model 1: Full model multiple linear regression

Formula: PT_BTTslopeComplex ~ Glx_R-SM1 + Glx_L-SM1 + Glx_L-PMd + GROUP + Glx_R-SM1:GROUP + Glx_L-SM1:GROUP + Glx_L-PMd:GROUP

Coefficients:

| | Estimate | Std. Error | t-value | Pr(>\|t\|) | |
|---|---|---|---|---|---|
| **(Intercept)** | -0.05263 | 0.08906 | -0.591 | 0.5571 | |
| **Glx_R-SM1** | -0.00094 | 0.00366 | -0.258 | 0.7976 | |
| **Glx_L-SM1** | 0.00575 | 0.00426 | 1.349 | 0.1829 | |
| **Glx_L-PMd** | 0.00561 | 0.00322 | 1.743 | 0.0871 | . |
| **GROUPcomp** | 0.16797 | 0.11757 | 1.429 | 0.1590 | |
| **Glx_R-SM1:GROUPcomp** | 0.00075 | 0.00551 | 0.137 | 0.8919 | |
| **Glx_L-SM1:GROUPcomp** | -0.00915 | 0.00549 | -1.666 | 0.1015 | |
| **Glx_L-PMd:GROUPcomp** | -0.00356 | 0.00446 | -0.798 | 0.4282 | |

*Signif. codes: 0 '***' 0.001 '**' 0.01 '*' 0.05 '.' 0.1 ' ' 1*
*Residual standard error: 0.02093 on 53 degrees of freedom (1 observation deleted due to missingness)*
*Multiple R-squared: 0.5076, Adjusted R-squared: 0.4426*
*F-statistic: 7.805 on 7 and 53 DF, p-value: 1.796e-06*

Anova Table (Type III tests)

| | Sum Sq | DF | F value | Pr(>F) | |
|---|---|---|---|---|---|
| **(Intercept)** | 1.53E-04 | 1 | 0.349 | 0.5571 | |
| **Glx_R-SM1** | 2.9E-05 | 1 | 0.066 | 0.7976 | |
| **Glx_L-SM1** | 8.0E-04 | 1 | 1.821 | 0.1829 | |
| **Glx_L-PMd** | 1.3E-03 | 1 | 3.039 | 0.0871 | . |
| **GROUP** | 8.9E-04 | 1 | 2.041 | 0.1590 | |
| **Glx_R-SM1:GROUP** | 8.2E-06 | 1 | 0.019 | 0.8919 | |
| **Glx_L-SM1:GROUP** | 1.2E-03 | 1 | 2.777 | 0.1016 | |
| **Glx_L-PMd:GROUP** | 2.8E-04 | 1 | 0.637 | 0.4282 | |
| **Residuals** | 2.3E-02 | 53 | | | |

*Signif. codes: 0 '***' 0.001 '**' 0.01 '*' 0.05 '.' 0.1 ' ' 1*

Check assumptions:

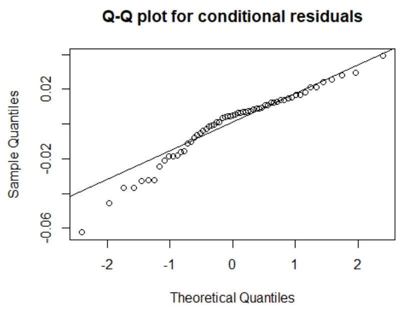

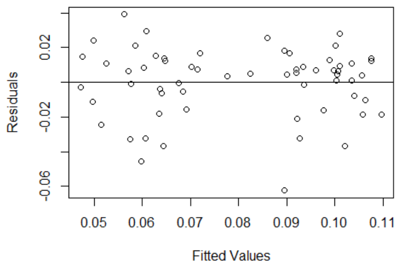

Shapiro-Wilk normality test: p = 0.0072

studentized Breusch-Pagan test: p = 0.4224

### Model 2: Full model multiple linear regression with quadratic transformation

Formula: $(PT\_BTTslopeComplex)^2$ ~ Glx_R-SM1 + Glx_L-SM1 + Glx_L-PMd + GROUP + Glx_R-SM1:GROUP + Glx_L-SM1:GROUP + Glx_L-PMd:GROUP

Coefficients:

| | Estimate | Std. Error | t-value | Pr(>\|t\|) |
|---|---|---|---|---|
| **(Intercept)** | -0.00670 | 0.01270 | -0.527 | 0.6000 |
| **Glx_R-SM1** | -0.00028 | 0.00052 | -0.53 | 0.5980 |
| **Glx_L-SM1** | 0.00070 | 0.00061 | 1.151 | 0.2550 |
| **Glx_L-PMd** | 0.00059 | 0.00046 | 1.281 | 0.2060 |
| **GROUPcomp** | 0.01724 | 0.01677 | 1.028 | 0.3090 |
| **Glx_R-SM1:GROUPcomp** | 0.00040 | 0.00079 | 0.51 | 0.6120 |
| **Glx_L-SM1:GROUPcomp** | -0.00129 | 0.00078 | -1.648 | 0.1050 |
| **Glx_L-PMd:GROUPcomp** | -0.00017 | 0.00064 | -0.269 | 0.7890 |

*Residual standard error: 0.002985 on 53 degrees of freedom (1 observation deleted due to missingness)*
*Multiple R-squared:  0.5527, Adjusted R-squared: 0.4936*
*F-statistic: 9.354 on 7 and 53 DF, p-value: 1.722e-07*

Anova Table (Type III tests)

| | Sum Sq | DF | F value | Pr(>F) |
|---|---|---|---|---|
| **(Intercept)** | 2.48E-06 | 1 | 0.278 | 0.6002 |
| **Glx_R-SM1** | 2.5E-06 | 1 | 0.281 | 0.5984 |
| **Glx_L-SM1** | 1.2E-05 | 1 | 1.325 | 0.2549 |
| **Glx_L-PMd** | 1.5E-05 | 1 | 1.640 | 0.2058 |
| **GROUP** | 9.4E-06 | 1 | 1.057 | 0.3086 |
| **Glx_R-SM1:GROUP** | 2.3E-06 | 1 | 0.260 | 0.6124 |
| **Glx_L-SM1:GROUP** | 2.4E-05 | 1 | 2.718 | 0.1052 |
| **Glx_L-PMd:GROUP** | 6.4E-07 | 1 | 0.072 | 0.7893 |
| **Residuals** | 4.7E-04 | 53 | | |

Check assumptions:

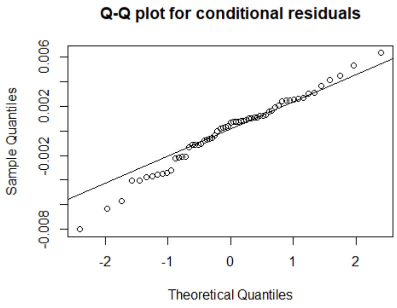

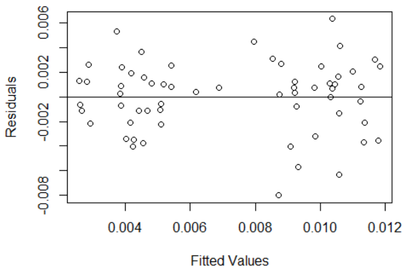

Shapiro-Wilk normality test: p = 0.3335

studentized Breusch-Pagan test: p = 0.3711

### Model 3: Removing Glx_L-PMd:GROUP interaction (based on p = 0.7893 in Model 2)

Formula: $(PT\_BTTslopeComplex)^2$ ~ Glx_R-SM1 + Glx_L-SM1 + Glx_L-PMd + GROUP + Glx_R-SM1:GROUP + Glx_L-SM1:GROUP

Anova Table (Type III tests)

| | Sum Sq | DF | F value | Pr(>F) |
|---|---|---|---|---|
| **(Intercept)** | 1.91E-06 | 1 | 0.218 | 0.6428 |
| **Glx_R-SM1** | 4.1E-06 | 1 | 0.467 | 0.4973 |
| **Glx_L-SM1** | 1.1E-05 | 1 | 1.288 | 0.2615 |
| **Glx_L-PMd** | 2.2E-05 | 1 | 2.508 | 0.1191 |
| **GROUP** | 2.3E-05 | 1 | 2.679 | 0.1075 |
| **Glx_R-SM1:GROUP** | 5.9E-06 | 1 | 0.674 | 0.4154 |
| **Glx_L-SM1:GROUP** | 2.4E-05 | 1 | 2.716 | 0.1052 |
| **Residuals** | 4.7E-04 | 54 | | |

*Signif. codes:  0 '***' 0.001 '**' 0.01 '*' 0.05 '.' 0.1 ' ' 1*

### Model 4: Removing Glx_R-SM1:GROUP interaction (based on p = 0.4154 in Model 3)

Formula: $(PT\_BTTslopeComplex)^2$ ~ Glx_R-SM1 + Glx_L-SM1 + Glx_L-PMd + GROUP + Glx_L-SM1:GROUP

Anova Table (Type III tests)

| | Sum Sq | DF | F value | Pr(>F) | |
|---|---|---|---|---|---|
| **(Intercept)** | 2.54E-06 | 1 | 0.292 | 0.5911 | |
| **Glx_R-SM1** | 5.2E-07 | 1 | 0.060 | 0.8081 | |
| **Glx_L-SM1** | 7.4E-06 | 1 | 0.848 | 0.3612 | |
| **Glx_L-PMd** | 1.9E-05 | 1 | 2.225 | 0.1415 | |
| **GROUP** | 4.3E-05 | 1 | 4.916 | 0.0308 | * |
| **Glx_L-SM1:GROUP** | 1.8E-05 | 1 | 2.060 | 0.1569 | |
| **Residuals** | 4.8E-04 | 55 | | | |

*Signif. codes:  0 '***' 0.001 '**' 0.01 '*' 0.05 '.' 0.1 ' ' 1*

### Model 5: Removing Glx_R-SM1 main effect (based on p = 0.8081 in Model 4)

Formula: $(PT\_BTTslopeComplex)^2$ ~ Glx_L-SM1 + Glx_L-PMd + GROUP + Glx_L-SM1:GROUP

Anova Table (Type III tests)

| | Sum Sq | DF | F value | Pr(>F) | |
|---|---|---|---|---|---|
| **(Intercept)** | 7.04E-06 | 1 | 0.822 | 0.3684 | |
| **Glx_L-SM1** | 7.2E-06 | 1 | 0.836 | 0.3646 | |
| **Glx_L-PMd** | 3.5E-05 | 1 | 4.081 | 0.0482 | * |
| **GROUP** | 4.5E-05 | 1 | 5.227 | 0.0261 | * |
| **Glx_L-SM1:GROUP** | 1.9E-05 | 1 | 2.217 | 0.1421 | |
| **Residuals** | 4.8E-04 | 56 | | | |

*Signif. codes:  0 '***' 0.001 '**' 0.01 '*' 0.05 '.' 0.1 ' ' 1*

### Model 6: Removing Glx_L-SM1:GROUP interaction (based on p = 0.1421 in Model 5)

Formula: $(PT\_BTTslopeComplex)^2$ ~ Glx_L-SM1 + Glx_L-PMd + GROUP

Anova Table (Type III tests)

| | Sum Sq | DF | F value | Pr(>F) | |
|---|---|---|---|---|---|
| **(Intercept)** | 3.00E-08 | 1 | 0.003 | 0.9566 | |
| **Glx_L-SM1** | 6.6E-07 | 1 | 0.075 | 0.7846 | |
| **Glx_L-PMd** | 2.7E-05 | 1 | 3.101 | 0.0836 | . |
| **GROUP** | 4.6E-04 | 1 | 52.473 | <0.0001 | *** |
| **Residuals** | 5.0E-04 | 57 | | | |

*Signif. codes: 0 '***' 0.001 '**' 0.01 '*' 0.05 '.' 0.1 ' ' 1*

### Model 7: Removing Glx_L-SM1 main effect (based on p = 0.7846 in Model 6) → **FINAL MODEL**

Formula: $(PT\_BTTslopeComplex)^2$ ~ Glx_L-PMd + GROUP

Coefficients:

| | Estimate | Std. Error | t-value | Pr(>\|t\|) | |
|---|---|---|---|---|---|
| **(Intercept)** | -0.00141 | 0.00250 | -0.563 | 0.5753 | |
| **Glx_L-PMd** | 0.00050 | 0.00021 | 2.328 | 0.0234 | * |
| **GROUPcomp** | 0.00567 | 0.00074 | 7.655 | 0.0000 | *** |

*Signif. codes: 0 '***' 0.001 '**' 0.01 '*' 0.05 '.' 0.1 ' ' 1*
*Residual standard error: 0.002915 on 59 degrees of freedom*
*Multiple R-squared: 0.5253, Adjusted R-squared: 0.5092*
*F-statistic: 32.64 on 2 and 59 DF, p-value: 2.854e-10*

Anova Table (Type III tests)

| | Sum Sq | DF | F value | Pr(>F) | |
|---|---|---|---|---|---|
| **(Intercept)** | 2.70E-06 | 1 | 0.317 | 0.5753 | |
| **Glx_L-PMd** | 4.6E-05 | 1 | 5.419 | 0.0234 | * |
| **GROUP** | 5.0E-04 | 1 | 58.594 | 0.0000 | *** |
| **Residuals** | 5.0E-04 | 59 | | | |

*Signif. codes: 0 '***' 0.001 '**' 0.01 '*' 0.05 '.' 0.1 ' ' 1*

Check assumptions:

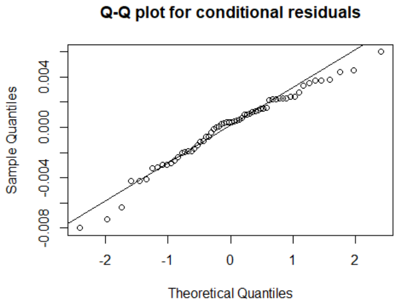

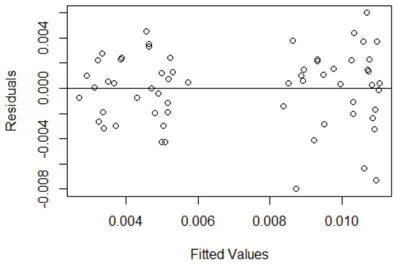

Shapiro-Wilk normality test: p = 0.2029

studentized Breusch-Pagan test: p = 0.2254

### Appendix 4. Analysis details regarding changes in resting neurometabolite levels with motor learning.

Appendix 4A: Linear mixed model of motor learning-related changes in resting Glx levels of the left SM1.

#### Model 1: Full linear mixed model

Formula: Glx_L-SM1 ~ GROUP + SESSION + GROUP:SESSION + (1 | SubjectID)

| AIC | BIC | logLik | deviance | df.resid |
|---|---|---|---|---|
| 560.1 | 585.9 | -272 | 544.1 | 177 |

Random effects:

| Groups | Name | Variance | Std.Dev. |
|---|---|---|---|
| **SubjectID** | (Intercept) | 0.586 | 0.765 |
| **Residual** | | 0.738 | 0.859 |

*Number of obs: 185, groups: SubjectID, 62*

Fixed effects:

| | Estimate | Std. Error | df | t-value | Pr(>\|t\|) | |
|---|---|---|---|---|---|---|
| **(Intercept)** | 10.662 | 0.210 | 133.09 | 50.751 | <0.0001 | *** |
| **GROUP[comp]** | 0.656 | 0.294 | 134.52 | 2.232 | 0.0273 | * |
| **SESSION[MID]** | 0.397 | 0.222 | 122.91 | 1.789 | 0.0762 | . |
| **SESSION[POST]** | 0.367 | 0.222 | 122.91 | 1.655 | 0.1005 | |
| **GROUP[comp]:SESSION[MID]** | -0.604 | 0.310 | 123.24 | -1.945 | 0.0540 | . |
| **GROUP[comp]:SESSION[POST]** | -0.947 | 0.310 | 123.24 | -3.053 | 0.0028 | ** |

*Signif. codes: 0 '***' 0.001 '**' 0.01 '*' 0.05 '.' 0.1 ' ' 1*

Type III Analysis of Variance Table with Satterthwaite's method:

| | Sum Sq | Mean Sq | NumDF | DenDF | F value | Pr(>F) | |
|---|---|---|---|---|---|---|---|
| **GROUP** | 0.266 | 0.266 | 1 | 61.873 | 0.3604 | 0.55049 | |
| **SESSION** | 1.259 | 0.6295 | 2 | 123.127 | 0.8528 | 0.4287 | |
| **GROUP:SESSION** | 7.0401 | 3.5201 | 2 | 123.127 | 4.7688 | 0.01012 | * |

*Signif. codes: 0 '***' 0.001 '**' 0.01 '*' 0.05 '.' 0.1 ' ' 1*

Normality of residuals:

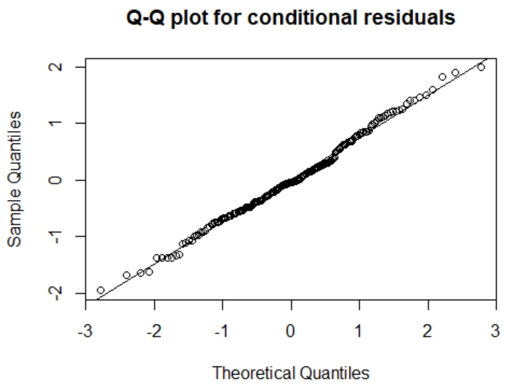

Shapiro-Wilk normality test: p = 0.5238

### Model 2A: Linear mixed model for simple training group GROUP

Formula: Glx_L-SM1[GROUPsimp] ~ SESSION + (1 | SubjectID)

| AIC | BIC | logLik | deviance | df.resid |
|---|---|---|---|---|
| 271 | 283.5 | -130.5 | 261 | 85 |

Random effects:

| Groups | Name | Variance | Std.Dev. |
|---|---|---|---|
| **SubjectID** | (Intercept) | 0.505 | 0.711 |
| **Residual** | | 0.732 | 0.856 |

*Number of obs: 90, groups: SubjectID, 30*

Fixed effects:

| | Estimate | Std. Error | df | t-value | Pr(>\|t\|) | |
|---|---|---|---|---|---|---|
| **(Intercept)** | 10.662 | 0.203 | 67.49 | 52.489 | <2e-16 | *** |
| **SESSION[MID]** | 0.397 | 0.221 | 60.00 | 1.796 | 0.0776 | . |
| **SESSION[POST]** | 0.367 | 0.221 | 60.00 | 1.661 | 0.1019 | |

*Signif. codes: 0 '***' 0.001 '**' 0.01 '*' 0.05 '.' 0.1 ' ' 1*

Type III Analysis of Variance Table with Satterthwaite's method:

| | Sum Sq | Mean Sq | NumDF | DenDF | F value | Pr(>F) |
|---|---|---|---|---|---|---|
| **SESSION** | 2.9306 | 1.4653 | 2 | 60 | 2.0008 | 0.1441 |

*Signif. codes: 0 '***' 0.001 '**' 0.01 '*' 0.05 '.' 0.1 ' ' 1*

Normality of residuals:

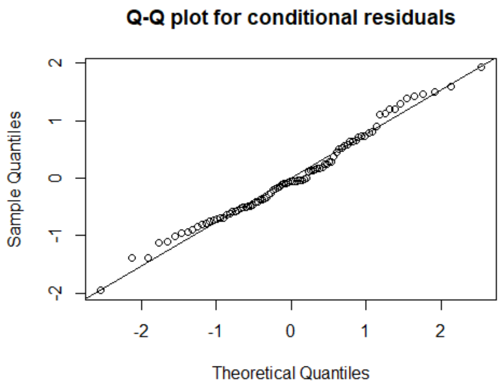

Shapiro-Wilk normality test: p = 0.3295

### Model 2B: Linear mixed model for complex training group GROUP

Formula: Glx_L-SM1[GROUPcomp] ~ SESSION + (1 | SubjectID)

| AIC | BIC | logLik | deviance | df.resid |
|---|---|---|---|---|
| 292.8 | 305.5 | -141.4 | 282.8 | 90 |

Random effects:

| Groups | Name | Variance | Std.Dev. |
|---|---|---|---|
| **SubjectID** | (Intercept) | 0.662 | 0.814 |
| **Residual** | | 0.743 | 0.862 |

*Number of obs: 95, groups: SubjectID, 32*

Fixed effects:

| | Estimate | Std. Error | df | t-value | Pr(>\|t\|) | |
|---|---|---|---|---|---|---|
| **(Intercept)** | 11.319 | 0.212 | 67.50 | 53.412 | < 2e-16 | *** |
| **SESSION[MID]** | -0.208 | 0.218 | 63.26 | -0.955 | 0.3435 | |
| **SESSION[POST]** | -0.581 | 0.218 | 63.26 | -2.669 | 0.0097 | ** |

*Signif. codes: 0 '***' 0.001 '**' 0.01 '*' 0.05 '.' 0.1 ' ' 1*

Type III Analysis of Variance Table with Satterthwaite's method:

| | Sum Sq | Mean Sq | NumDF | DenDF | F value | Pr(>F) | |
|---|---|---|---|---|---|---|---|
| **SESSION** | 5.4611 | 2.7305 | 2 | 63.151 | 3.6728 | 0.03098 | * |

*Signif. codes: 0 '***' 0.001 '**' 0.01 '*' 0.05 '.' 0.1 ' ' 1*

Normality of residuals:

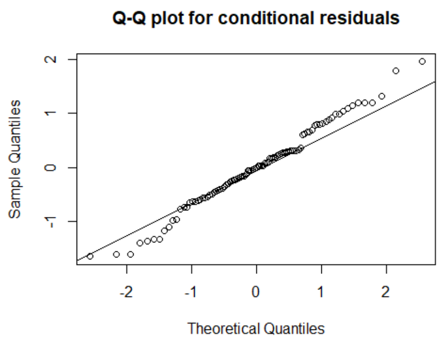

Shapiro-Wilk normality test: p = 0.5784

### Post hoc tests for complex training GROUP

| contrast | mean difference | 95% CI | t-value | df | p-value | significance |
|---|---|---|---|---|---|---|
| **GROUP_complex: PRE - MID** | 0.2089 | -0.2283 – 0.6462 | 0.9759 | 30 | 0.3369 | |
| **GROUP_complex: PRE - POST** | 0.6072 | 0.1347 – 1.0797 | 2.624 | 30 | 0.0135 | * |
| **GROUP_complex: MID - POST** | 0.3734 | -0.0738 – 0.8205 | 1.703 | 31 | 0.0986 | |

*Asterisks indicate results p < α, with Bonferroni correction for 5 tests (α = 0.05/3 = 0.0167).*
*Abbreviations: CI = confidence interval; df = degrees of freedom.*

Appendix 4B: Linear mixed model analysis of motor learning-related changes in resting Glx levels of the right SM1.

### Model 1: Full linear mixed model

Formula: Glx_R-SM1 ~ GROUP + SESSION + GROUP:SESSION + (1 | SubjectID)

| AIC | BIC | logLik | deviance | df.resid |
|---|---|---|---|---|
| 607.1 | 632.9 | -295.6 | 591.1 | 177 |

Random effects:

| Groups | Name | Variance | Std.Dev. |
|---|---|---|---|
| **SubjectID** | (Intercept) | 1.165 | 1.079 |
| **Residual** | | 0.821 | 0.906 |

*Number of obs: 185, groups: SubjectID, 62*

Fixed effects:

| | Estimate | Std. Error | df | t-value | Pr(>\|t\|) | |
|---|---|---|---|---|---|---|
| **(Intercept)** | 11.482 | 0.257 | 109.93 | 44.629 | <0.0001 | *** |
| **GROUP[comp]** | -0.038 | 0.360 | 111.21 | -0.106 | 0.9157 | |
| **SESSION[MID]** | 0.625 | 0.234 | 123.02 | 2.672 | 0.0086 | ** |
| **SESSION[POST]** | 0.213 | 0.234 | 123.02 | 0.909 | 0.3653 | |
| **GROUP[comp]:SESSION[MID]** | -0.294 | 0.327 | 123.24 | -0.896 | 0.3718 | |
| **GROUP[comp]:SESSION[POST]** | 0.040 | 0.327 | 123.24 | 0.121 | 0.9037 | |

*Signif. codes: 0 '***' 0.001 '**' 0.01 '*' 0.05 '.' 0.1 ' ' 1*

Type III Analysis of Variance Table with Satterthwaite's method:

| | Sum Sq | Mean Sq | NumDF | DenDF | F value | Pr(>F) | |
|---|---|---|---|---|---|---|---|
| **GROUP** | 0.1331 | 0.1331 | 1 | 62.00 | 0.162 | 0.6887 | |
| **SESSION** | 7.018 | 3.509 | 2 | 123.17 | 4.273 | 0.0161 | * |
| **GROUP:SESSION** | 1.0234 | 0.5117 | 2 | 123.17 | 0.623 | 0.5380 | |

*Signif. codes: 0 '***' 0.001 '**' 0.01 '*' 0.05 '.' 0.1 ' ' 1*

Normality of residuals:

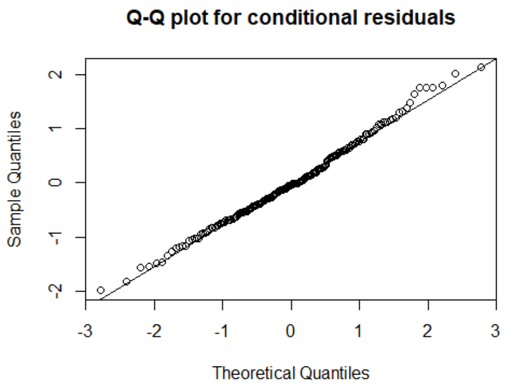

Shapiro-Wilk normality test: p = 0.5243

### Model 2: Removing GROUP:SESSION interaction (based on p = 0.5380 in Model 1)

Formula: Glx_R-SM1 ~ GROUP + SESSION + (1 | SubjectID)

| AIC | BIC | logLik | deviance | df.resid |
|---|---|---|---|---|
| 604.4 | 623.7 | -296.2 | 592.4 | 179 |

Random effects:

| Groups | Name | Variance | Std.Dev. |
|---|---|---|---|
| **SubjectID** | (Intercept) | 1.161 | 1.078 |
| **Residual** | | 0.830 | 0.911 |

*Number of obs: 185, groups: SubjectID, 62*

Fixed effects:

| | Estimate | Std. Error | df | t-value | Pr(>\|t\|) | |
|---|---|---|---|---|---|---|
| **(Intercept)** | 11.525 | 0.239 | 85.87 | 48.284 | <0.0001 | *** |
| **GROUP[comp]** | -0.123 | 0.305 | 62.00 | -0.405 | 0.6871 | |
| **SESSION[MID]** | 0.475 | 0.165 | 123.26 | 2.885 | 0.0046 | ** |
| **SESSION[POST]** | 0.234 | 0.165 | 123.26 | 1.423 | 0.1574 | |

*Signif. codes: 0 '***' 0.001 '**' 0.01 '*' 0.05 '.' 0.1 ' ' 1*

Type III Analysis of Variance Table with Satterthwaite's method:

| | Sum Sq | Mean Sq | NumDF | DenDF | F value | Pr(>F) | |
|---|---|---|---|---|---|---|---|
| **GROUP** | 0.1359 | 0.1359 | 1 | 62.00 | 0.164 | 0.6871 | |
| **SESSION** | 6.9095 | 3.4547 | 2 | 123.18 | 4.164 | 0.0178 | * |

*Signif. codes: 0 '***' 0.001 '**' 0.01 '*' 0.05 '.' 0.1 ' ' 1*

### Model 3: Removing GROUP main effect (based on p = 0.6871 in Model 2) → **FINAL MODEL**

Formula: Glx_R-SM1 ~ SESSION + GROUP:SESSION + (1 | SubjectID)

| AIC | BIC | logLik | deviance | df.resid |
|---|---|---|---|---|
| 602.5 | 618.6 | -296.3 | 592.5 | 180 |

Random effects:

| Groups | Name | Variance | Std.Dev. |
|---|---|---|---|
| **SubjectID** | (Intercept) | 1.165 | 1.079 |
| **Residual** | | 0.830 | 0.911 |

*Number of obs: 185, groups: SubjectID, 62*

Fixed effects:

| | Estimate | Std. Error | df | t-value | Pr(>\|t\|) | |
|---|---|---|---|---|---|---|
| **(Intercept)** | 11.462 | 0.180 | 111.67 | 63.605 | <0.0001 | *** |
| **SESSION[MID]** | 0.474 | 0.165 | 123.28 | 2.884 | 0.0046 | ** |
| **SESSION[POST]** | 0.234 | 0.165 | 123.28 | 1.421 | 0.1578 | |

*Signif. codes: 0 '***' 0.001 '**' 0.01 '*' 0.05 '.' 0.1 ' ' 1*

Type III Analysis of Variance Table with Satterthwaite's method:

| | Sum Sq | Mean Sq | NumDF | DenDF | F value | Pr(>F) | |
|---|---|---|---|---|---|---|---|
| **SESSION** | 6.902 | 3.451 | 2 | 123.20 | 4.159 | 0.0179 | * |

*Signif. codes: 0 '***' 0.001 '**' 0.01 '*' 0.05 '.' 0.1 ' ' 1*

Normality of residuals:

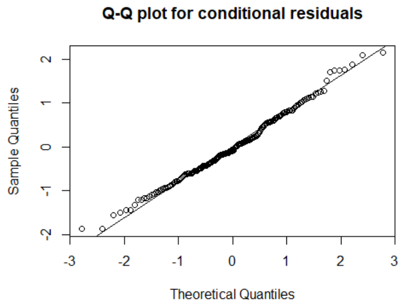

Shapiro-Wilk normality test: p = 0.3703

### Post hoc tests based on Model 3 (pooled for GROUP):

| contrast | mean difference | 95% CI | t-value | df | p-value | significance |
|---|---|---|---|---|---|---|
| **PRE - MID** | -0.4763 | -0.7972 – -0.1553 | -2.968 | 60 | 0.0043 | * |
| **PRE - POST** | -0.2223 | -0.5272 – 0.0826 | -1.459 | 60 | 0.1499 | |
| **MID - POST** | 0.2406 | -0.1252 – 0.6065 | 1.315 | 61 | 0.1933 | |

*Asterisks indicate results p < α, with Bonferroni correction for 5 tests (α = 0.05/3 = 0.0167).*
*Abbreviations: CI = confidence interval; df = degrees of freedom.*

**Appendix 5. Modulation of neurometabolite levels in left PMd.**

Appendix 5A: Linear mixed model analysis of left PMd Glx levels, resulting in a change of pooled left PMd data, but no modulation within each session.

Model 1: Full linear mixed model

Formula: Glx_L-PMd ~ GROUP + SESSION + TASK + GROUP:SESSION + SESSION:TASK + GROUP:TASK + Water_FWHM + (1 | SubjectID)

| AIC | BIC | logLik | deviance | df.resid |
|---|---|---|---|---|
| 1948.7 | 2045.6 | -953.4 | 1906.7 | 723 |

Random effects:

| Groups | Name | Variance | Std.Dev. |
|---|---|---|---|
| **SubjectID** | (Intercept) | 2.378 | 1.542 |
| **Residual** | | 0.545 | 0.738 |

*Number of obs: 744, groups: SubjectID, 62*

Fixed effects:

| | Estimate | Std. Error | df | t-value | Pr(>\|t\|) | |
|---|---|---|---|---|---|---|
| **(Intercept)** | 9.477 | 0.809 | 665.36 | 11.719 | <0.0001 | *** |
| **GROUP[complex]** | 0.194 | 0.415 | 74.50 | 0.469 | 0.6406 | |
| **SESSION[MID]** | -0.126 | 0.149 | 681.88 | -0.841 | 0.4006 | |
| **SESSION[POST]** | -0.047 | 0.149 | 681.92 | -0.315 | 0.7530 | |
| **TASK[BTT1]** | -0.043 | 0.154 | 681.88 | -0.279 | 0.7803 | |
| **TASK[BTT2]** | -0.120 | 0.154 | 681.90 | -0.774 | 0.4389 | |
| **TASK[RESTafter]** | -0.075 | 0.154 | 681.92 | -0.484 | 0.6285 | |
| **Water_FWHM** | 0.190 | 0.074 | 740.28 | 2.574 | 0.0103 | * |
| **GROUP[complex]:SESSION[MID]** | 0.222 | 0.133 | 681.91 | 1.672 | 0.0950 | . |
| **GROUP[complex]:SESSION[POST]** | -0.256 | 0.133 | 682.47 | -1.920 | 0.0553 | . |
| **SESSION[MID]:TASK[BTT1]** | 0.069 | 0.188 | 681.89 | 0.370 | 0.7111 | |
| **SESSION[POST]:TASK[BTT1]** | -0.028 | 0.188 | 681.93 | -0.148 | 0.8826 | |
| **SESSION[MID]:TASK[BTT2]** | 0.125 | 0.188 | 681.88 | 0.665 | 0.5061 | |
| **SESSION[POST]:TASK[BTT2]** | -0.010 | 0.188 | 681.90 | -0.054 | 0.9572 | |
| **SESSION[MID]:TASK[RESTafter]** | 0.038 | 0.188 | 681.95 | 0.203 | 0.8394 | |
| **SESSION[POST]:TASK[RESTafter]** | -0.071 | 0.188 | 681.99 | -0.378 | 0.7055 | |
| **GROUP[complex]:TASK[BTT1]** | 0.140 | 0.153 | 681.88 | 0.912 | 0.3623 | |
| **GROUP[complex]:TASK[BTT2]** | 0.165 | 0.153 | 681.91 | 1.079 | 0.2809 | |
| **GROUP[complex]:TASK[RESTafter]** | 0.050 | 0.153 | 681.93 | 0.329 | 0.7421 | |

*Signif. codes: 0 ‘***’ 0.001 ‘**’ 0.01 ‘*’ 0.05 ‘.’ 0.1 ‘ ’ 1*

Type III Analysis of Variance Table with Satterthwaite's method:

| | Sum Sq | Mean Sq | NumDF | DenDF | F value | Pr(>F) | |
|---|---|---|---|---|---|---|---|
| **GROUP** | 0.256 | 0.256 | 1 | 62.65 | 0.469 | 0.4958 | |
| **SESSION** | 8.497 | 4.248 | 2 | 681.90 | 7.791 | 0.0005 | *** |
| **TASK** | 0.969 | 0.323 | 3 | 682.05 | 0.592 | 0.6201 | |
| **Water_FWHM** | 3.612 | 3.612 | 1 | 740.28 | 6.624 | 0.0103 | * |
| **GROUP:SESSION** | 7.043 | 3.521 | 2 | 682.20 | 6.458 | 0.0017 | ** |
| **SESSION:TASK** | 0.449 | 0.075 | 6 | 681.91 | 0.137 | 0.9914 | |
| **GROUP:TASK** | 0.827 | 0.276 | 3 | 681.90 | 0.506 | 0.6784 | |

*Signif. codes: 0 '***' 0.001 '**' 0.01 '*' 0.05 '.' 0.1 ' ' 1*

Normality of residuals:

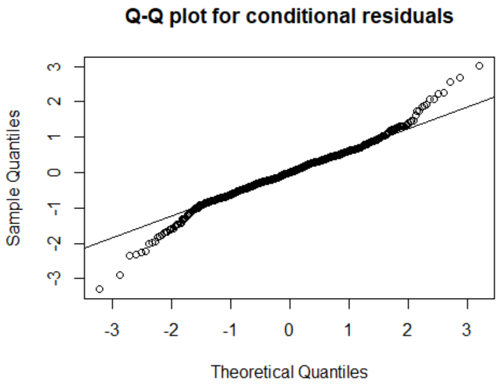

Shapiro-Wilk normality test: p = 8.581e-10

<u>NOTE:</u> Although normality of residuals was not complied with (light tails of distribution), no transformation could correct for this properly (best approximation: quadratic transformation with Shapiro-Wilk p = 2.428e-08). Due to the relatively robust nature of LMMs to deviations from the assumption of normally distributed residuals, the insufficient improvement of the residuals' normality after transformation and the higher interpretability of untransformed data, the untransformed model has been further refined and finally interpreted.

### Model 2: Removing SESSION:TASK interaction (based on p = 0.9914 in Model 1)

Formula: Glx_L-PMd ~ GROUP + SESSION + TASK + GROUP:SESSION + GROUP:TASK + Water_FWHM + (1 | SubjectID)

| AIC | BIC | logLik | deviance | df.resid |
|---|---|---|---|---|
| 1937.5 | 2006.7 | -953.8 | 1907.5 | 729 |

Random effects:

| Groups | Name | Variance | Std.Dev. |
|---|---|---|---|
| **SubjectID** | (Intercept) | 2.378 | 1.542 |
| **Residual** | | 0.546 | 0.739 |

*Number of obs: 744, groups: SubjectID, 62*

Fixed effects:

| | Estimate | Std. Error | df | t-value | Pr(>\|t\|) | |
|---|---|---|---|---|---|---|
| **(Intercept)** | 9.485 | 0.804 | 662.82 | 11.796 | <0.0001 | *** |
| **GROUP[complex]** | 0.194 | 0.415 | 74.52 | 0.467 | 0.6416 | |
| **SESSION[MID]** | -0.067 | 0.095 | 681.90 | -0.707 | 0.4799 | |
| **SESSION[POST]** | -0.074 | 0.096 | 682.32 | -0.774 | 0.4392 | |
| **TASK[BTT1]** | -0.029 | 0.110 | 681.91 | -0.264 | 0.7917 | |
| **TASK[BTT2]** | -0.081 | 0.110 | 681.97 | -0.738 | 0.4610 | |
| **TASK[RESTafter]** | -0.086 | 0.110 | 682.29 | -0.775 | 0.4386 | |
| **Water_FWHM** | 0.189 | 0.074 | 740.27 | 2.551 | 0.0109 | * |
| **GROUP[complex]:SESSION[MID]** | 0.222 | 0.133 | 681.92 | 1.670 | 0.0953 | . |
| **GROUP[complex]:SESSION[POST]** | -0.256 | 0.133 | 682.47 | -1.921 | 0.0552 | . |
| **GROUP[complex]:TASK[BTT1]** | 0.140 | 0.153 | 681.88 | 0.911 | 0.3626 | |
| **GROUP[complex]:TASK[BTT2]** | 0.165 | 0.153 | 681.91 | 1.078 | 0.2813 | |
| **GROUP[complex]:TASK[RESTafter]** | 0.050 | 0.153 | 681.93 | 0.328 | 0.7427 | |

*Signif. codes: 0 '***' 0.001 '**' 0.01 '*' 0.05 '.' 0.1 ' ' 1*

Type III Analysis of Variance Table with Satterthwaite's method:

| | Sum Sq | Mean Sq | NumDF | DenDF | F value | Pr(>F) | |
|---|---|---|---|---|---|---|---|
| **GROUP** | 0.255 | 0.255 | 1 | 62.65 | 0.467 | 0.4970 | |
| **SESSION** | 8.495 | 4.247 | 2 | 681.9 | 7.780 | 0.0005 | *** |
| **TASK** | 0.967 | 0.322 | 3 | 682.05 | 0.590 | 0.6214 | |
| **Water_FWHM** | 3.553 | 3.553 | 1 | 740.27 | 6.508 | 0.0109 | * |
| **GROUP:SESSION** | 7.049 | 3.524 | 2 | 682.20 | 6.456 | 0.0017 | ** |
| **GROUP:TASK** | 0.827 | 0.276 | 3 | 681.90 | 0.505 | 0.6790 | |

*Signif. codes: 0 '***' 0.001 '**' 0.01 '*' 0.05 '.' 0.1 ' ' 1*

### Model 3: Removing GROUP:TASK interaction (based on p = 0.6790 in Model 2)

Formula: Glx_L-PMd ~ GROUP + SESSION + TASK + GROUP:SESSION + Water_FWHM + (1 | SubjectID)

| AIC | BIC | logLik | deviance | df.resid |
|---|---|---|---|---|
| 1933 | 1988.4 | -954.5 | 1909 | 732 |

Random effects:

| Groups | Name | Variance | Std.Dev. |
|---|---|---|---|
| **SubjectID** | (Intercept) | 2.377 | 1.542 |
| **Residual** | | 0.547 | 0.740 |

*Number of obs: 744, groups: SubjectID, 62*

Fixed effects:

| | Estimate | Std. Error | df | t-value | Pr(>\|t\|) | |
|---|---|---|---|---|---|---|
| **(Intercept)** | 9.447 | 0.804 | 662.35 | 11.751 | <0.0001 | *** |
| **GROUP[complex]** | 0.282 | 0.404 | 67.21 | 0.699 | 0.4871 | |
| **SESSION[MID]** | -0.067 | 0.096 | 681.90 | -0.706 | 0.4804 | |
| **SESSION[POST]** | -0.074 | 0.096 | 682.32 | -0.772 | 0.4402 | |
| **TASK[BTT1]** | 0.043 | 0.077 | 681.91 | 0.561 | 0.5753 | |
| **TASK[BTT2]** | 0.004 | 0.077 | 681.94 | 0.053 | 0.9577 | |
| **TASK[RESTafter]** | -0.060 | 0.077 | 682.34 | -0.774 | 0.4394 | |
| **Water_FWHM** | 0.188 | 0.074 | 740.30 | 2.538 | 0.0113 | * |
| **GROUP[complex]:SESSION[MID]** | 0.222 | 0.133 | 681.92 | 1.668 | 0.0958 | . |
| **GROUP[complex]:SESSION[POST]** | -0.256 | 0.133 | 682.47 | -1.919 | 0.0553 | . |

*Signif. codes: 0 ‘***’ 0.001 ‘**’ 0.01 ‘*’ 0.05 ‘.’ 0.1 ‘ ’ 1*

Type III Analysis of Variance Table with Satterthwaite's method:

| | Sum Sq | Mean Sq | NumDF | DenDF | F value | Pr(>F) | |
|---|---|---|---|---|---|---|---|
| **GROUP** | 0.255 | 0.255 | 1 | 62.66 | 0.466 | 0.4975 | |
| **SESSION** | 8.494 | 4.247 | 2 | 681.9 | 7.762 | 0.0005 | *** |
| **TASK** | 0.995 | 0.332 | 3 | 682.05 | 0.606 | 0.6112 | |
| **Water_FWHM** | 3.525 | 3.525 | 1 | 740.3 | 6.442 | 0.0113 | * |
| **GROUP:SESSION** | 7.052 | 3.526 | 2 | 682.20 | 6.444 | 0.0017 | ** |

*Signif. codes: 0 ‘***’ 0.001 ‘**’ 0.01 ‘*’ 0.05 ‘.’ 0.1 ‘ ’ 1*

Model 4: Removing TASK main effect (based on p = 0.6112 in Model 3) → **FINAL MODEL**

Formula: Glx_L-PMd ~ GROUP + SESSION + GROUP:SESSION + Water_FWHM + (1 | SubjectID)

| AIC | BIC | logLik | deviance | df.resid |
|---|---|---|---|---|
| 1928.9 | 1970.4 | -955.4 | 1910.9 | 735 |

Random effects:

| Groups | Name | Variance | Std.Dev. |
|---|---|---|---|
| **SubjectID** | (Intercept) | 2.376 | 1.541 |
| **Residual** | | 0.549 | 0.741 |

*Number of obs: 744, groups: SubjectID, 62*

Fixed effects:

| | Estimate | Std. Error | df | t-value | Pr(>\|t\|) | |
|---|---|---|---|---|---|---|
| **(Intercept)** | 9.504 | 0.804 | 662.18 | 11.827 | <0.0001 | *** |
| **GROUP[complex]** | 0.280 | 0.404 | 67.23 | 0.694 | 0.4904 | |
| **SESSION[MID]** | -0.067 | 0.096 | 681.90 | -0.704 | 0.4818 | |
| **SESSION[POST]** | -0.073 | 0.096 | 682.32 | -0.766 | 0.4441 | |
| **Water_FWHM** | 0.182 | 0.074 | 740.24 | 2.463 | 0.0140 | * |
| **GROUP[complex]:SESSION[MID]** | 0.222 | 0.133 | 681.92 | 1.664 | 0.0965 | . |
| **GROUP[complex]:SESSION[POST]** | -0.257 | 0.134 | 682.47 | -1.923 | 0.0548 | . |

*Signif. codes: 0 '***' 0.001 '**' 0.01 '*' 0.05 '.' 0.1 ' ' 1*

Type III Analysis of Variance Table with Satterthwaite's method:

| | Sum Sq | Mean Sq | NumDF | DenDF | F value | Pr(>F) | |
|---|---|---|---|---|---|---|---|
| **GROUP** | 0.251 | 0.251 | 1 | 62.66 | 0.457 | 0.5015 | |
| **SESSION** | 8.487 | 4.243 | 2 | 681.91 | 7.734 | 0.0005 | *** |
| **Water_FWHM** | 3.328 | 3.328 | 1 | 740.24 | 6.065 | 0.0140 | * |
| **GROUP:SESSION** | 7.072 | 3.536 | 2 | 682.2 | 6.445 | 0.0017 | ** |

*Signif. codes: 0 '***' 0.001 '**' 0.01 '*' 0.05 '.' 0.1 ' ' 1*

Normality of residuals:

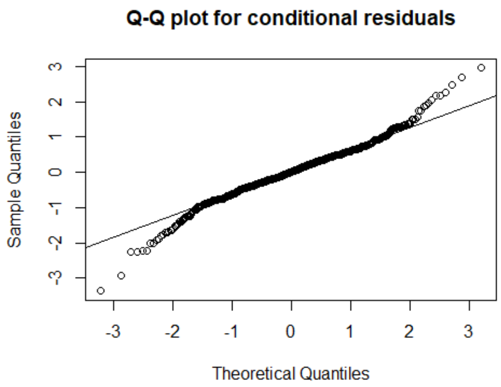

Shapiro-Wilk normality test: p = 1.195e-09

Visualization of the final results (mean value of the four acquisitions of left PMd Glx level per subject):

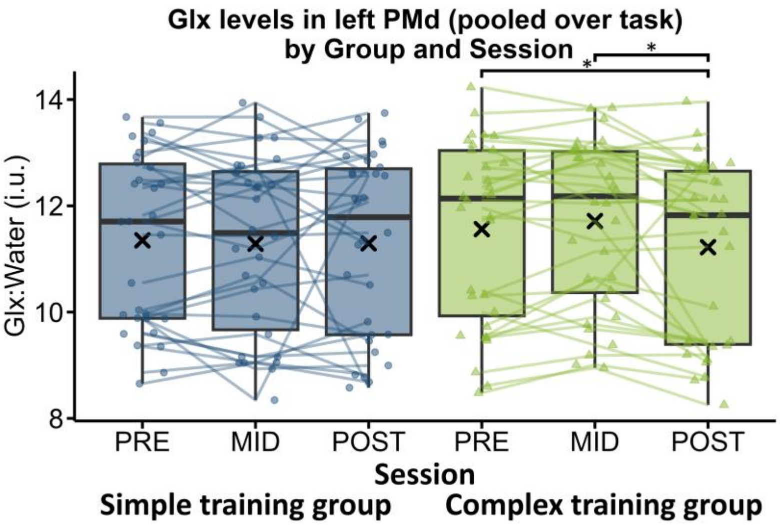

Post hoc tests:

GROUP comparison per measurement SESSION:

| Contrast (S - C) | mean difference | 95% CI | t-value | df | p-value | significance |
|---|---|---|---|---|---|---|
| **PRE** | -0.2800 | -1.099 – 0.539 | -0.539 | 69.6 | 0.4974 | |
| **MID** | -0.5016 | -1.321 – 0.318 | -1.221 | 69.7 | 0.2261 | |
| **POST** | -0.0232 | -0.844 – 0.797 | -0.056 | 70.2 | 0.9553 | |

*Asterisks indicate results p < α, with Bonferroni correction for 5 tests (α = 0.05/3 = 0.0167).*
*Abbreviations: C = complex training group; CI = confidence interval; df = degrees of freedom; S = simple training group.*

SESSION comparison for simple training group:

| contrast | mean difference | 95% CI | t-value | df | p-value | significance |
|---|---|---|---|---|---|---|
| **GROUP_simple: PRE - MID** | 0.0673 | -0.121 – 0.256 | 0.701 | 687 | 0.4834 | |
| **GROUP_simple: PRE - POST** | 0.0734 | -0.116 – 0.262 | 0.763 | 687 | 0.4458 | |
| **GROUP_simple: MID - POST** | 0.0061 | -0.183 – 0.195 | 0.063 | 687 | 0.9495 | |

*Asterisks indicate results p < α, with Bonferroni correction for 5 tests (α = 0.05/3 = 0.0167).*
*Abbreviations: CI = confidence interval; df = degrees of freedom.*

SESSION comparison for complex training group:

| contrast | mean difference | 95% CI | t-value | df | p-value | significance |
|---|---|---|---|---|---|---|
| **GROUP_complex: PRE - MID** | -0.1542 | -0.337 – 0.028 | -1.660 | 687 | 0.0974 | |
| **GROUP_complex: PRE - POST** | 0.3303 | 0.148 – 0.513 | 3.551 | 687 | 0.0004 | * |
| **GROUP_complex: MID - POST** | 0.4845 | 0.302 – 0.667 | 5.212 | 687 | <0.0001 | * |

*Asterisks indicate results p < α, with Bonferroni correction for 5 tests (α = 0.05/3 = 0.0167).*
*Abbreviations: CI = confidence interval; df = degrees of freedom.*

Appendix 5B: Visualization of task-related changes in GABA+ and Glx per subject and session.

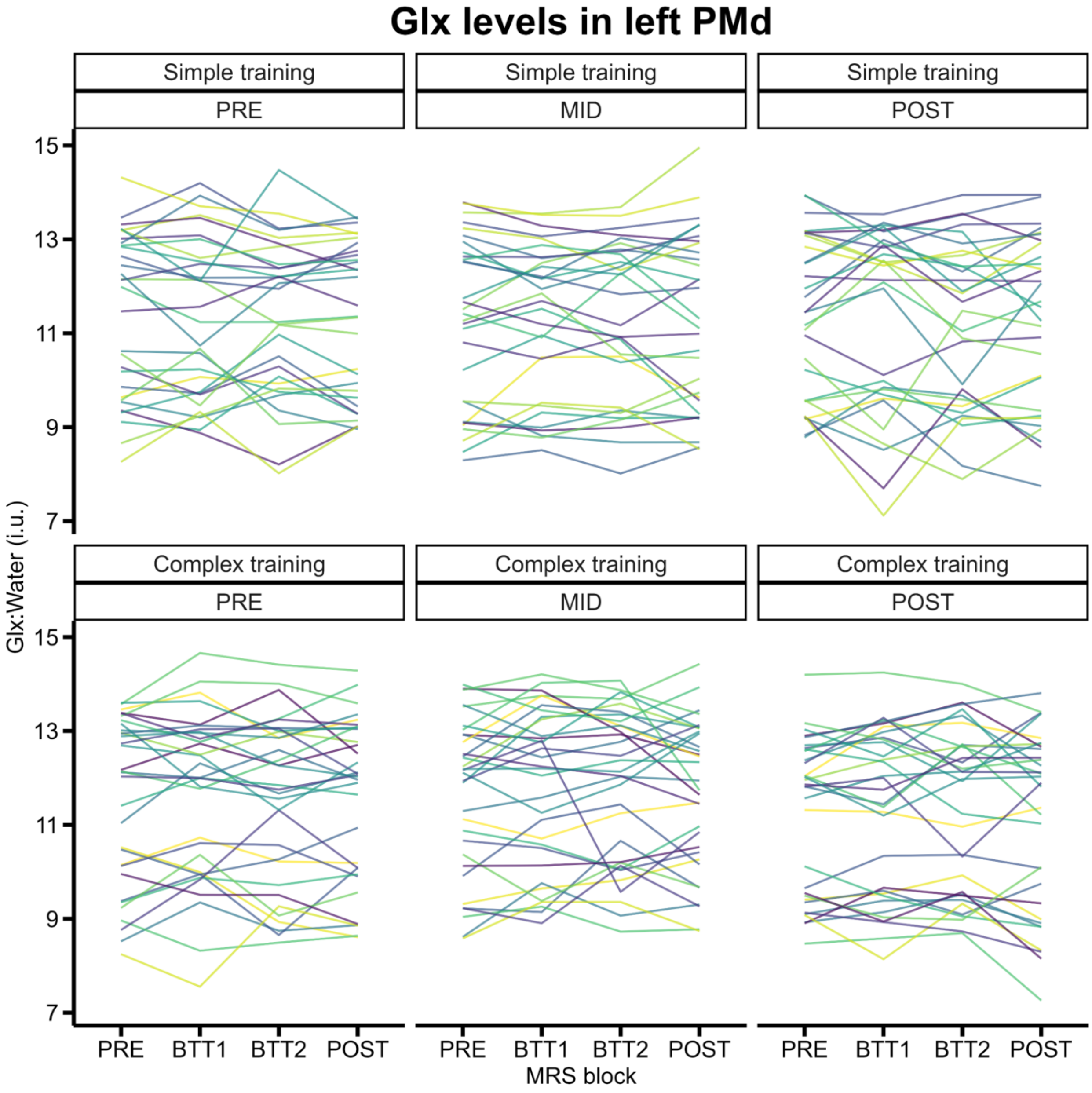

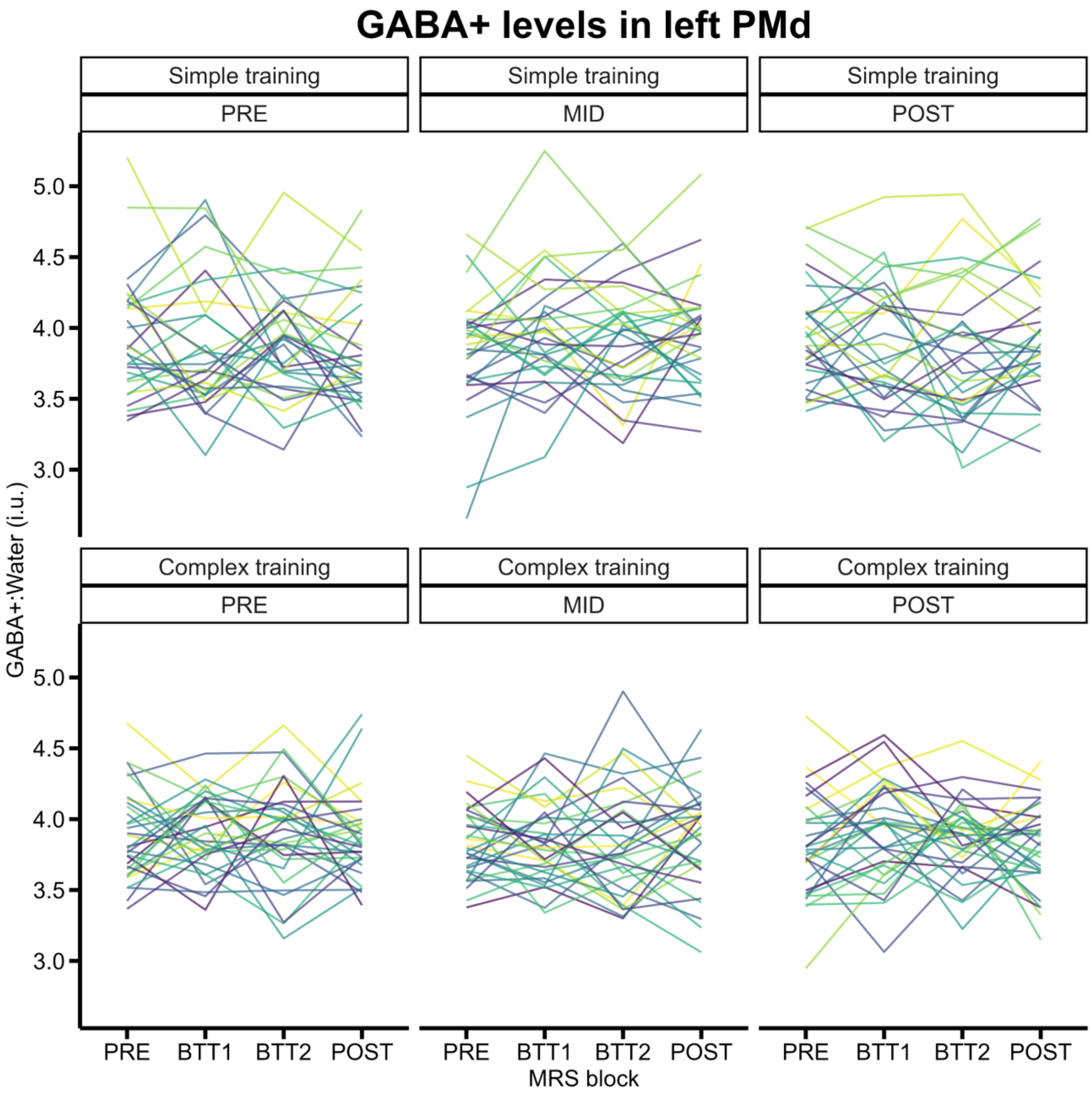
GABA+ levels in left PMd
Simple training
PRE
Simple training
MID
Simple training
POST
Complex training
PRE
Complex training
MID
Complex training
POST
GABA+:Water (i.u.)
5.0
4.5
4.0
3.5
3.0
PRE
BTT1
BTT2
POST
MRS block

Appendix 5C: Multiple linear regression analysis details for the prediction of short-term motor learning based on task-related GABA+ and Glx modulation in left dorsal premotor cortex (PMd).

### Model 1: Full model multiple linear regression

Formula: $MR\_BTTslope_{allLines}$ ~ GABA_MODmax_abs + Glx_MODmax_abs + ~ GABA_MODmax_abs:Glx_MODmax_abs

Coefficients:

| | Estimate | Std. Error | t-value | Pr(>\|t\|) | |
|---|---|---|---|---|---|
| **(Intercept)** | -0.0194 | 0.0556 | -0.349 | 0.7286 | |
| **GABA_MODmax_abs** | 0.2395 | 0.1060 | 2.260 | 0.0276 | * |
| **Glx_MODmax_abs** | 0.1288 | 0.0649 | 1.984 | 0.0520 | . |
| **GABA_MODmax_abs:Glx_MODmax_abs** | -0.1880 | 0.1189 | -1.581 | 0.1193 | |

*Signif. codes: 0 '***' 0.001 '**' 0.01 '*' 0.05 '.' 0.1 ' ' 1*
*Residual standard error: 0.1019 on 58 degrees of freedom*
*Multiple R-squared: 0.1151, Adjusted R-squared: 0.0693*
*F-statistic: 2.514 on 3 and 58 DF, p-value: 0.06725*

Anova Table (Type III tests)

| | Sum Sq | DF | F value | Pr(>F) | |
|---|---|---|---|---|---|
| **(Intercept)** | 0.00126 | 1 | 0.122 | 0.7286 | |
| **GABA_MODmax_abs** | 0.05301 | 1 | 5.109 | 0.0276 | * |
| **Glx_MODmax_abs** | 0.04085 | 1 | 3.936 | 0.0520 | . |
| **GABA_MODmax_abs:Glx_MODmax_abs** | 0.02594 | 1 | 2.500 | 0.1193 | |
| **Residuals** | 0.60183 | 58 | | | |

*Signif. codes: 0 '***' 0.001 '**' 0.01 '*' 0.05 '.' 0.1 ' ' 1*

Check assumptions:

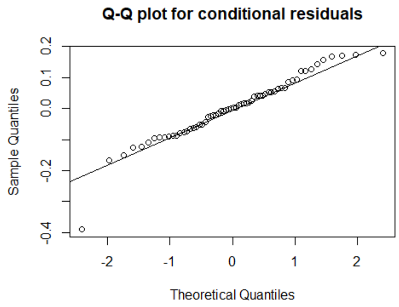

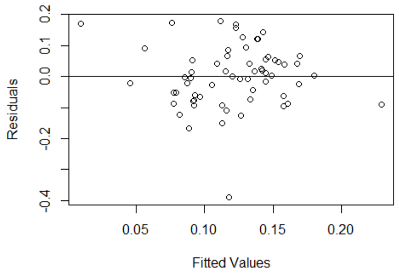

Shapiro-Wilk normality test: p = 0.0100

studentized Breusch-Pagan test: p = 0.3690

Model 2: Full model multiple linear regression with removed outlier (lowest data point in QQ-plot and predicted-by-residual plot of Model 1) → **FINAL MODEL**

Formula: MR_BTTslopeAllLines ~ GABA_MODmax_abs + Glx_MODmax_abs + ~ GABA_MODmax_abs:Glx_MODmax_abs

Coefficients:

| | Estimate | Std. Error | t-value | Pr(>\|t\|) | |
|---|---|---|---|---|---|
| **(Intercept)** | -0.02454 | 0.04793 | -0.512 | 0.6106 | |
| **GABA_MODmax_abs** | 0.24508 | 0.09125 | 2.686 | 0.0095 | ** |
| **Glx_MODmax_abs** | 0.16621 | 0.05649 | 2.942 | 0.0047 | ** |
| **GABA_MODmax_abs:Glx_MODmax_abs** | -0.24415 | 0.10310 | -2.368 | 0.0213 | * |

*Signif. codes: 0 '***' 0.001 '**' 0.01 '*' 0.05 '.' 0.1 ' ' 1*
*Residual standard error: 0.08771 on 57 degrees of freedom*
*Multiple R-squared: 0.161, Adjusted R-squared: 0.1168*
*F-statistic: 3.645 on 3 and 57 DF, p-value: 0.01783*

Anova Table (Type III tests)

| | Sum Sq | DF | F value | Pr(>F) | |
|---|---|---|---|---|---|
| **(Intercept)** | 0.00202 | 1 | 0.262 | 0.6106 | |
| **GABA_MODmax_abs** | 0.05551 | 1 | 7.214 | 0.0095 | ** |
| **Glx_MODmax_abs** | 0.06661 | 1 | 8.657 | 0.0047 | ** |
| **GABA_MODmax_abs:Glx_MODmax_abs** | 0.04315 | 1 | 5.608 | 0.0213 | * |
| **Residuals** | 0.43855 | 57 | | | |

*Signif. codes: 0 '***' 0.001 '**' 0.01 '*' 0.05 '.' 0.1 ' ' 1*

Check assumptions:

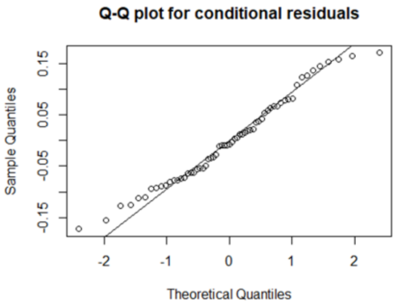

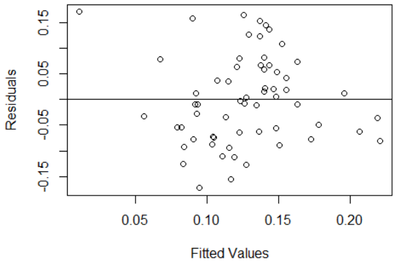

Shapiro-Wilk normality test: p = 0.2753

studentized Breusch-Pagan test: p = 0.1498
